# Domain-adapted language model using reinforcement learning for various dementias

**DOI:** 10.64898/2026.03.17.26348154

**Authors:** Sahana S. Kowshik, Varuna H. Jasodanand, Matteo Bellitti, Shreyas Puducheri, Lingyi Xu, Yi Liu, Ketan S. Saichandran, Brigid C. Dwyer, Audrey Gabelle, Honglin Hao, Sachin Kedar, Daniel L. Murman, Sarah A. O’Shea, Marie-Helene Saint-Hilaire, Niyatee P. Samudra, Emmett A. Sartor, Arun Swaminathan, Olga Taraschenko, Jing Yuan, Rhoda Au, Vijaya B. Kolachalama

## Abstract

Large language models excel at processing complex clinical data and advanced reasoning, yet domain-specific adaptation is essential to realize their full potential in fields such as Alzheimer’s disease and related dementias (ADRD). Here, we present a generative language model for ADRD fine-tuned via reinforcement learning with verifiable rewards using a self-certainty-aware advantage. Model development and validation leveraged data from five ADRD cohorts, totaling 54, 535 participants. Our framework integrates demographics, personal and family medical histories, medication use, neuropsychological test results, functional assessments, physical and neurological examination findings, laboratory data and multimodal neuroimaging to construct comprehensive clinical profiles. On held-out testing data involving 36, 688 participants, our model achieved robust performance on syndromic classification, primary etiological diagnosis and biomarker prediction. Model predictions were validated against postmortem-confirmed diagnoses, and clinical utility was demonstrated in a controlled within-subjects crossover study where board-certified neurologists reviewed cases with and without model assistance, showing that exposure to model responses improved diagnostic performance. These results demonstrate that targeted domain adaptation with reinforcement learning can enable language models to deliver accurate, reasoning-driven support in ADRD evaluation. Prospective validation will be essential to translate these advances into improved patient outcomes.

---

Alzheimer’s disease (AD) and related dementias (ADRD) continue to affect millions globally, creating an escalating public health crisis. The increasing caseload of ADRD cases poses a significant challenge, which is exacerbated by a shortage of skilled professionals trained to manage these complex conditions (*1–4*). These neurodegenerative conditions are marked by overlapping symptoms, diverse etiologies and progressive decline, making early and accurate differential diagnosis particularly challenging. This diagnostic complexity, combined with the scarcity of specialized care, often leads to delayed interventions, high screen failure rates in clinical trials (*5*), and suboptimal patient outcomes. While existing diagnostic tools like magnetic resonance imaging (MRI), positron emission tomography (PET) scans, cerebrospinal fluid (CSF) analysis, and emerging blood-based biomarkers provide valuable insights, they are limited in scope, typically focusing on isolated aspects of disease pathology (*6*). MRI is widely used to detect structural changes in the brain, primarily serving to identify findings indicative of pathologies like AD-related brain atrophy and to rule out non-degenerative structural lesions (*7, 8*). However, while valuable, MRI often lacks the sensitivity required to differentiate between various forms of dementia. PET scans effectively detect amyloid plaques and tau tangles but are costly and limited in capturing co-occurring pathologies (*9*). CSF biomarkers provide biochemical evidence of neurodegeneration but are invasive, lack spatial resolution, and primarily reflect AD-specific pathology (*10*). Similarly, blood-based biomarkers, though scalable, lack regional specificity and are influenced by non-neurological factors (*11*). Importantly, all these modalities are largely insensitive to mixed or non-AD etiologies, limiting their utility in differentiating complex neurodegenerative processes (*12*). Despite these challenges, the recent approval of disease-modifying therapies targeting A*β* has provided renewed optimism. However, ADRD often presents as mixed dementia (*13–15*), where overlapping vascular, tau and amyloid pathologies complicate diagnosis and treatment selection. Effective therapy depends on identifying the predominant pathology, yet many patients exhibit coexisting neurodegenerative processes that may not respond uniformly to a single targeted treatment. This underscores the need for improved diagnostic frameworks that integrate diverse data sources to enhance ADRD assessment.

Recent advancements in large language models (LLMs) have shown promise in medical applications (*16–20*), including generating clinical notes, summarizing patient histories, interpreting imaging reports, and assisting in clinical decision-making by synthesizing information from medical literature. These general-purpose medical LLMs, trained on extensive datasets, are proficient at encoding medical knowledge and performing complex reasoning across diverse clinical domains. However, their substantial computational and financial costs, often requiring vast resources for training, inference and deployment, render them inefficient for many targeted medical applications. This limitation is especially pronounced in settings that require task-specific precision, as general-purpose architectures can lead to higher operational overhead and limited adaptability to domain-specific intricacies (*21–23*). Effectively tackling such targeted clinical problems requires the analysis of complex, multimodal patterns that involve the integration of specialized data such as neuroimaging, cognitive assessments and clinical histories. A bespoke disease-specific small language model (SLM) tailored for ADRD could provide a more detailed and dynamic understanding of the disease, support more accurate differential diagnoses, and inform personalized treatment strategies. All of this can be achieved while offering greater efficiency, reduced computational costs, and enhanced performance in specialized contexts compared to relying on massive general-purpose models (*24, 25*). Fine-tuning such a model via supervised fine-tuning (SFT) is a natural first approach (*26, 27*), but generating high-quality chain-of-thought (CoT) reasoning data is costly and labor-intensive. SFT constrains the model’s reasoning to patterns present in the training data, limiting generalization to novel clinical presentations. Reinforcement learning with verifiable rewards (RLVR), by contrast, enables the model to develop reasoning capabilities without requiring expensive, annotated CoT data, offering better generalization and adaptability in out-of-distribution settings (*28, 29*). Recent work has demonstrated that RLVR can elicit strong reasoning even in smaller, resource-constrained models (*30*), making it particularly well-suited for building an efficient, domain-specific SLM for ADRD assessment.

In this work, we introduce LUNAR (Language model for Unified Neurological Assessment and Reasoning), a domain-adapted language model fine-tuned with RLVR using self-certainty-aware advantage for ADRD assessment, integrating diverse multimodal data from standard neurology evaluations. Our framework processes demographics, medical and family histories, medication use, neuropsychological assessments, functional evaluations and multisequence magnetic resonance imaging to construct a comprehensive cognitive profile of an individual. By synthesizing these inputs, LUNAR provides standardized assessments that may improve diagnostic accuracy and support early interventions. This framework has the potential to assist clinicians in decision-making across the care continuum, from early diagnosis to long-term management. Prospective studies are needed to further evaluate its broader impact on healthcare outcomes.

## Results

Our domain-specific language model (LUNAR) was developed by fine-tuning a compact 3-billion-parameter model (Qwen2.5-3B-Instruct, Q3B) using RLVR on a large multimodal dataset from the NACC cohort (table S1, fig. S1). The training framework incorporated chain-of-thought prompting, a self-certainty–aware advantage and task-specific oversampling of rare etiologies to promote human-interpretable reasoning and concise outputs (Fig. 1, figs. S2 - S9, table S2). Evaluations were performed on held-out cases from the NACC cohort (internal validation) and four independent external cohorts (tables S3 - S8). Whenever feasible, we compared LUNAR against two general-purpose baselines: the original Q3B (used as the base for fine-tuning) and a larger 7-billion-parameter model (Qwen2.5-7B-Instruct or Q7B). LUNAR demonstrated advantages in generation efficiency and clinical task performance, highlighting how RL can refine reasoning traces, improve calibration and allow a compact model to rival or surpass larger baselines (Fig. 2, figs. S10 & S11). Assessments encompassed syndromic classification of cognitive status and primary etiological diagnosis; prediction of key biomarkers, including A*β* PET imaging, cerebrospinal fluid A*β* levels, and dopamine transporter (DaT) imaging evidence; and, where autopsy data were available, direct comparison of etiological predictions against neuropathological evidence (Fig. 3, figs. S12 - S15, tables S9 - S19). Clinical interpretability and practical utility were further evaluated through a blinded head-to-head comparison by expert neurologists, who assessed model-generated diagnostic reasoning summaries against reference clinical summaries (Fig. 4, figs. S16 - S22, table S20).

**Figure 1:**
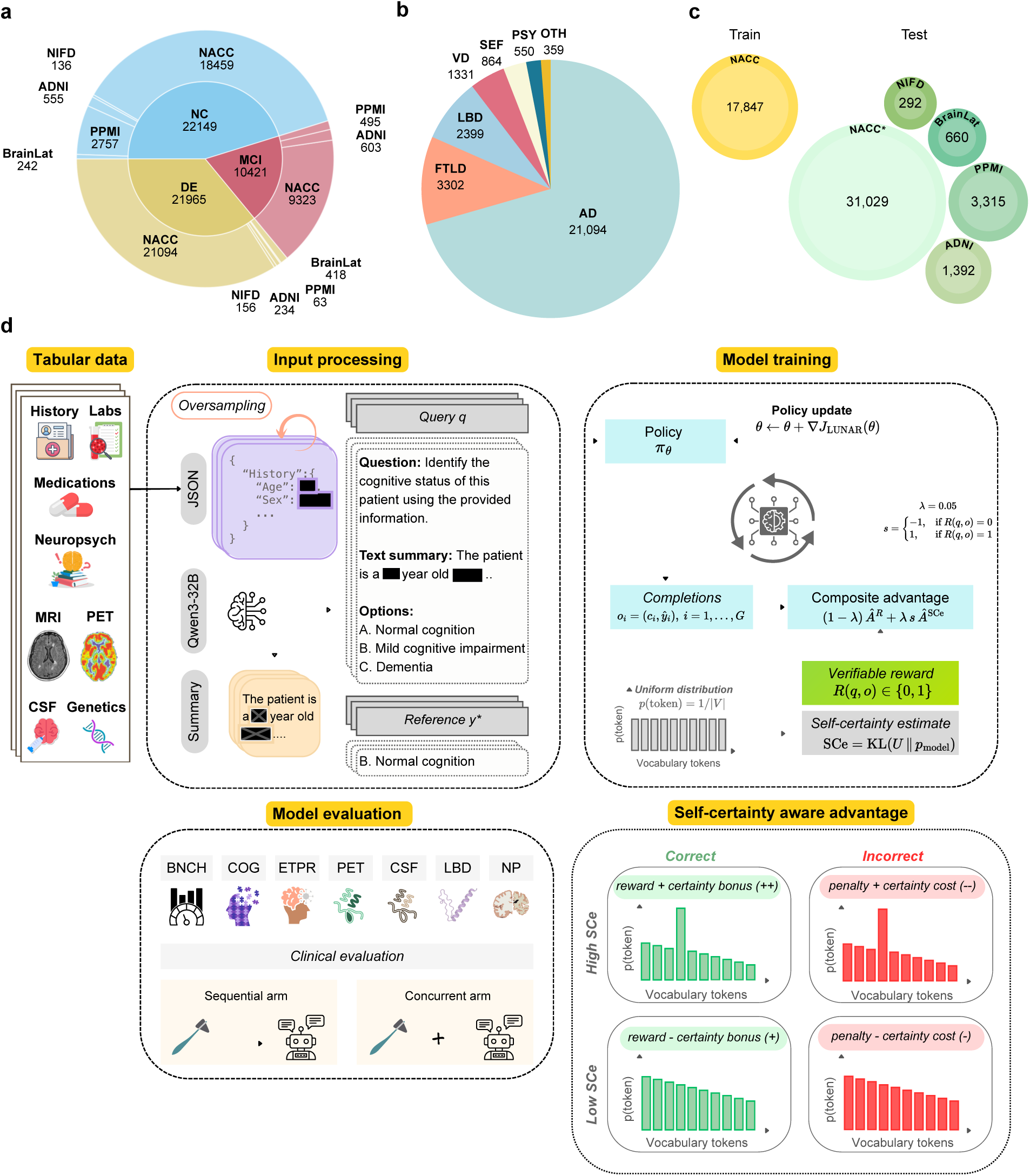
LUNAR framework. (a) Overlap and breakdown of study populations across cohorts used for model development, training and evaluation. Venn diagram shows participant numbers from key sources (NACC, ADNI, BrainLat, NIFD and PPMI), stratified by cognitive status: normal cognition (NC), mild cognitive impairment (MCI), or dementia (DE). Numbers indicate the total unique participants contributed by each cohort. (b) Primary etiologic breakdown among cases with cognitive impairment, displayed as a pie chart with proportions for Alzheimer’s disease (AD), frontotemporal lobar degeneration and its variants, including primary progressive aphasia, corticobasal degeneration and progressive supranuclear palsy, and with or without amyotrophic lateral sclerosis (FTLD), Lewy body disease and/or Parkinson’s disease (LBD), vascular brain injury or vascular dementia including stroke (VD), systemic and environmental factors including infectious diseases (HIV included), metabolic, substance abuse / alcohol, medications, systemic disease and delirium (SEF), psychiatric conditions including schizophrenia, depression, bipolar disorder, anxiety and posttraumatic stress disorder (PSY), and other conditions including moderate/severe traumatic brain injury, repetitive head injury and chronic traumatic encephalopathy, neoplasms, Down syndrome, multiple systems atrophy, Huntington’s disease and seizures, normal pressure hydrocephalus and Prion disease including Creutzfeldt-Jakob disease (OTH). (c) Cohort partitioning for model training, internal testing and external validation. Training utilized data from NACC. Internal testing drew from held-out NACC subsets; external validation included data from ADNI, BrainLat, NIFD and PPMI. (d) Overview of the LUNAR training pipeline and reinforcement learning framework. Multimodal clinical inputs (demographics/history, labs, medications, neuropsychological testing, MRI/PET neuroimaging, CSF biomarkers, genetics) were processed into structured JSON-like queries. A large language model was used to generate textual summaries. These generated summaries, together with a corresponding question and a set of diagnostic options, were then used as inputs to train LUNAR. Bottom panels illustrate model evaluation: benchmark performance across modalities/domains (BNCH: biomarkers, COG: cognition, ETPR: etiology/pathology, PET, CSF, LBD, NP: neuropathology) and clinical human-in-the-loop assessment in sequential and concurrent arms, comparing unaugmented versus augmented performance.

**Figure 2:**
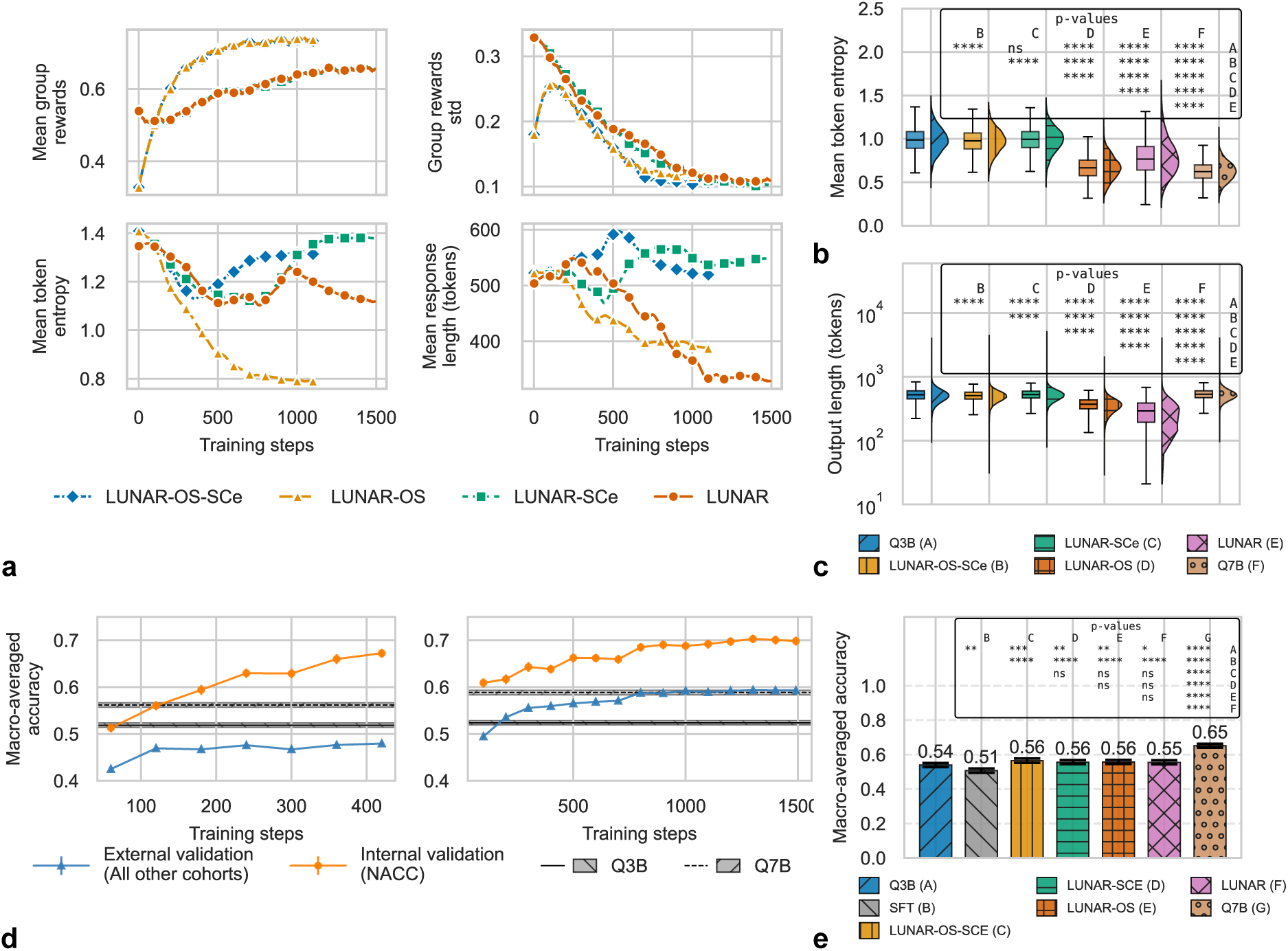
Benefits of reinforcement learning. (a) Training curves of LUNAR and its ablations across optimization steps: mean group rewards (top left), reward standard deviation (top right), mean token entropy (bottom left), and mean response length in tokens (bottom right) for LUNAR, LUNAR–OS, LUNAR–SCe, and LUNAR–OS–SCe. Curves illustrate the impact of oversampling (OS) and self-certainty estimation (SCe) on reward optimization, calibration, and conciseness. (b) Distribution of mean token entropy (mean Shannon entropy across generated tokens, approximated using the top-20 tokens per position) across a random sample of testing data (up to 1,000 cases randomly sampled per testing cohort across NIFD, ADNI, NACC, PPMI, and BrainLat, with each entry representing a unique case–task pair) for LUNAR, its ablations, and two baseline models (Q3B and Q7B). Violin plots with embedded boxplots show medians and interquartile ranges. (c) Distribution of output lengths (tokens) aggregated across all tasks and datasets for the same models. Violin plots with embedded boxplots show medians and interquartile ranges. Pairwise significance for (b) and (c) panels (pairwise two-sided Welch’s t-test, FDR-corrected via Benjamini-Hochberg procedure at *α* = 0.05) is indicated by asterisks above the plot. (d) Macro–averaged accuracy as a function of training steps during SFT (left, fig. S4) and LUNAR (right, fig. S3). Performance is shown separately for the internal validation set (NACC; orange) and external validation cohorts (blue), with 95% bootstrap confidence intervals (non–parametric bootstrap, 1, 000 iterations resampling rows within task–cohort combinations before macro-averaging). Dashed gray lines and shaded areas represent baseline performance (Q3B and Q7B) with their 95% bootstrap CIs (baselines do not distinguish internal and external validation, as they were not trained). (e) Macro-averaged accuracy across five medical benchmarks (MedQA, MedMCQA, MedExpQA, MMLU–Clinical Knowledge and MMLU–Anatomy) for LUNAR, its ablations, SFT, and two baselines (Q3B and Q7B). All models were evaluated using chain-of-thought prompting (fig. S3), except for the SFT model, which was prompted to provide only the final answer (fig. S4). Bars show macro-averaged accuracy with 95% bootstrap percentile confidence intervals (1,000 iterations), computed via non–parametric bootstrap resampling of rows within each benchmark prior to macro-averaging. Statistical significance of differences in medians is assessed via non–parametric permutation tests (10, 000 permutations). Significance levels are denoted as ‘ns’ (not significant) for *p* ≥ 0.05; * for *p <* 0.05; ** for *p <* 0.01; *** for *p <* 0.001; and **** for *p <* 0.0001.

**Figure 3:**
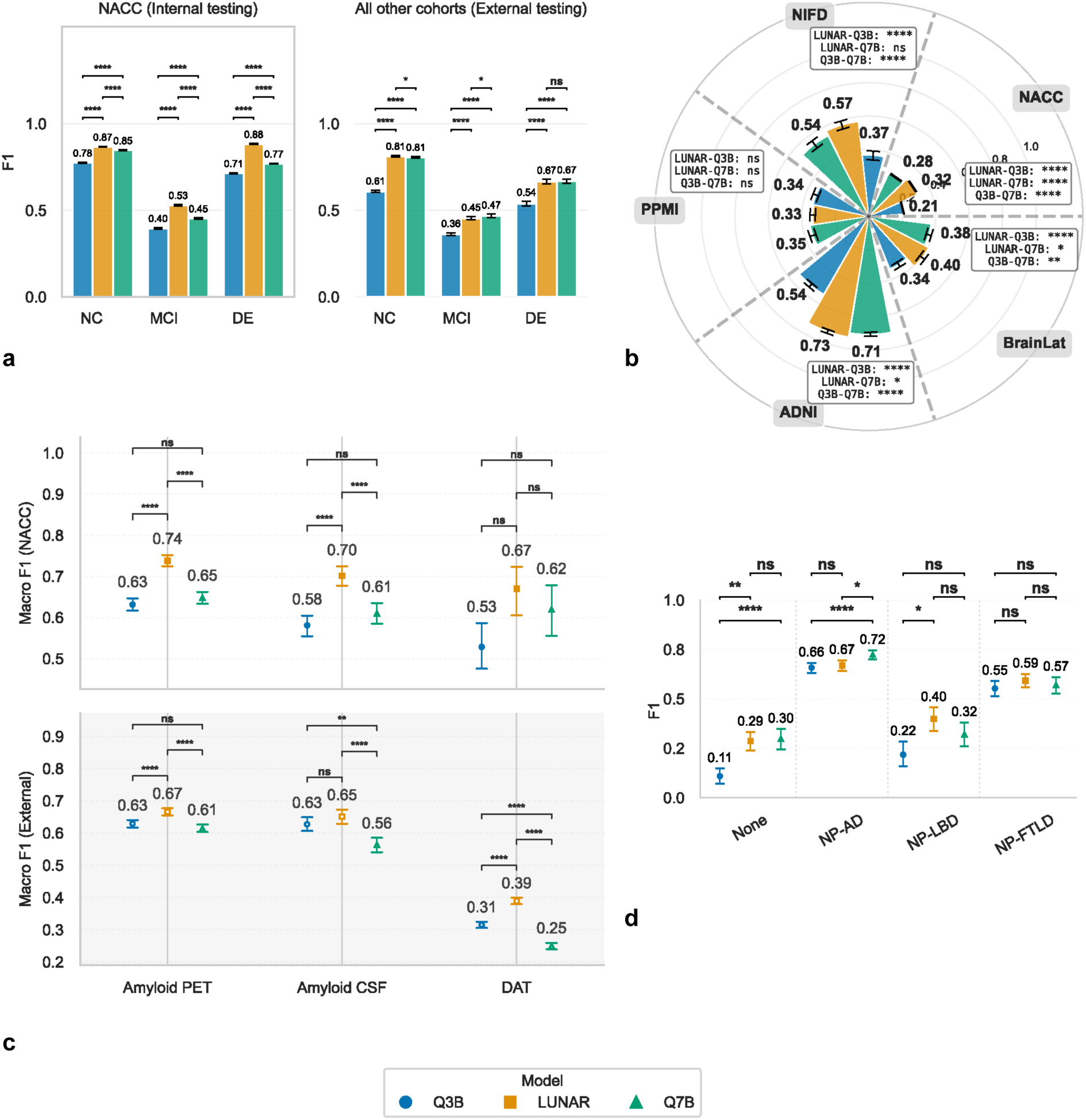
Performance of LUNAR. (a) Class-wise F1 scores for the cognitive classification task COG: normal cognition (NC), mild cognitive impairment (MCI), dementia (DE) on internal testing (NACC; left) and external cohorts (all others; right). Bars represent medians from 1,000 bootstrap samples, with error bars showing 95% bootstrap confidence intervals (2.5th and 97.5th percentiles). (b) Radial (circular) bar plot of macro F1 scores for the primary etiology classification task (ETPR) across cohorts (NACC, ADNI, BrainLat, NIFD and PPMI), comparing LUNAR against baseline models (Q3B and Q7B). Each sector corresponds to a cohort, with bars indicating the medians and 95% bootstrap CIs; statistical annotations reflect pairwise comparisons. (c) Point-and-error plots summarizing macro F1 scores for biomarker positivity classification tasks on amyloid PET, amyloid CSF, and DaT imaging. Results are stratified by internal and external validation sets. Internal validation for NACC included 808 subjects for PET, 288 for CSF, and 71 for DAT. External sets included ADNI for PET (*N* = 1392) and CSF (*N* = 406), and PPMI for DAT (*N* = 1276). Points and lines denote medians and 95% bootstrap CIs. (d) Class-wise F1 scores for the neuropathology one-class (NP-ONE) task (etiologies: None, AD, LBD, FTLD), with medians of the 95% bootstrap CIs shown as point estimates. Across panels, statistical significance of pairwise differences (LUNAR versus Q3B/Q7B) is from non–parametric, two-sided permutation tests (10, 000 permutations at the participant level). The p-values were adjusted using false discovery rate (FDR) correction via the Benjamini-Hochberg procedure, applied within each cohort, class, and metric for panels (a), (c), and (d), and within each cohort and metric for panel (b). Significance levels are denoted as ‘ns’ (not significant) for *p* ≥ 0.05; * for *p* < 0.05; ** for *p <* 0.01; *** for *p <* 0.001; and **** for *p <* 0.0001.

**Figure 4:**
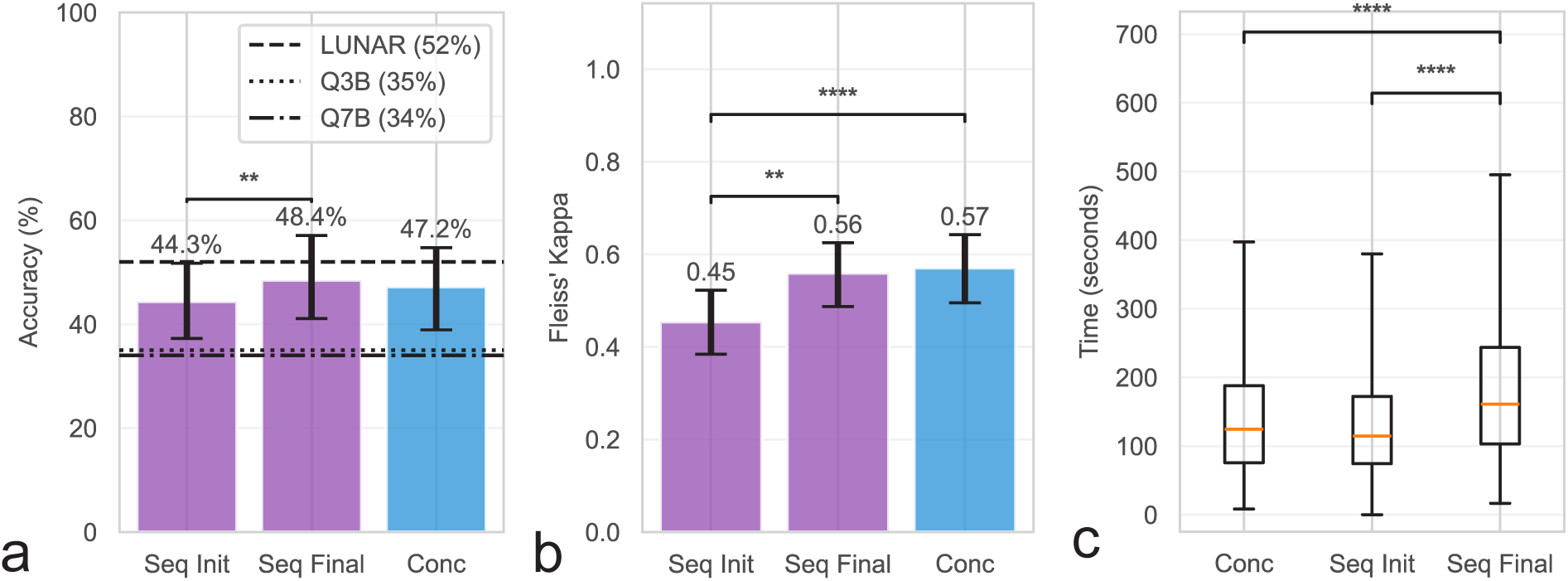
LUNAR-augmented neurologist assessments. (a) Diagnostic accuracy (%) across conditions: initial sequential assessment prior to viewing any LLM output (Seq Init), final sequential assessment after viewing LUNAR response with the opportunity to revise (Seq Final), and con-current assessment where the case was presented alongside LUNAR response (Conc). Baseline accuracies were 35% (Q3B) and 34% (Q7B), whereas LUNAR achieved an accuracy of 52%. Dashed lines indicate LUNAR, Q3B, and Q7B performance on the same cases. (b) Inter-rater agreement (Fleiss’ kappa) for the same three conditions, with 95% bootstrap confidence intervals. (c) Response time (seconds) by condition, after removing outliers (values *>* 1.5 x IQR); shown as boxplots indicating median, interquartile range and whiskers (min/max). In panels (a)-(c), pairwise significance brackets are from case-level permutation tests, and in panels (a) and (b), bars represent mean accuracy with 95% bootstrap confidence intervals case-level resampling); non-significant comparisons are not shown. Asterisks denote: * *p <* 0.05, ** *p <* 0.01, *** *p <* 0.001, **** *p* < 0.0001.

### Synergistic benefits of reinforcement learning

Reinforcement learning (RL) produced improvements in reward optimization, calibration, conciseness and generalization relative to supervised fine-tuning (SFT) and ablated variants (Fig. 2). All configurations showed increasing mean group rewards over training, but the trajectories diverged markedly by ablation (Fig. 2a, top). Models without oversampling (LUNAR-OS-SCe, LUNAR-OS) achieved the fastest early reward gains, consistent with training on an easier distribution that under-represented rare and difficult cases. In contrast, models trained using OS data (LUNAR-SCe, LUNAR) displayed slower initial reward growth due to increased difficulty introduced by reweighting rare examples, yet converged more steadily with progressively lower reward variance. This pattern indicates that OS enhances training stability and robustness by preventing premature convergence on common classes.

Mean token entropy trajectories revealed complementary roles of OS and self-certainty (SCe) in shaping calibration (Fig. 2a, bottom-left). Removing SCe (LUNAR-OS-SCe, LUNAR-SCe) resulted in persistently higher entropy, indicating less confident policies. Incorporating SCe reduced entropy throughout training, consistent with increased self-certainty. However, in the absence of OS (LUNAR-OS), entropy collapsed rapidly, suggesting overly confident and potentially brittle policy updates. In contrast, the full model (LUNAR) exhibited a gradual and stable entropy decrease, indicating confidence increased progressively over training rather than rising too quickly early on. Mean response length for the models trained with SCe decreased most sharply in the absence of OS (LUNAR-OS), aligning with rapid entropy collapse and aggressive reward maximization (Fig. 2a, bottom-right). The full model maintained moderate response compression, balancing conciseness with stability. Together, these results suggest that OS and SCe play complementary roles: OS makes training more challenging and prevents the model from becoming overconfident too quickly, while SCe increases model confidence over time. Combined, they lead to slower but more stable learning. Mean token entropy on held-out testing data corroborated the training dynamics observed in Fig. 2a (Fig. 2b). Comparing the distribution of response-level mean Shannon entropy (computed across generated tokens and approximated using the top-20 tokens per position) across a sample of testing data revealed that models trained with SCe, including the full LUNAR model, exhibited significantly lower median entropy relative to their non-SCe counterparts. This is consistent with the entropy reduction observed during training, and confirms that the effect of SCe on model confidence generalizes beyond the training distribution to unseen cohorts. Notably, Q7B exhibited the lowest entropy overall, suggesting that the larger baseline model is inherently more confident in its predictions, whereas the models trained with SCe approached comparable confidence levels despite having roughly half the parameters.

RL altered output length distributions relative to the baseline models (Fig. 2c). The reductions in median output length were observed in models trained with SCe, including the full LUNAR model, whereas variants without SCe generated longer responses. This pattern is consistent with the entropy dynamics in Figs. 2a, 2b, where SCe lowered entropy and promoted more confident, concise generations. Macro-averaged accuracies (averaged across all ADRD cohorts and tasks) improved steadily during RL for both internal (NACC) and external cohorts (Fig. 2d, right), with external validation showing a clear upward trend across training steps. In contrast, SFT (Fig. 2d, left) improved performance mainly on the internal validation set, while performance on external cohorts remained largely flat. Notably, RL (LUNAR and all ablated variants; fig. S10) surpassed the fixed baseline performance (Q3B and Q7B) on both internal and external cohorts and across nearly all evaluation tasks (fig. S11), suggesting improved generalization rather than cohort-specific over-fitting. RL-trained models maintained or slightly improved the macro-averaged accuracy across five standard medical benchmarks (MedQA, MedMCQA, MedExpQA, MMLU–Clinical Knowledge and MMLU–Anatomy) compared to its baseline (Q3B), whereas SFT reduced the performance (0.54 to 0.51) (Fig. 2e). These results indicate that the gains achieved through RL training do not come at the expense of performance on established medical QA benchmarks.

### Performance on cognitive status and etiological diagnosis

LUNAR consistently outperformed the baseline models (Q3B and Q7B) across cognitive and etiologic classification tasks, with statistically robust improvements in most comparisons after FDR correction. For cognitive status classification (COG: normal cognition [NC], mild cognitive impairment [MCI], dementia [DE]), LUNAR achieved higher class-wise F1 scores than both baselines on internal testing (NACC) and external cohorts (Fig. 3a). On NACC, LUNAR F1 medians were 0.87 (NC), 0.53 (MCI), and 0.88 (DE), significantly exceeding Q3B (0.78, 0.40, 0.71) and Q7B (0.85, 0.45, 0.77) in nearly all classes (\*\*\*\**p <* 0.0001 for most pairwise tests). External cohort performance followed a similar pattern, with LUNAR showing gains in all categories (e.g., significant improvements over Q3B in most external sets; \**p <* 0.05 to \*\*\*\**p <* 0.0001), though some ns comparisons appeared in smaller or heterogeneous subsets. Bar plots showing comparisons of precision and recall values are provided in the supplement (fig. S12), and further model-specific performance metrics are detailed in table S9. Macro F1 scores for etiology assessment (ETPR) across all five cohorts are summarized in the radial plot (Fig. 3b). LUNAR attained the highest macro F1 medians in four of five cohorts (e.g., 0.32 in NACC, 0.57 in NIFD, 0.73 in ADNI, 0.40 in BrainLat), surpassing Q3B and Q7B in multiple instances (\*\*\*\**p <* 0.0001 in several cohort-model pairs). The model showed particularly strong performance in cohorts with richer multimodal data (e.g., ADNI and BrainLat), while differences were smaller or ns in others (e.g., PPMI). Circular bar plots showing comparisons of precision and recall values are provided in the supplement (fig. S13), and further model-specific performance metrics are detailed in tables S10 - S15.

### Alignment with biomarker status and postmortem evidence

Biomarker positivity classification tasks yielded clear advantages for LUNAR (Fig. 3c). In amyloid PET classification, LUNAR macro F1 medians were superior on both NACC (0.74 vs. 0.63 Q3B and 0.65 Q7B; \*\*\**p <* 0.0001) and external validation (0.67 vs. 0.63 Q3B and 0.61 Q7B; \*\*\*\**p <* 0.0001). Similar patterns held for amyloid CSF (NACC: 0.70 vs. 0.58 and 0.61; \*\*\*\**p <* 0.0001 and \*\*\**p <* 0.001, external: 0.65 vs. 0.63 and 0.56; \*\*\*\**p <* 0.0001 vs. Q7B) and DaT imaging (external: 0.39 vs. 0.31 and 0.25; \*\*\*\**p <* 0.0001), with most pairwise differences significant after FDR adjustment. In the neuropathology one-class task (NP-ONE: None, AD, LBD, FTLD), LUNAR demonstrated improved class-wise F1 medians relative to baselines (Fig. 3d). Notable gains occurred in None (0.29 vs. 0.11 Q3B and 0.30 Q7B; \*\**p <* 0.01 vs. Q3B) and LBD (0.40 vs. 0.22 and 0.32; \**p < 0.05* vs. Q3B), while differences were ns in some classes (e.g., AD and FTLD). Dot and line plots showing comparisons of precision and recall values are provided in the supplement (figs. S14, S15), and further model-specific performance metrics are detailed in tables **??** - S19. Across all panels, medians and 95% bootstrap confidence intervals (1,000 resamples) supported the reliability of these differences, with participant-level non–parametric permutation tests (10,000 iterations) and Benjamini-Hochberg FDR correction applied appropriately within cohorts, classes, and metrics. These results highlight LUNAR’s superior discriminative ability in differential diagnosis, particularly for challenging MCI, mixed etiologies, and biomarker-defined subgroups.

### Neurologist-level validation

In a balanced, within-subjects experiment involving 12 board-certified neurologists and 100 randomly selected cases from the NACC testing set (figs. S16-S19), exposure to LUNAR responses improved diagnostic performance. Mean diagnostic accuracy was lowest in the initial sequential arm assessments (prior to LLM exposure) at 44.3%. After reviewing LUNAR outputs, final sequential arm accuracy rose to 48.4% (*p <* 0.01), while concurrent arm accuracy (LLM response presented alongside the case from the start) reached 47.2%. The concurrent arm did not differ significantly from either sequential initial (*p* = 0.17) or final accuracy *(p = 0.55),* in unpaired comparisons (Fig. 4a). In comparison, baseline accuracies were 35% for Q3B and 34% for Q7B, while LUNAR achieved 52%. Inter-rater agreement, measured by Fleiss’ kappa, improved with LUNAR assistance. Baseline agreement in the initial sequential arm was *k* = 0.45 (95% CI: 0.38–0.52), with significant increases to *k* = 0.56 (95% CI: 0.49–0.63, *p* = 0.01) in the final sequential arm and to *k* = 0.57 (95% CI: 0.49–0.63, *p <* 0.0001) in the concurrent arm (fig. 4b). Agreement did not differ between the two AI-assisted conditions (*p* = 0.75).

Response times reflected the review process. Median time for the full sequential arm (initial + post-LUNAR review) was 161.0s, significantly longer than both the initial sequential assessment alone (median = 114.6s, *p <* 0.0001) and the concurrent arm (median = 124.5s, *p <* 0.0001) (Fig. 4c). Initial sequential and concurrent times did not differ significantly (*p* = 0.13), indicating that simultaneous presentation of LUNAR outputs may not have imposed the additional temporal burden associated with sequential review. Among *n* = 578 sequential-arm instances with paired pre- and post-LUNAR diagnoses, neurologists revised their answer in 88 cases (15.2%) (fig. S21). Revisions were net beneficial: 37 initially incorrect diagnoses were revised to correct after LUNAR review, compared with only 14 initially correct diagnoses that were changed to incorrect (odds ratio = 2.64, McNemar’s test *χ^2^* = 9.49, *p* = 0.002) (Fig. S22). Direct head-to-head comparisons of diagnostic quality (5-point Likert table S20) across five dimensions showed that neurologists rated LUNAR responses as significantly more concise than those from the baseline Q3B model (*p <* 0.001). No significant differences were observed in primary etiology identification, biomarker evidence usage, clinical-biomarker linkage or differential diagnosis appropriateness, indicating LUNAR maintained comparable reasoning quality while achieving greater clarity and brevity (Fig. S20).

## Discussion

In this work, we introduce LUNAR, a generative language model designed to support diagnostic reasoning in Alzheimer’s and related dementias (ADRD) across a range of clinical data availability. We established a standardized processing pipeline that harmonizes heterogeneous multimodal data from multiple cohorts into structured, narrative summaries suitable for language model processing. Building on this foundation, we fine-tuned a compact 3-billion-parameter model using reinforcement learning with verifiable rewards (RLVR) on a large NACC dataset, incorporating task-specific oversampling of rare etiologies and self-certainty regularization to enhance calibration and interpretability. The resulting system was evaluated comprehensively on held-out NACC data and four external cohorts (ADNI, BrainLat, NIFD and PPMI), spanning syndromic classification, primary etiological diagnosis, biomarker alignment (A*β* PET, CSF A*β*, and DaT imaging), and structured reasoning in complex mixed dementia cases. Where possible, predictions were validated against neuropathological findings, and clinical utility was assessed through blinded expert neurologist review comparing model outputs with reference summaries.

Our approach offers some practical advantages. RLVR removes the need for curated chain-of-thought annotations, which are costly and difficult to obtain at scale, especially for clinical tasks. Instead, supervision relies only on final answers, enabling more scalable training while still providing informative learning signals. In addition, oversampling improves coverage of underrepresented etiologies in the training data, helping mitigate class imbalance and improving robustness across diagnostic categories. Finally, the self-certainty-aware advantage encourages lower predictive entropy, leading to more concise and confident responses, which is particularly important in medical settings, where reliable and decisive outputs are critical for clinical decision support. As such, LUNAR is deliberately engineered as a compact, domain-adapted specialist rather than a broad generalist. This targeted design prioritizes efficiency, calibration and clinical fidelity over raw parameter scale, while enabling accessible, open-source LLM development for medical applications. Consequently, direct comparisons with frontier general-purpose LLMs offer limited insight into LUNAR’s intended strengths: clinical performance on heterogeneous ADRD presentations, generalizability across diverse cohorts and resource settings, and feasibility for secure, low-resource deployment. Importantly, the resulting 3-billion-parameter model is lightweight enough to support local or edge-computing deployment in under-resourced environments (e.g., community clinics or rural hospitals), providing a pathway to low-latency, privacy-preserving AI assistance without dependence on cloud infrastructure or proprietary APIs.

In many clinical contexts, patients undergo comprehensive neuroimaging and biomarker evaluations, while others have only limited cognitive testing or basic medical histories. AI models that require fully standardized multimodal inputs are therefore poorly suited to such heterogeneous environments. By contrast, our approach processes each case through modality-specific pipelines, aggregating structured summaries into a unified, individual-level representation. This design enables the model to learn from information that is available and still provide clinically meaningful insights, even when certain modalities (e.g., imaging or biomarker data) are unavailable. We trained the model on datasets encompassing multiple combinations of input modalities to enhance generalizability across diverse practice settings, including those reflected in the testing cohorts. This workflow mirrors real-world practice, where clinicians synthesize available information and determine whether additional evaluations, such as biomarker testing or neuroimaging, are warranted. At inference, the model adaptively integrates available data sources (e.g., neuroimaging, cognitive assessments) without requiring complete datasets, dynamically adjusting its reasoning to account for missing modalities. This adaptability aligns with how ADRD evaluations are typically conducted, given variability in diagnostic resources due to institutional constraints, patient preferences, or healthcare system factors. Consequently, the model is well suited for diverse clinical settings, including memory clinics, general neurology, primary care, and family medicine, where it can support differential diagnosis, guide management decisions, and inform the need for additional testing.

Our study has a few limitations. First, while the model was trained and evaluated on a large set of multimodal cohorts, data heterogeneity across cohorts can introduce variability in imaging protocols and clinical assessments, which may affect generalizability. To partially mitigate this, we implemented automated processing pipelines to derive individual-level summaries and ensured that training incorporated diverse data distributions to enhance robustness. Second, although we employed reinforcement learning using diagnostic summaries, the dataset remained limited in size, potentially constraining the model’s ability to capture the full spectrum of ADRD presentations. Third, the model was primarily evaluated using retrospective datasets, limiting its validation in clinical settings. To approximate real-world applicability, we conducted a blinded assessment in which expert clinicians assessed model outputs for accuracy, reasoning and clinical validity. Fourth, while model-generated impressions were validated against postmortem neuropathology findings, the number of neuropathology-confirmed cases was limited. Finally, while our framework successfully enables multimodal learning by using modality-specific pipelines to integrate diverse data sources, the effectiveness of feature extraction and the integration of modality-specific information into model training can be further improved. Enhancing feature extraction techniques and the way modality-specific data is embedded during model training could lead to more useful summaries. Our current framework is designed for seamless extension, allowing future refinements in both modality-specific pipelines and multimodal representation learning to further improve its utility. Additionally, while our initial focus was on standard neurology work-up data, such as neuroimaging and clinical assessments, future extensions could integrate emerging modalities, including voice and wearable sensor data, further enriching the model’s diagnostic capabilities. Despite these limitations, our work provides proof-of-concept for a foundation model capable of integrating and reasoning across multiple medical data types.

In conclusion, our study presents LUNAR as an effective domain-adapted language model capable of synthesizing heterogeneous clinical data into coherent, structured diagnostic summaries for ADRD. Through rigorous evaluation, including question-answering tasks, biomarker prediction, alignment with neuropathological findings and blinded clinician assessment, LUNAR demonstrates robust diagnostic reasoning and clinical relevance. Although prospective validation in real-world settings remains essential, these results highlight the value of targeted fine-tuning with reinforcement learning to enable compact language models to support AI-assisted decision support across complex domains in medicine.

## Data Availability

Data from ADNI, NIFD and PPMI are available to download from the Laboratory of Neuro Imaging website at https://ida.loni.usc.edu. NACC data can be requested and downloaded at https://naccdata.org. Instructions to download BrainLat data are provided at https://doi.org/10.1038/s41597-023-02806-8.

## Acknowledgments

None.

## Funding

This project was supported by grants from the National Institute on Aging’s Artificial Intelligence and Technology Collaboratories (P30-AG073104 & P30-AG073105), and the National Institutes of Health (R01-HL159620, R01-AG062109, R01-NS142076 and R01-AG083735). We acknowledge the efforts of several individuals from the ADNI, BrainLat, NACC, NIFD and PPMI for providing access to data.

This work used Delta GPUs at NCSA through allocation CIS250628 from the Advanced Cyberinfrastructure Coordination Ecosystem: Services & Support (ACCESS) program (*31*), which is supported by U.S. National Science Foundation grants #2138259, #2138286, #2138307, #2137603, and #2138296.

Data collection and sharing for this project was funded by the Alzheimer’s Disease Neuroimaging Initiative (ADNI) (National Institutes of Health Grant U01 AG024904) and DOD ADNI (Department of Defense award number W81XWH-12-2-0012). ADNI is funded by the National Institute on Aging, the National Institute of Biomedical Imaging and Bioengineering, and through generous contributions from the following: AbbVie, Alzheimer’s Association; Alzheimer’s Drug Discovery Foundation; Araclon Biotech; BioClinica, Inc.; Biogen; Bristol-Myers Squibb Company; CereSpir, Inc.; Cogstate; Eisai Inc.; Elan Pharmaceuticals, Inc.; Eli Lilly and Company; EuroImmun; F. Hoffmann-La Roche Ltd and its affiliated company Genentech, Inc.; Fujirebio; GE Healthcare; IXICO Ltd.; Janssen Alzheimer Immunotherapy Research & Development, LLC.; Johnson & Johnson Pharmaceutical Research & Development LLC.; Lumosity; Lundbeck; Merck & Co., Inc.; Meso Scale Diagnostics, LLC.; NeuroRx Research; Neurotrack Technologies; Novartis Pharmaceuticals Corporation; Pfizer Inc.; Piramal Imaging; Servier; Takeda Pharmaceutical Company; and Transition Therapeutics. The Canadian Institutes of Health Research is providing funds to support ADNI clinical sites in Canada. Private sector contributions are facilitated by the Foundation for the National Institutes of Health (www.fnih.org). The grantee organization is the Northern California Institute for Research and Education, and the study is coordinated by the Alzheimer’s Therapeutic Research Institute at the University of Southern California. ADNI data are disseminated by the Laboratory for Neuro Imaging at the University of Southern California.

Information on BrainLat is available here: https://doi.org/10.1038/s41597-023-02806-8.

The NACC database is funded by an NIA grant U24-AG072122. NACC data are contributed by the NIA-funded ADRCs: P30 AG062429 (PI James Brewer, MD, PhD), P30 AG066468 (PI Oscar Lopez, MD), P30 AG062421 (PI Bradley Hyman, MD, PhD), P30 AG066509 (PI Thomas Grabowski, MD), P30 AG066514 (PI Mary Sano, PhD), P30 AG066530 (PI Helena Chui, MD), P30 AG066507 (PI Marilyn Albert, PhD), P30 AG066444 (PI John Morris, MD), P30 AG066518 (PI Jeffrey Kaye, MD), P30 AG066512 (PI Thomas Wisniewski, MD), P30 AG066462 (PI Scott Small, MD), P30 AG072979 (PI David Wolk, MD), P30 AG072972 (PI Charles DeCarli, MD), P30 AG072976 (PI Andrew Saykin, PsyD), P30 AG072975 (PI David Bennett, MD), P30 AG072978 (PI Neil Kowall, MD), P30 AG072977 (PI Robert Vassar, PhD), P30 AG066519 (PI Frank LaFerla, PhD), P30 AG062677 (PI Ronald Petersen, MD, PhD), P30 AG079280 (PI Eric Reiman, MD), P30 AG062422 (PI Gil Rabinovici, MD), P30 AG066511 (PI Allan Levey, MD, PhD), P30 AG072946 (PI Linda Van Eldik, PhD), P30 AG062715 (PI Sanjay Asthana, MD, FRCP), P30 AG072973 (PI Russell Swerdlow, MD), P30 AG066506 (PI Todd Golde, MD, PhD), P30 AG066508 (PI Stephen Strittmatter, MD, PhD), P30 AG066515 (PI Victor Henderson, MD, MS), P30 AG072947 (PI Suzanne Craft, PhD), P30 AG072931 (PI Henry Paulson, MD, PhD), P30 AG066546 (PI Sudha Seshadri, MD), P20 AG068024 (PI Erik Roberson, MD, PhD), P20 AG068053 (PI Justin Miller, PhD), P20 AG068077 (PI Gary Rosenberg, MD), P20 AG068082 (PI Angela Jefferson, PhD), P30 AG072958 (PI Heather Whitson, MD), P30 AG072959 (PI James Leverenz, MD).

Data collection and sharing for this project was funded by the Frontotemporal Lobar Degeneration Neuroimaging Initiative (National Institutes of Health Grant R01-AG032306). The study is coordinated through the University of California, San Francisco, Memory and Aging Center. FTLDNI data are disseminated by the Laboratory for Neuro Imaging at the University of Southern California.

Data used in the preparation of this article was obtained on 2024-07-29 from the Parkinson’s Progression Markers Initiative (PPMI) database (www.ppmi-info.org/access-data-specimens/ download-data), RRID:SCR_006431. For up-to-date information on the study, visit www.ppmi-info. org. PPMI – a public-private partnership – is funded by the Michael J. Fox Foundation for Parkinson’s Research, and funding partners.

## Data and materials availability

Python scripts, model checkpoints, help files and information on the study population will be made available on GitHub upon acceptance of the manuscript.

## Supplementary material

Methods

Supplementary text Figs. S1 to S22

Tables S1 to S20

References (*7–50*)

