## Supplement for "Domain-adapted language model using reinforcement learning for various dementias"

Kowshik et. al.,

**This PDF file includes:**

Methods

Supplementary text

Figures S1 to S22

Tables S1 to S20

### Methods

All data were obtained in de-identified form from the respective study centers, with appropriate ethical oversight for each cohort. Study procedures and secondary data usage were approved by the institutional review boards or ethics committees responsible for the original studies, and written informed consent had been obtained from all participants or their legally authorized representatives prior to data collection. Specifically, the Alzheimer's Disease Neuroimaging Initiative (ADNI; `https://adni.loni.usc.edu/`) and the Parkinson's Progression Markers Initiative (PPMI; `https://www.ppmi-info.org/`) were approved by institutional review boards at each participating site. Data from the National Alzheimer's Coordinating Center (NACC; `https://naccdata.org/`) were collected under protocols approved by institutional review boards at each contributing Alzheimer's Disease Research Center. The Frontotemporal Lobar Degeneration Neuroimaging Initiative (NIFD/FTLDNI; `https://memory.ucsf.edu/research-trials/research/allftd`) received approval from the institutional review board at each participant site. For the BrainLat project, ethical approval for data collection and sharing was provided by the institutional ethics boards at each recruitment site (detailed in the Ethics section of Prado et al. (*32*)).

**Study population.** We aggregated comprehensive multimodal data from five independent cohorts, comprising a total of $54,535$ participants (table S1). This included demographic information, personal and family medical history, laboratory results, physical and neurological examination findings, medication records, neuropsychological test scores, and functional assessments. Also, when available, we included multi-sequence magnetic resonance imaging (MRI), dopamine transporter (DaT) imaging, amyloid positron emission tomography (PET), and neuropathological data. The cohorts were the National Alzheimer's Coordinating Center (NACC) ($n = 48,876$) (*33*), the Alzheimer's Disease Neuroimaging Initiative (ADNI) ($n = 1,392$) (*34*), the frontotemporal lobar degeneration neuroimaging initiative (NIFD) ($n = 292$) (*35*), the Parkinson's Progression Marker Initiative (PPMI) ($n = 3,315$) (*36*), and BrainLat ($n = 660$) (*32*). This resulted in $22,149$ participants with normal cognition (NC), $10,421$ participants with mild cognitive impairment (MCI), and $21,965$ participants with dementia.

The primary etiologies of cognitive impairment in the study population were derived from the clinical diagnoses provided by each contributing cohort and were distributed as follows: $21,094$

participants had Alzheimer's disease (AD); $2,399$ had Lewy body disease and/or Parkinson's disease (LBD); $1,331$ had vascular dementia or cognitive impairment resulting from vascular brain injury, including stroke (VD); $3,302$ had frontotemporal lobar degeneration spectrum disorders (FTLD), encompassing its clinical variants, corticobasal degeneration, progressive supranuclear palsy, and cases with or without concomitant amyotrophic lateral sclerosis; 864 had cognitive impairment due to systemic and external factors (SEF), including infections, metabolic disturbances, substance abuse (alcohol or medications), delirium and systemic illnesses; 550 presented with cognitive impairment primarily attributed to psychiatric disorders (PSY) such as schizophrenia, major depression, bipolar disorder, anxiety disorders, and post-traumatic stress disorder; and 359 had cognitive impairment arising from other etiologies (OTH), comprising neoplasms, normal pressure hydrocephalus, prion diseases (including Creutzfeldt-Jakob disease), multiple system atrophy, traumatic brain injury, essential tremor, Huntington's disease, Down syndrome, and epilepsy or seizure disorders.

**Selection criterion.** Participants were eligible for inclusion if they carried a diagnosis of NC, MCI or dementia. The cases from the NACC cohort were split into a training set ($n = 17,847$) and an internal test set ($n = 31,029$). The remaining four cohorts (ADNI, NIFD, PPMI and BrainLat) were reserved exclusively for external, out-of-distribution validation. Cohort-specific selection criteria were applied as follows. In NACC and PPMI, only the most recent visit per participant was included. In NIFD, only the final visit was included, restricted to individuals with NC or a diagnosis of a subtype of frontotemporal dementia (FTD): behavioral variant (bvFTD), semantic variant (svFTD), and progressive non-fluent aphasia (PNFA). For ADNI, participants were required to have an A$\beta$ PET scan and an MRI acquired within a 12-month window. From the BrainLat cohort, we initially selected all available cases, then excluded those with Parkinson's disease (no cognitive testing performed) and multiple sclerosis (outside study scope), and removed two additional participants with duplicate non-unique identifiers, yielding a final sample of $n = 660$ with diagnoses of NC, bvFTD or AD. No participants from any cohort were excluded due to missing information.

**Data processing.** To enable unified processing of heterogeneous multimodal data across cohorts, we implemented a standardized processing pipeline that converted all tabular clinical, laboratory, neuropsychological and demographic data into structured, human-readable text embedded within JSON files. Each cohort was processed according to its respective data dictionary, which guided the mapping of original variable names, coded values and units to consistent natural language descriptions. All references to the missing values were omitted from the textual representation, ensuring that only observed information was presented to the model. No participants were excluded due to incomplete records or absent diagnostic labels.

The resulting JSON structure for each participant was divided into two principal sections: (1) `patient_summary`, containing objective raw participant data organized by modality (e.g., demographics, medical history, physical and neurological examination findings, laboratory results, neuroimaging findings, medication lists, neuropsychological test scores, and functional assessments); and (2) `diag_summary`, reserved for clinician-derived interpretations, diagnostic impressions, and formal cognitive syndrome or etiological diagnoses. This separation allowed flexible control during training and inference, enabling experiments that either withheld or included prior clinical interpretations.

The NACC cohort provides radiological findings from MRI and PET scans, which we incorporated directly into the `patient_summary`. In contrast, the ADNI, PPMI, NIFD, and BrainLat cohorts do not supply radiology reports for MRI data, so we processed their imaging files to produce the corresponding textual descriptions. All available T1-weighted MRI scans were processed using FastSurfer (*37*) to obtain whole-brain segmentation and parcellation. Regional cortical and subcortical volumes were extracted and aggregated into seven bilateral regions of interest relevant to neurodegenerative disease: hippocampus, medial temporal lobe, lateral temporal lobe, medial parietal lobe, lateral parietal lobe, frontal lobe, and occipital lobe. Volumes from corresponding left- and right-hemisphere subregions were summed. To derive biologically meaningful normative references and detect atrophy, we fitted a generalized additive model for location, scale, and shape (GAMLSS) with a generalized gamma distribution to MRI data from cognitively normal individuals across all testing cohorts ($n = 2,087$), adjusting for age, sex, and cohort membership following established methodologies (*38*). Age- and sex-specific centile curves were generated using 1000 bootstrap resamples. Regional volumes were then expressed as centile scores relative to this normative reference. Atrophy severity was classified as mild (5th–10th centile), moderate (1st–5th centile), or severe (<1st centile).

All MRI-derived volumetric features and corresponding normative interpretations were appended as additional structured text entries within the `patient_summary` section of each participant's JSON file. Because the current model is purely language-based and we lacked sufficient high-quality image–report pairs for vision-language pretraining, no direct visual processing of MRI scans was performed; all imaging information was represented exclusively in textual/numeric form, as described above. An illustrative JSON schema is provided in Supplementary Material (figs. S6, S7).

**Person-level case narrative generation.** After all clinical, laboratory, neuropsychological, and imaging-derived data for each participant had been consolidated into a standardized JSON file, we converted these records into concise, clinically coherent prose narratives using the Qwen3-32B language model in its "non-thinking" mode (*39*). This mode explicitly disables chain-of-thought reasoning to maximize fidelity to the source material, which is optimal for summarization tasks requiring adherence to provided evidence. The narrative generation step served two essential purposes. First, it produced inputs that resemble real-world clinical case presentations, enabling the model to learn reasoning patterns. Second, it reduced input length and improved the signal-to-noise ratio, which is an important factor for stable large language model performance. Across cohorts, the median token count decreased from $3,339$ (95% CI: $928 - 5,556$) in raw JSON format to 824 tokens (95% CI: $460 - 1,156$) after summarization, corresponding to a median 75% reduction (95% CI: $28 - 86$%). The prompt template and representative before-and-after examples are provided in Supplementary Material (figs. S2, S6, S7, & S8).

**Question generation and task formulation.** LUNAR was trained using reinforcement learning with verifiable rewards (RLVR) (detailed below). To support this strategy, all tasks were formulated as closed-ended multiple-choice questions with a single correct answer to fully automate the scoring of model responses without human intervention or reward models.

We defined the following clinically relevant question types: current cognitive status including normal cognition, mild cognitive impairment, or dementia (COG); primary etiological diagnosis of cognitive impairment (ETPR); A$\beta$ positivity by PET imaging (PET) or cerebrospinal fluid analysis (CSF); presence of Lewy body pathology according to dopamine transporter imaging

(DAT); neuropathologically confirmed single predominant pathology limited to NP-AD, NP-LBD, or NP-FTLD (NP-ONE); and neuropathologically confirmed mixed pathology comprising any combination thereof (NP-MIXED).

Within the NACC training partition, each participant was randomly assigned to one of the seven question types, ensuring that every case contributed exactly one training example. However, during internal and external validation across all cohorts, every applicable question type was posed for each participant. To further enhance linguistic variation and prevent models from developing a preference for any specific label, five distinct paraphrases were generated for each question type, and the answer options were randomly shuffled on every presentation during training and validation. Full descriptions of question types, answer choices, and example prompts are provided in Supplementary Material (tables S2 - S8, figs. S3 & S4).

**Reinforcement learning with oversampling and self-certainty-aware advantage.** We framed our task as a multiple-choice question-answering problem with a single correct answer and trained the language model using RLVR. Each training instance consisted of a person-level narrative, one of the six diagnostic question types (randomly assigned), and randomly shuffled answer choices. Formally, let the dataset $P_Q$ consist of tuples $(q, y^*)$, where $q$ denotes the query comprising the person's background, a diagnostic question drawn from the previously defined set $Q$, and a set of answer choices $\{a_1, a_2, \ldots, a_n\}$ that are randomly shuffled for each instance. The corresponding $y^*$ represents the reference label derived from the original cohort annotations (clinical consensus, imaging biomarkers, or neuropathology as appropriate). Given a query $q$, the model $\pi_\theta$ generates an output $o$ containing a chain of thought $c$ and a final predicted answer $\hat{y}$.

We trained the model to maximize the expected reward for generating the correct answer. The overall optimization objective is defined as:

$$\mathcal{J}(\theta) = \mathbb{E}_{q \sim P_Q,\, o \sim \pi_\theta(\cdot|q)} \left[ R(q, o) \right], \tag{S1}$$

where $R(q, o)$ is a binary reward function given by

$$R(q, o) = \begin{cases} 1, & \text{if the model generates } \hat{y} \text{ within } \texttt{\textbackslash boxed\{\}} \text{ and } \hat{y} \text{ (in the final boxed expression) } = y^*, \\ 0, & \text{otherwise.} \end{cases} \tag{S2}$$

To optimize this objective, we used the Done Right Group Relative Policy Optimization (Dr.GRPO) (*40*) algorithm, with an added per-token KL-penalty between the current policy $\pi_\theta$ and a reference policy $\pi_{\text{ref}}$, and a self-certainty-aware advantage function.

We proposed a self-certainty-aware advantage by adopting the self-certainty metric (SCe) (*41, 42*) defined as the average KL divergence between a uniform distribution U over the vocabulary V and the model's next-token distribution. For each response $o_i$ within a sampled group, we computed two distinct advantage estimates. First, we calculated the group-centered reward advantage as

$$\hat{A}^{\text{R}}_{i,t} = R(q, o_i) - \overline{R}, \tag{S3}$$

where $R(q, o_i)$ denotes the reward assigned to response $o_i$ given query $q$, and $\overline{R}$ represents the mean reward across the group. Second, we added a self-certainty-based advantage

$$\hat{A}^{\text{SCe}}_{i,t} = \frac{u_i - \overline{u}}{\sigma_U}, \tag{S4}$$

where $u_i$ = self-certainty$(o_i \mid q)$ quantifies the model's internal confidence for response $o_i$ conditioned on query $q$, $\overline{u}$ denotes the mean self-certainty across the group, and $\sigma_U$ represents the standard deviation of self-certainty scores. Without this term, more than 50% of prompts produced zero advantage at later stages of training, resulting in no gradient updates. Incorporating $\hat{A}^{\text{SCe}}_{i,t}$ provides a small directional optimization signal even in these cases, enabling continued parameter updates and more stable optimization. The composite self-certainty-aware advantage function is subsequently defined as

$$\hat{A}^{\text{comp}}_{i,t} = (1 - \lambda_{\text{SCe}})\hat{A}^{\text{R}}_{i,t} + \lambda_{\text{SCe}}\ s_i\ \tanh(\hat{A}^{\text{SCe}}_{i,t}), \tag{S5}$$

where $s_i = 2R(q, o_i) - 1 \in \{-1, +1\}$ preserves reward polarity, and $\lambda_{\text{SCe}} \in [0, 1]$ controls the relative contribution of the self-certainty advantage $\hat{A}^{\text{SCe}}_{i,t}$ with respect to the reward-based advantage $\hat{A}^{\text{R}}_{i,t}$. This mechanism explicitly rewards high confidence when correct and penalizes high confidence when incorrect. The tanh function is applied to $\hat{A}^{\text{SCe}}_{i,t}$ to bound its scale within $(-1, 1)$, ensuring consistency with the range of the reward-based advantage. Therefore, we maximized the following clipped surrogate objective:

$$\begin{aligned}\mathcal{J}_{\text{LUNAR}}(\theta) = {} & \mathbb{E}_{q\sim P(Q),\ \{o_i\}_{i=1}^{G}\sim\pi_{\theta_{\text{old}}}(O|q)} \\ & \frac{1}{G}\sum_{i=1}^{G}\sum_{t=1}^{|o_i|} \min\left(r_{i,t}\hat{A}^{\text{comp}}_{i,t},\ \text{clip}\left(r_{i,t},\ 1-\epsilon,\ 1+\epsilon\right)\hat{A}^{\text{comp}}_{i,t}\right) - \beta\mathbb{D}_{\text{KL}}\left[\pi_\theta \,\|\, \pi_{\text{ref}}\right]\end{aligned}, \tag{S6}$$

where $r_{i,t} = \frac{\pi_\theta(o_{i,t} \mid q, o_{i,<t})}{\pi_{\theta_{\text{old}}}(o_{i,t} \mid q, o_{i,<t})}$, $G$ is the group-size, and $\epsilon$ and $\beta$ are the hyperparameters.

To address severe class imbalance in the NACC training data, where AD dominated most question types, we performed task-specific oversampling with replacement. For each question type independently, we identified the majority-class size $N_{\text{max}}$ and repeatedly sampled minority classes until every label reached $N_{\text{max}}$ instances (the maximum unique count under each question type in table S2). This ensured equal exposure to rare etiologies without distorting underlying data characteristics and reduced the risk of model collapse toward the majority class prediction (the question variant the ordering of the options remained the same for the oversampled cases).

**Supervised fine-tuning.** To provide a direct comparison with our reinforcement learning framework, we trained a supervised fine-tuning (SFT) baseline using the identical training partition and question formulation. Each training instance consisted of a person-level narrative, one randomly assigned diagnostic question type (from the same seven used in RL training), and randomly shuffled answer choices. The target output was formatted to include a final answer enclosed within `\boxed{}`. More details are presented in fig. S4.

The SFT model was optimized via standard cross-entropy loss over all output tokens. Formally, given a query $q$ and reference output $o^*$, the objective is

$$\mathcal{L}_{\text{SFT}}(\theta) = -\mathbb{E}_{(q,o^*)\sim P_Q}\left[\sum_{t=1}^{|o^*|} \log \pi_\theta(o_t^* \mid q, o_{<t}^*)\right]. \tag{S7}$$

To maintain comparability with the RL setting, we applied the same task-specific oversampling strategy to mitigate class imbalance within each question type (table S2). Model selection was performed based on validation loss.

**Experimental setup.** LUNAR was trained using the Open-R1 framework (*43*), an open-source implementation of DeepSeek-R1 released by Hugging Face. We used Qwen2.5-3B-Instruct (Q3B) (*44*) as the backbone model. We applied a single gradient update per generation step, where each update processed 16 cases, with 16 candidate responses generated per case using vLLM (*45*). The generation temperature was set to 1.0, with top-$p$ sampling disabled. Training used a KL penalty of $\beta = 0.001$, a learning rate of $1.0 \times 10^{-6}$, $\lambda_{SCe} = 0.05$, and a maximum prompt and completion length of 2,000 tokens. The model was trained for 1,491 steps (1 epoch) using the

AdamW optimizer with 0.01 weight decay and a cosine learning-rate scheduler. We compare our model to Q3B (the backbone model for LUNAR) and the larger Qwen2.5-7B-Instruct (Q7B) models (*44*). We also evaluate LUNAR against its ablated variants: LUNAR–OS, trained without oversampled data; LUNAR–SCe, trained without the self-certainty–aware advantage (i.e., using only the group-centered reward advantage); and LUNAR–OS–SCe, trained without oversampled data and self-certainty–aware advantage.

We additionally trained a supervised fine-tuning (SFT) baseline using the same Qwen2.5-3B-Instruct (Q3B) backbone for comparison. Training was performed on the task-specific oversampled NACC partition described above. The model was optimized with AdamW at a learning rate of $1.0 \times 10^{-5}$ and weight decay of 0.01, employing a cosine learning-rate scheduler. The maximum sequence length was set to 4,000 tokens. Training proceeded for a maximum of 50 epochs with early stopping (patience = 3) based on validation loss. Early stopping was triggered at epoch 21, and the checkpoint with the lowest validation loss was restored at completion (epoch 18). All models were trained using DeepSpeed ZeRO-2 (*46*) to enable efficient distributed optimization.

For evaluations of RL-trained models, both the RL models and their baselines were prompted using chain-of-thought prompting, with the final answer enclosed in `\boxed{}` (fig. S3). In contrast, evaluations involving SFT-trained models prompted both the SFT models and their baselines to produce only the final answer, also enclosed in `\boxed{}` (fig. S4). To account for sampling stochasticity, each case was presented five independent times at temperature $T = 1.0$ without nucleus (top-$p$) sampling. For each case, if the model failed to generate an answer within `\boxed{}` or generates multiple such expressions, we used the Qwen3-14B model (non-thinking mode) (*39*) to extract the final answer (fig. S5).

**Evaluation on standard medical benchmarks** We evaluated our model on five standard medical question-answering benchmarks. MedQA (*47*) ($n$ = 1,255) consists of multiple-choice questions drawn from the United States Medical Licensing Examination (USMLE). MedMCQA (*48*) ($n$ = 4,183) is a large-scale dataset sourced from Indian medical entrance examinations (AIIMS and NEET PG), spanning over 2,400 healthcare topics across 21 medical subjects. MedExpQA (*49*) ($n$ = 121) is derived from the Spanish Medical Intern Resident (MIR) examination. From the Massive Multitask Language Understanding (MMLU) benchmark (*50*), we included two subsets targeting complementary aspects of medical knowledge: anatomy ($n$ = 135), which assesses foundational knowledge of human body structure, and clinical knowledge ($n$ = 265), which tests applied understanding of disease mechanisms, diagnosis, and patient management.

**Evaluation on ADRD benchmarks.** We evaluated the models on custom datasets constructed directly from the de-identified multimodal ADRD cohort data. Inclusion of standard benchmarks (e.g., MedQA) enabled direct comparison with general-purpose large language models, whereas the custom benchmarks provided a more faithful assessment of real-world diagnostic performance. Real-world clinical records are inherently noisy, frequently incomplete, and may contain irrelevant details without a single "textbook" answer, making our benchmarks substantially more representative of actual diagnostic challenges than highly curated examination-style questions. Moreover, using internal cohort cases eliminates the risk of test-set contamination or memorization that could artificially inflate scores on widely circulated public benchmarks whose training overlap with pretrained models is unknown.

Six clinically focused multiple-choice tasks were defined. The cognitive status task (COG) required classification into NC, MCI, or DE. For NACC, ADNI, BrainLat and NIFD cohorts, the primary etiology task (ETPR) required the selection of the most likely cause of cognitive impairment (if present) from eight mutually exclusive options: Alzheimer's disease, Lewy body disease, vascular dementia, frontotemporal lobar degeneration, psychiatric disorder, systemic/external factors, other specified etiology, or no impairment. For the PPMI cohort, the ETPR task required the selection of the most likely cause of cognitive impairment (if present) from four mutually exclusive options: Idiopathic Parkinson's disease, Prodromal Parkinson's disease, Other neurological disorder(s), or no Parkinson's disease nor other neurological disorder. The neuropathology task (NP-ONE) was restricted to autopsy-confirmed cases exhibiting a single clearly dominant pathology; the model predicted whether this was Alzheimer's disease, Lewy body disease or frontotemporal lobar degeneration. Finally, three independent biomarker-prediction tasks (PET, CSF and DAT) required binary prediction of A$\beta$ positivity by PET, A$\beta$ positivity in CSF, and presence of Lewy body pathology by DaT imaging, respectively. More details can be found in tables S3 - S8. Improvements for the neuropathologically confirmed mixed pathology (NP-MIXED) task were inconsistent across training steps (S11). Therefore, the NP-MIXED task was excluded from further analysis.

**Biomarker-level validation.** In the NACC and ADNI datasets, persons with PET imaging or CSF biomarker data available were used to evaluate the model's ability to identify amyloid-beta pathology. Similarly, in the NACC and PPMI datasets, persons with DaT imaging biomarker data available were used to evaluate the model's ability to identify Lewy body disease pathology. Classification performance was evaluated using macro-averaged precision, recall, and F1 score to account for class imbalance.

**Postmortem validation.** To further validate the model's ability to identify the presence of neuropathology, we evaluated its performance for the one-class (NP-ONE) task against post-mortem neuropathological assessments in individuals with autopsy-confirmed diagnoses. Testing samples were drawn exclusively from the NACC dataset. Ground-truth neuropathology labels were obtained directly from the dataset annotations and included no pathology (None), Alzheimer's disease (NP-AD), Lewy body disease (NP-LBD), and frontotemporal lobar degeneration (NP-FTLD). Model performance was assessed using class-wise precision, recall, and F1 score. For each neuropathology, precision quantified the proportion of model-predicted positive cases that were confirmed at post-mortem, whereas recall quantified the proportion of post-mortem-confirmed cases that were correctly identified by the model. F1 score was defined as the harmonic mean of precision and recall and was used to summarize the balance between the two.

**Neurologist-level validation.** Clinical utility of the model-generated diagnostic summaries was evaluated through a blinded, randomized crossover study by 12 fellowship-trained, board-certified neurologists. Each neurologist independently reviewed 100 de-identified cases selected from the NACC testing cohort. The 100 cases comprised 16 randomly selected individuals with normal cognition, 42 with mild cognitive impairment (MCI), and 42 with dementia. The MCI and dementia groups each included six randomly selected cases from the following etiological categories: AD, FTLD, LBD, PSY, SEF, VD, and Other. Cases were randomized at the within-participant level with approximately balanced allocation to one of two presentation formats: (1) sequential presentation, in which neurologists first answered using participant-level data alone and subsequently viewed the LUNAR response with the option to revise their answer (figs. S16 & S17), and (2) concurrent presentation, in which participant-level data and the LUNAR response were shown side-by-side prior to answer submission (fig. S18). In each case, neurologists were asked to answer the same multiple-choice question: `Determine the primary etiology underlying the person's cognitive impairment based on the provided information, if applicable.` After completing each case (in either arm), neurologists completed a side-by-side AI comparison task in which two AI responses were shown side-by-side, labeled only as `A (the response you previously viewed)` (LUNAR) and `B (a different response)` (Q3B) (fig. S19). Neurologists then answered the 5 questions spanning diagnostic accuracy, biomarker integration, clinical reasoning coherence, differential diagnosis appropriateness, and clinical utility on 5-point Likert scales (fig. S19, table S20). Of the 1, 200 case evaluations assigned, 1, 178 (98.2%) were completed. Twenty-two sequential-arm responses (1.8%) were not recorded due to unanticipated server downtime; these cases were excluded from sequential pre-post comparisons. All initial diagnoses were recorded. We then evaluated clinician performance under each condition as well as the time taken to reach a diagnosis. Given that variance decomposition revealed substantial case-level variation in accuracy ($\eta^2 = 0.59$, $p \leq 0.0001$) but negligible clinician-level variation ($\eta^2 = 0.01$, $p = 0.790$), we employed case-level permutation tests for all pairwise comparisons, treating cases as the unit of resampling to account for the dominant source of variability in clinician assessments.

**Performance metrics.** For all evaluations, each model was queried five times per patient (five independent trials), and all trials were pooled for metric computation. Model predictions were evaluated as classification tasks against ground truth labels. For the medical knowledge benchmarks, macro-averaged accuracy was used as the primary metric. For cognitive status and neuropathology tasks, class-level precision, recall, and F1-score were computed and reported per class. For the primary etiology and biomarker tasks, macro-averaged precision, recall, and F1-score were reported, weighting each class equally regardless of prevalence. Macro-averaged accuracy was additionally computed as a summary measure of overall performance across all classification tasks, and tracked across training steps during both supervised fine-tuning and reinforcement learning. Model uncertainty was quantified using mean token entropy, defined as the mean Shannon entropy of the model's next-token distribution across generated tokens, approximated using the top-20 tokens at each position.

**Statistical analysis.** We used a one-way ANOVA and the two-sided $\chi^2$ test for continuous and categorical variables, respectively, to assess overall differences in population characteristics across diagnostic groups within each study cohort. We then performed comprehensive statistical analyses to evaluate the effectiveness of our model's training strategy and compare its performance with various baseline models. First, we aimed to assess the role of incorporating self-certainty (SCe) by evaluating differences in mean token entropies and output token lengths across all model pairs. We used pairwise two-sided Welch's t-tests for these comparisons as they do not assume equal variances between groups. We then sought to evaluate the efficacy of our RL training regimen as opposed to that of supervised fine-tuning (SFT) by measuring macro-averaged accuracy across training steps. To estimate uncertainty, we used a non–parametric bootstrap with $1,000$ resamples, sampling with replacement within each task–cohort combination independently to derive 95% confidence intervals (CIs). The bootstrap median served as the point estimate, and the CIs were derived from the 2.5th and 97.5th percentiles of the bootstrap distribution. Lastly, we conducted additional experiments to assess LUNAR's performance on traditional medical knowledge benchmarks to ensure that model training did not impair its underlying medical knowledge, and compared its performance against baselines and an SFT model. The 95% confidence intervals were derived using non–parametric bootstrap resampling ($1,000$ resamples) within each benchmark prior to macro-averaging. Here, a non–parametric permutation test with $10,000$ permutations was used to assess differences in medians between models.

After our evaluation of the RL training efficacy of RL, we proceeded to measure LUNAR's performance on various ADRD diagnostic tasks, including identification of cognitive status, etiology of cognitive impairment, and classification of biomarker positivity. We also evaluated the alignment between LUNAR's answers and post-mortem neuropathological grades. All 95% confidence intervals were estimated using a non–parametric bootstrap (1,000 resamples) with sampling performed with replacement using a fixed random seed for reproducibility. The bootstrap median was used as the point estimate, and confidence intervals were obtained from the 2.5th and 97.5th percentiles of the bootstrap distribution, with resampling performed independently for each model–cohort combination. Statistical significance of performance differences between pairs of models was assessed using two-sided permutation tests conducted at the patient level. For each pairwise comparison, predictions from two models were aligned by patient ID and trial number, with each patient having

5 independent trials. Under the null hypothesis of no difference between models, patient-level label swaps were performed: for each of $10,000$ permutations, each unique patient ID was independently assigned to either retain or swap the predictions of the two models across all of that patient's trials. This ID-level permutation preserves within-patient correlation structure, ensuring that all trials belonging to the same patient are swapped together. For each permutation, the difference in the metric of interest between two models was computed, yielding a null distribution of metric differences. The two-sided p-value was calculated as the proportion of permuted absolute differences that were greater than or equal to the observed absolute difference. All p-values were corrected for multiple comparisons using the Benjamini-Hochberg false discovery rate procedure at $\alpha = 0.05$.

**Computational hardware and software.** The model was trained using Python 3.11.13 with CUDA 12.8, Triton 3.2.0, PyTorch 2.6.0, vLLM 0.8.5.post1, Transformers 4.52.4, TRL 0.19.0 and DeepSpeed 0.16.8. For evaluation and data analysis, we used Python 3.12.4 with pandas 2.2.2, numpy 1.26.3, matplotlib 3.9.1, PyTorch 2.4.0, scipy 1.14.0, scikit-learn 1.5.1, seaborn 0.13.2 and vLLM 0.12.0. All figures were finalized using Canva and Adobe Illustrator. Each training run used four NVIDIA H200 GPUs, with an average runtime of 9 hours (1,491 steps; 1 epoch). Inference on individual cases on an NVIDIA L40S GPU required less than one minute.

### Supplementary text

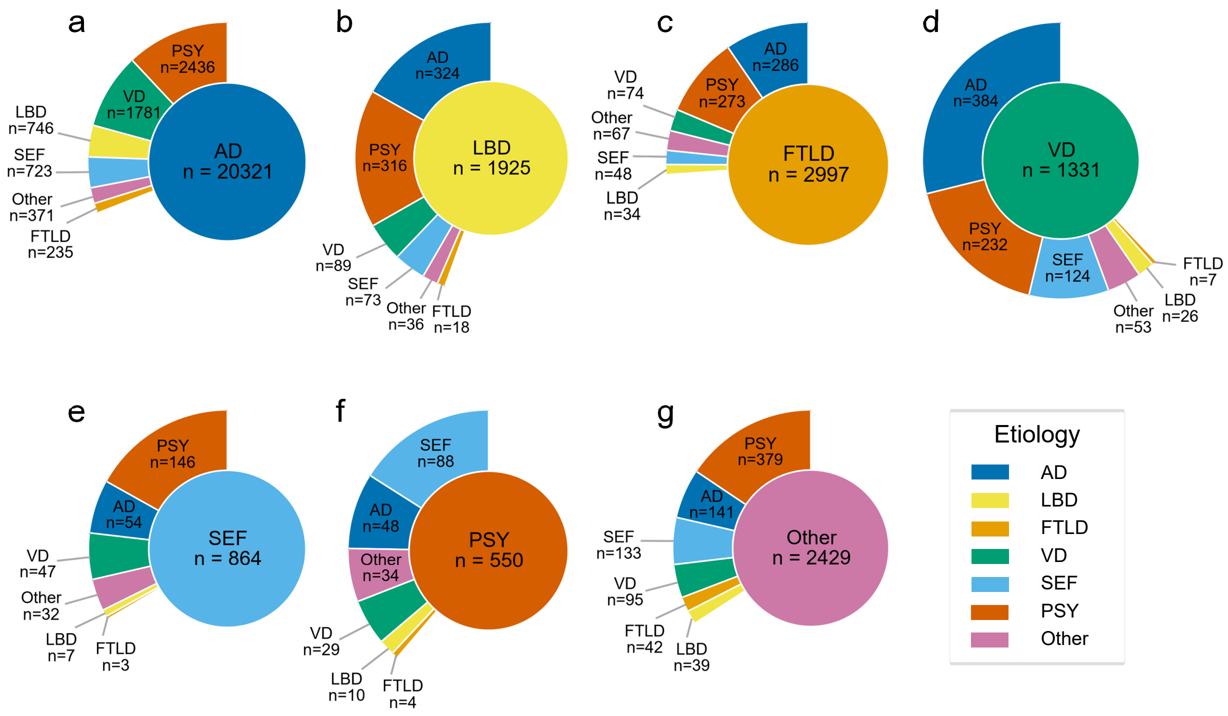


**Figure S1**: **NACC population breakdown by primary and secondary etiology.** a-g. Secondary etiology breakdown within each primary etiology (AD, LBD, FTLD,VD, SEF, PSY, or other). Comorbid secondary etiologies were treated as separate datapoints.

**JSON summarization prompt**

You will receive patient data enclosed within `<data>` and `</data>` tags. Carefully summarize this information accurately and objectively, without making any assumptions, interpretations, diagnoses, or conclusions. Focus solely on what is explicitly stated in the data. Your summary should address each subsection listed: {`json_keys`}. If neuropsychological battery summary scores are present, incorporate them appropriately. Present your summary in continuous prose using complete sentences. Do not use bullet points, numbered lists, or section headings; instead, craft coherent paragraphs and transition naturally between topics. Ensure that your response does not exceed 2,000 tokens.

```
<data>
{patient}
</data>
```

**Figure S2**: **Prompt used for narrative summarization of structured JSON person-level data.** The prompt instructs the language model (Qwen3-32B in non-thinking mode) to generate a concise, objective clinical narrative from the harmonized JSON input. It explicitly restricts the output to factual content within specified subsections ({`json_keys`}), prohibits interpretations or diagnoses, and enforces a continuous prose format without lists or headings to produce case presentations for downstream diagnostic reasoning.

Training and evaluation prompts (RL)

System prompt

Please reason step by step, and put your final answer within \\boxed{}.

User prompt (single correct answer)

Question: {`visit_summary`}

{`question`}

Answer Choices:

{`options`}

**Figure S3**: **Prompts used for training and evaluating the models trained using RL.** The system prompt instructs the model to perform explicit step-by-step reasoning and enclose its final answer within \\boxed{} delimiters, facilitating automated extraction and verification. The user prompt presents the model a person-level case narrative, one randomly assigned diagnostic question type (from the same six used in RL training), and randomly shuffled answer choices.

**Training and evaluation prompts (SFT)**

**System prompt**

Provide only the final answer in the format: The answer is \\boxed{option_letter}: option_text, with no reasoning.

**User prompt (single correct answer)**

Question: {`visit_summary`}

{`question`}

Answer Choices:
{`options`}

**Answer format**

The answer is \\boxed{option_letter}: option_text

**Figure S4**: **Prompts used for training and evaluating the models trained using SFT.** The system prompt instructs the model to provide the answer within \\boxed{} delimiters without any reasoning,. The user prompt presents the model a person-level case narrative, one randomly assigned diagnostic question type, and randomly shuffled answer choices. The answer format shows the target output format.

**Answer extraction**

You will be given a text enclosed within <text> and </text> tags. Your task is to extract the final answer from the text and match it with one of the option letter (or number) listed between <options> and </options>. Do not try to come up with a new answer, just identify what is in the text. Do not make assumptions beyond the provided content. Return only a single letter or number answer inside \\boxed{{}}. Do not output anything else.
Important: If you cannot identify the final answer symbol, output \\boxed{{-}}.
Example 1:
Example 1 text: ”... the final answer is \\boxed{{\\text{{Dementia (DE)}}}}.”
Example 1 options: ”A. Normal Cognition B. Mild Cognitive Impairment C. Dementia”
Example 1 output: ”\\boxed{{C}}”
Example 2:
Example 2 text: ”... ANSWER: \\boxed{{\\text{{Alzheimer’s Disease}}}}.”
Example 2 options: ”1. Alzheimer’s Disease 2. Lewy Body Dementia 3. Vascular Dementia”
Example 2 output: ”\\boxed{{1}}”
<text>
{answer}
</text>
<options>
{options}
</options>

**Figure S5**: **Prompt used for final answer extraction.** The prompt instructs Qwen3-14B to identify the final answer from a model-generated response and map it to the corresponding option letter or number provided in the candidate list. The model is required to return only a single answer enclosed in \\boxed{}, or \\boxed{-} when no final answer can be identified. This procedure was used when the original response did not contain a valid boxed answer or contained multiple boxed expressions.

Example patient data (NACC) as JSON (patient summary)


```
{
    "Subject Demographics": {
        "Subject's age at visit": [redacted],
        "Principal referral source": "Professional contact: clinician,
            nurse, doctor, other health care provider, or other staff (ADC
            and non-ADC)",
        "Subject's sex": [redacted],
        "Hispanic/Latino ethnicity": [redacted],
        "Race": [redacted],
        "Primary language": [redacted],
        "Years of education": [redacted],
        "Marital status": [redacted],
        "Living situation": [redacted],
        "Level of independence": [redacted],
        "Type of residence": [redacted],
        "Is the subject left- or right-handed?": "[redacted]-handed",
        "Derived NIH race definitions": [redacted]
    },
    "Subject Family History": {
        "Indicator of first-degree family member with cognitive impairment":
            [redacted],
        "Indicator of mother with cognitive impairment": [redacted],
        "Indicator of father with cognitive impairment": [redacted],
```

```
    "In this family, is there evidence for an AD mutation (from list of
       specific mutations)?": [redacted],
    "In this family, is there evidence for an FTLD mutation (from list
       of specific mutations)?": [redacted],
    "Source of evidence for [redacted],
    "In this family, is there evidence for a mutation other than an AD
       or FTLD mutation?": [redacted]
    "In this family, is there evidence for an FTLD mutation?": [redacted]
},
"Subject Medications": {
    "Subject taking any medications": "Yes",
    "Name of medications used within two weeks of UDS visit":
       "OMEPRAZOLE, ALBUTEROL, PSYLLIUM, CHOLECALCIFEROL,
       CALCIUM-VITAMIN D, MULTIVITAMIN, CARBIDOPA-LEVODOPA, CETIRIZINE,
       UBIQUINONE, FLUTICASONE-SALMETEROL",
    "Total number of medications reported at each visit (range: (0.0 - 40
       .0))": 10.0,
    "Reported current use of an antiparkinson agent": "Reported use at
       visit"
},
"Subject Health History": {
    "Heart attack/cardiac arrest": "Absent",
    "Atrial fibrillation": "Absent",
    "Angioplasty/endarterectomy/stent": "Absent",
    "Cardiac bypass procedure": "Absent",
    "Pacemaker and/or defibrillator": "Absent",
    "Congestive heart failure": "Absent",
    "Angina": "Absent",
    "Heart valve replacement or repair": "Absent",
```

```
"Other cardiovascular disease": "Absent",
"Stroke": "Absent",
"Transient ischemic attack (TIA)": "Absent",
"Parkinson's disease (PD)": "Absent",
"Other parkinsonian disorder": "Recent/Active",
"Seizures": "Absent",
"History of traumatic brain injury (TBI)": "No",
"Traumatic brain injury (TBI)": "Absent",
"Traumatic brain injury (TBI) with brief loss of consciousness":
    "No",
"TBI with extended loss of consciousness - 5minutes of longer": "No",
"TBI without loss of consciousness - as might result from military
    detonations or sports injury": "No",
"Diabetes": "Absent",
"Hypertension": "Absent",
"Hypercholesterolemia": "Absent",
"Vitamin b12 deficiency": "Absent",
"Thyroid disease": "Absent",
"Arthritis": "Absent",
"Arthritis, region affected - upper extremity": "No",
"Arthritis, region affected - lower extremity": "No",
"Arthritis, region affected - spine": "No",
"Region affected - unknown": "No",
"Incontinence - urinary": "Absent",
"Incontinence - bowel": "Absent",
"Sleep apnea history reported at Initial Visit": "Absent",
"REM sleep behavior disorder (RBD) history reported at Initial
    Visit": "Absent",
"Hyposomnia/insomnia history reported at Initial Visit": "Absent",
"Other sleep disorder history reported at Initial Visit": "Absent",
```

```
    "Alcohol abuse - clinically significant occurring over a 12-month
      period manifested in one of the following areas: work, driving,
      legal, or social": "Absent",
    "Other abused substances - clinically significant impairment
      occurring over a 12-month period manifested in one of the
      following areas: work, driving, legal, or social": "Absent",
    "Post-traumatic stress disorder (PTSD)": "Absent",
    "bipolar disorder": "Absent",
    "Schizophrenia": "Absent",
    "Active depression in the last two years": "No",
    "Depression episodes more than two years ago": "No",
    "Anxiety": "Absent",
    "Obsessive-compulsive disorder (OCD)": "Absent",
    "Developmental neuropsychiatric disorders (e.g., autism spectrum
      disorder [ASD], attention-deficit hyperactivity disorder [ADHD],
      dyslexia)": "Absent",
    "Other psychiatric disorder": "Absent"
  },
  "Physical": {
    "Subject’s height (inches) (range: (36.0 - 87.9))": 60.0,
    "Subject’s weight (lbs) (range: (50.0 - 400.0))": 140.0,
    "body mass index (BMI) (range: (10.0 - 100.0))": 27.3
  },
  "Physical/Neurological Exam Findings": {
    "Were all findings unremarkable?": "Abnormal findings",
    "Were there abnormal neurological exam findings?": "Yes - abnormal
      findings were consistent with syndromes listed in Questions 2-8",
    "Parkinsonian signs": "Yes",
    "Slowing of fine motor movements - left side": "Yes",
    "Slowing of fine motor movements - right side": "Yes",
```

```
"Rigidity - left arm": "Yes",
"Rigidity - right arm": "Yes",
"bradykinesia": "Yes",
"Parkinsonian gait disorder": "Yes",
"Postural instability": "Yes",
"Neurological sign considered by examiner to be most likely
   consistent with cerebrovascular disease": "No",
"Cortical cognitive deficit (e.g., aphasia, apraxia, neglect)": "No",
"Focal or other neurological findings consistent with SIVD
   (subcortical ischemic vascular dementia)": "No",
"Motor (may include weakness of combination of face, arm, and leg;
   reflex changes, etc.) - left side": "No",
"Motor (may include weakness of combination of face, arm, and leg;
   reflex changes, etc.) - right side": "No",
"Cortical visual field loss - left side": "No",
"Cortical visual field loss - right side": "No",
"Somatosensory loss - left side": "No",
"Somatosensory loss - right side": "No",
"Higher cortical visual problem suggesting posterior cortical
   atrophy (e.g., prosopagnosia, simultagnosia, balint's syndrome)
   or apraxia of gaze": "No",
"Findings suggestive of progressive supranuclear palsy (PSP),
   corticobasal syndrome (CBS), or other related disorders": "Yes",
"Eye movement changes consistent with PSP": "Yes",
"Dysarthria consistent with PSP": "Yes",
"Axial rigidity consistent with PSP": "Yes",
"Gait disorder consistent with PSP": "Yes",
"Apraxia of speech": "Yes",
"Ataxia consistent with CBS - left side": "No",
"Ataxia consistent with CBS - right side": "No",
```

```
    "Alien limb consistent with CBS - left side": "No",
    "Alien limb consistent with CBS - right side": "No",
    "Dystonia consistent with CBS, PSP, or related disorder - left
       side": "Yes",
    "Dystonia consistent with CBS, PSP, or related disorder - right
       side": "Yes",
    "Myoclonus consistent with CBS - left side": "No",
    "Myoclonus consistent with CBS - right side": "No",
    "Findings suggesting ALS (e.g., muscle wasting, fasciculations,
       upper motor and/or lower motor neuron signs)": "No",
    "Normal pressure hydrocephalus - gait apraxia": "No",
    "Other findings (e.g., cerebella ataxia, chorea, myoclonus)": "No"
},
"Neuropsychological Battery Summary Scores": {
    "The remainder of the battery was administered:": "In ADC/clinic",
    "Language of test administration": "English",
    "Total score for copy of benson figure (range: (0.0 - 17.0))": 12.0,
    "Total score for 10- to 15-minute delayed drawing of benson figure
       (range: (0.0 - 17.0))": 12.0,
    "Recognized original stimulus from among four options": "Yes",
    "Animals - Total number of animals named in 60seconds (range: (0.0 -
       77.0))": 12.0,
    "Vegetable - Total number of vegetables named in 60seconds (range:
       (0.0 - 77.0))": 4.0,
    "Trail Making Test Part A - Total number of seconds to complete
       (range: (0.0 - 150.0))": "Unable to complete due to Other
       problem",
    "Part A - Number of commission errors (range: (0.0 - 40.0))":
       "Unable to complete due to Other problem",
```

"Part A - Number of correct lines (range: (0.0 - 24.0))": "Unable to complete due to Other problem",
"Trail Making Test part B - Total number of seconds to complete (range: (0.0 - 300.0))": "Unable to complete due to Other problem",
"part B - Number of commission errors (range: (0.0 - 40.0))": "Unable to complete due to Other problem",
"part B - Number of correct lines (range: (0.0 - 24.0))": "Unable to complete due to Other problem",
"Number of correct F-words generated in 1minute (range: (0.0 - 40.0))": 5.0,
"Number of F-words repeated in 1minute (range: (0.0 - 15.0))": 2.0,
"Number of non-F-words and rule violation errors in 1minute (range: (0.0 - 15.0))": 0.0,
"Number of correct L-words generated in 1minute (range: (0.0 - 40.0))": 1.0,
"Number of L-words repeated in 1minute (range: (0.0 - 15.0))": 0.0,
"Number of non-L-words and rule violation errors in 1minute (range: (0.0 - 15.0))": 1.0,
"Total number of correct F-words and L-words (range: (0.0 - 80.0))": 6.0,
"Total number of F-word and L-word repetition errors (range: (0.0 - 30.0))": 2.0,
"Total number of non-F/L-words and rule violation errors (range: (0.0 - 30.0))": 1.0,
"Per clinician, based on the neuropsychological examination, the subject's cognitive status is deemed": "Three or more scores are abnormal or lower than expected",
"Was any part of MoCA administered?": "Yes",
"Where was MoCA administered?": "In ADC/clinic",

```
"Language of MoCA administration": "English",
"Subject was unable to complete one or more sections due to visual
   impairment": "No",
"Subject was unable to complete one or more sections due to hearing
   impairment": "No",
"MoCA Total Raw Score - uncorrected (range: (0.0 - 30.0))": 22.0,
"MoCA: Visuospatial/executive - Trails (range: (0.0 - 1.0))": 1.0,
"MoCA: Visuospatial/executive - Cube (range: (0.0 - 1.0))": 0.0,
"MoCA: Visuospatial/executive - Clock contour (range: (0.0 - 1.0))":
   1.0,
"MoCA: Visuospatial/executive - Clock numbers (range: (0.0 - 1.0))":
   0.0,
"MoCA: Visuospatial/executive - Clock hands (range: (0.0 - 1.0))": 0
   .0,
"MoCA: Language - Naming (range: (0.0 - 3.0))": 2.0,
"MoCA: Memory - Registration (two trials) (range: (0.0 - 10.0))": 7
   .0,
"MoCA: Attention - Digits (range: (0.0 - 2.0))": 2.0,
"MoCA: Attention - Letter A (range: (0.0 - 1.0))": 0.0,
"MoCA: Attention - Serial 7s (range: (0.0 - 3.0))": 3.0,
"MoCA: Language - Repetition (range: (0.0 - 2.0))": 1.0,
"MoCA: Language - Fluency (range: (0.0 - 1.0))": 1.0,
"MoCA: Abstraction (range: (0.0 - 2.0))": 1.0,
"MoCA: Delayed recall - No cue (range: (0.0 - 5.0))": 4.0,
"MoCA: Delayed recall - Category cue (range: (0.0 - 5.0))": 1.0,
"MoCA: Orientation - Date (range: (0.0 - 1.0))": 1.0,
"MoCA: Orientation - Month (range: (0.0 - 1.0))": 1.0,
"MoCA: Orientation - Year": 1.0,
"MoCA: Orientation - Day (range: (0.0 - 1.0))": 1.0,
"MoCA: Orientation - Place (range: (0.0 - 1.0))": 1.0,
```

```
"MoCA: Orientation - City (range: (0.0 - 1.0))": 1.0,
"MoCA Total Score - corrected for education (range: (0.0 - 30.0))": 2
    2.0,
"Craft Story 21Recall (Immediate) - Total story units recalled,
    verbatim scoring (range: (0.0 - 44.0))": 13.0,
"Craft Story 21Recall (Immediate) - Total story units recalled,
    paraphrase scoring (range: (0.0 - 25.0))": 13.0,
"Number Span Test: Forward - Number of correct trials (range: (0.0 -
    14.0))": 6.0,
"Number Span Test: Forward - Longest span forward (range: (0.0 - 9
    .0))": 6.0,
"Number Span Test: backward - Number of correct trials (range: (0.0
    - 14.0))": 6.0,
"Number Span Test: backward - Longest span backward (range: (0.0 - 8
    .0))": 5.0,
"Craft Story 21Recall (Delayed) - Total story units recalled,
    verbatim scoring (range: (0.0 - 44.0))": 6.0,
"Craft Story 21Recall (Delayed) - Total story units recalled,
    paraphrase scoring (range: (0.0 - 25.0))": 8.0,
"Craft Story 21Recall (Delayed) - Delay time (range: (0.0 - 85.0))":
    22.0,
"Craft Story 21Recall (Delayed) - Cue (boy) needed": "Yes",
"Multilingual Naming Test (MINT) - Total score (range: (0.0 - 32
    .0))": 26.0,
"Multilingual Naming Test (MINT) - Total correct without semantic
    cue (range: (0.0 - 32.0))": 24.0,
"Multilingual Naming Test (MINT) - Semantic cues: Number given
    (range: (0.0 - 32.0))": 4.0,
"Multilingual Naming Test (MINT) - Semantic cues: Number correct
    with cue (range: (0.0 - 32.0))": 2.0,
```

```
    "Multilingual Naming Test (MINT) - Phonemic cues: Number given
       (range: (0.0 - 32.0))": 6.0,
    "Multilingual Naming Test (MINT) - Phonemic cues: Number correct
       with cue (range: (0.0 - 32.0))": 1.0
},
"Clinician-assessed Medical Conditions": {
    "Cancer present in the last 12months (excluding non-melanoma skin
       cancer), primary or metastatic": "No",
    "Diabetes present at visit": "No",
    "Myocardial infarct present within the past 12months": "No",
    "Congestive heart failure present": "No",
    "Atrial fibrillation present": "No",
    "Hypertension present": "No",
    "Angina present": "No",
    "Hypercholesterolemia present": "No",
    "B12 deficiency present": "No",
    "Thyroid disease present": "No",
    "Arthritis present": "No",
    "Arthritis type": "No arthritis reported",
    "Arthritis region affected - upper extremity": "No arthritis
       reported",
    "Arthritis region affected - lower extremity": "No arthritis
       reported",
    "Arthritis region affected - spine": "No arthritis reported",
    "Arthritis region affected - unknown": "No arthritis reported",
    "Incontinence present - urinary": "Yes",
    "Incontinence present - bowel": "No",
    "Sleep apnea present": "No",
    "REM sleep behavior disorder (RBD) present": "No",
    "Hyposomnia/insomnia present": "No",
```

```
        "Other sleep disorder present": "No",
        "Carotid procedure: angioplasty, endarterectomy, or stent within the
            past 12months": "No",
        "Percutaneous coronary intervention: angioplasty and/or stent within
            the past 12months": "No",
        "Procedure: pacemaker and/or defibrillator within the past 12
            months": "No",
        "Procedure: heart valve replacement or repair within the past 12
            months": "No",
        "Antibody-mediated encephalopathy within the past 12months": "No",
        "Other medical conditions or procedures within the past 12months not
            listed above": "Yes",
        "Other medical conditions specification": "Scoliosis"
    },
    "Imaging evidence": {
        "Hippocampal atrophy": "No",
        "Structural MR evidence for frontal or anterior temporal atrophy for
            FTLD": "Yes",
        "Large vessel infarct(s)": "No",
        "Lacunar infarct(s)": "No",
        "Macrohemorrhage(s)": "No",
        "Microhemorrhage(s)": "No",
        "Moderate white-matter hyperintensity (CHS score 5-6)": "No",
        "Extensive white-matter hyperintensity (CHS score 7-8)": "No"
    }
}
```

**Figure S6**: **Example of harmonized person-level data from the NACC cohort (`patient_summary`).** Representative structured JSON representation of a single participant record after cohort-specific harmonization and preprocessing. The nested structure organizes objective clinical data into modality-specific sections (e.g., demographics, family history, medications, health history, physical/neurological findings, neuropsychological scores, clinician-assessed conditions and imaging evidence), serving as input to the narrative summarization pipeline for downstream diagnostic reasoning.

Example patient data as JSON (NIFD) (patient summary)


```
{
    'Subject Demographics': {
        'Gender': [redacted],
        'Years of Education': [redacted],
        'Race': [redacted],
        'Age': [redacted]
    },
    "Unified Parkinson's Disease Rating Scale (UPDRS)": {
        "MD Exam - Unified Huntington's Disease Rating Scale": 5,
        "MD Exam - Unified Parkinson's Disease Rating Scale": 2
    },
    'Geriatric Depression Scale (GDS)': {
        'Geriatric Depression Scale - Total (30 item)': 0
    },
    'Activities of Daily Living': {
        'Schwab and England Activities of Daily Living - Score': 100
    },
    'Neuropsychological Battery Summary Scores': {
        'Mini Mental State Exam - Total score': 28,
        'Modified Trails - Completion time, in seconds': 23,
        'Modified Trails - Correct lines': 14,
        'Modified Trails - Errors': 1,
        'Digit Span - Maximum forward recall span': 6,
        'Digit Span - Maximum backward recall span': 4,
        'Verbal Fluency - D Words - Correct D words generated': 16,
        'Verbal Fluency - Animals - Correct animals generated': 26,
        'Boston Naming Test (15-item) - BNT Total': 15,
        'Montreal Cognitive Assessment': 15,
        'Modified Benson Figure': 14,
```

```
    'Peabody Picture Vocabulary Test - Verbal': 4,
    'Peabody Picture Vocabulary Test - Descriptive': 4,
    'Peabody Picture Vocabulary Test - Animate': 4,
    'Peabody Picture Vocabulary Test - Inanimate': 4,
    'Montreal Cognitive Assessment - Total score, with adjustment for
      education level': 29,
    'Montreal Cognitive Assessment - Administered before or after the
      remaining neuropsychological battery': 'Before'
  },
  'Imaging evidence': {
    'MRI Report': {
      'MRI features extraction procedure': 'A Generalized Additive
        Model for Location, Scale, and Shape (GAMLSS) was fitted to
        FastSurfer-derived regional brain volumes acquired from 2,087
        healthy controls, following the Brain charts for the human
        lifespan methodology published in Nature (2022). Data were
        aggregated from five independent cohorts, and the analysis
        included region-of-interest (ROI) volumes for the medial and
        lateral temporal lobes, medial and lateral parietal lobes,
        frontal, occipital lobes, as well as the inferior lateral
        ventricles and hippocampus. To ensure a robust normative
        reference, participants with any known neurodegenerative
        disease were excluded and subjects with extreme brain volumes
        (>3.5 absolute z-scores) were excluded during quality control
        prior to model fitting. The Brain Charts pipeline was
        implemented to model age- and sex-specific normative centile
        curves for each ROI. Centile scores (scaled 0-1) were
        computed for both healthy controls and patient samples.
        Abnormality was defined based on a one-sided z-score
        threshold: mild atrophy/enlargement for scores corresponding
```

```
            to 1standard deviation (z=1), moderate for 1.5 SD (z=1.5),
            and severe for 2SD (z=2). For all lobar ROIs, lower centiles
            indicated greater atrophy, while for ventricular ROIs, higher
            centiles indicated enlargement (i.e., abnormality). The
            resulting findings are as follows: ',
        'T1 MRI Findings': 'No significant findings. Brain MRI appears
            age-appropriate.'
      }
   }
}
```


**Figure S7: Example of harmonized person-level data from the NIFD cohort (`patient_summary`).** Representative structured JSON representation of a single participant record after cohort-specific harmonization and preprocessing. The nested structure organizes objective clinical data into modality-specific sections (e.g., demographics, family history, medications, health history, physical/neurological findings, neuropsychological scores, clinician-assessed conditions and imaging evidence), serving as input to the narrative summarization pipeline for downstream diagnostic reasoning.

Converted text summary {visit summary}

The subject is a [redacted]. She is [redacted]-handed, speaks [redacted] as her primary language, and has completed [redacted] years of education. She is [redacted] and [redacted]. She requires some assistance with complex activities but is otherwise independent. The principal referral source for this evaluation is a professional contact, such as a clinician, nurse, or other healthcare provider.

In terms of family history, [redacted]

The subject is currently taking multiple medications, including OMEPRAZOLE, ALBUTEROL, PSYLLIUM, CHOLECALCIFEROL, CALCIUM-VITAMIN D, MULTIVITAMIN, CARBIDOPA-LEVODOPA, CETIRIZINE, UBIQUINONE, and FLUTICASONE-SALMETEROL, totaling 10 medications. She is reported to be using an antiparkinson agent at the time of the visit.

Regarding her health history, she does not have any cardiovascular conditions such as heart attack, atrial fibrillation, stroke, or angina. There is no history of traumatic brain injury, diabetes, hypertension, hypercholesterolemia, or vitamin B12 deficiency. She does not have a history of substance abuse, psychiatric disorders, or sleep disorders. However, urinary incontinence is present, while bowel incontinence is absent. An additional medical condition noted is scoliosis.

The subject’s physical characteristics include a height of 60 inches and a weight of 140 pounds, resulting in a body mass index of 27.3. On the physical and neurological examination, findings are abnormal, with evidence of parkinsonian signs, including slowing of fine motor movements on both sides, rigidity in both arms, bradykinesia, parkinsonian gait disorder, and postural instability. Eye movement changes, dysarthria, axial rigidity, and gait disorder consistent with progressive supranuclear palsy (PSP) are also present. Dystonia consistent with PSP or related disorders is noted bilaterally. No findings are consistent with cerebrovascular disease, subcortical ischemic vascular dementia, or corticobasal syndrome. There is no evidence of amyotrophic lateral sclerosis or normal pressure hydrocephalus.

Neuropsychological testing was conducted in English and administered in the ADC/clinic setting. The subject performed with some difficulty, particularly on the Trail Making Test, which could not be completed due to unspecified issues. The copy and delayed drawing of the Benson figure both resulted in a score of 12 out of 17, and the subject was able to recognize the original stimulus from among four options. In the fluency task, she generated 12 animals and 4 vegetables in one minute. On the Montreal Cognitive Assessment (MoCA), the subject achieved a raw score of 22 out of 30, indicating three or more scores that were abnormal or lower than expected. Performance varied across domains, with partial success in visuospatial tasks, attention, and memory. The subject also completed the Craft Story 21 Recall and the Multilingual Naming Test (MINT), with scores indicating moderate recall and naming ability.

Clinically assessed medical conditions show no evidence of cancer, diabetes, heart conditions, or thyroid disease within the past 12 months. While urinary incontinence is present, bowel incontinence is not. No sleep disorders or recent surgical procedures are reported, though scoliosis is

```
listed as an additional medical condition.

Imaging findings indicate no hippocampal atrophy or evidence of large
vessel or lacunar infarcts, microhemorrhages, or extensive white-matter
hyperintensity. However, there is structural MRI evidence of frontal or
anterior temporal atrophy consistent with FTLD.
```

**Figure S8**: **Example of converted narrative clinical summary (`visit_summary`).** Representative output from the summarization pipeline, transforming the structured JSON person-level data into continuous, objective prose. The narrative preserves factual content from all modality-specific subsections (demographics, family history, medications, health history, physical/neurological examination, neuropsychological testing, clinician-assessed conditions and imaging evidence) while maintaining a coherent, clinician-readable format without headings, lists or interpretive statements, as required for downstream diagnostic reasoning tasks.

**Example model response `COG task`**

```
Given the patient's detailed profile, the cognitive impairment is most
closely aligned with Frontotemporal Lobar Degeneration (FTLD), specifically
involving the FUS mutation. Here is why:

1. Multiple medications indicative of frontotemporal lobar degeneration: The
patient is taking an antiparkinson agent, which aligns with the progressive
supranuclear palsy (PSP) features, but also can be symptomatic of FTLD.

2. Neurological signs: She has parkinsonian signs, progressive supranuclear
palsy (PSP), and neurological features that fit FUS FTLD with PSP overlap,
including gait and motor disturbances.

3. Neuropsychological testing: While the test was not completed, her
performance was within a range that suggests mild to moderate cognitive
impairment, far from normal cognition.
```

```
4. Neuroimaging findings: Note of frontal or anterior temporal atrophy,
consistent with FTLD.

Given the structural MRI and multiple atrophy findings, the patient is
suffering from a condition consistent with FUS FTLD. Thus, the patient is
now categorized under Mild Cognitive Impairment (MCI).

The final answer is \boxed{C. Mild Cognitive Impairment (MCI)}.
```

**Figure S9**: **Example model response on the cognitive status task (COG).** Representative output from LUNAR illustrating chain-of-thought reasoning followed by a boxed final answer. The response integrates multimodal clinical data (family history, medications, neurological examination, neuropsychological testing, and imaging) to arrive at a syndromic classification, demonstrating the model's ability to synthesize evidence and provide structured diagnostic impressions suitable for downstream evaluation.

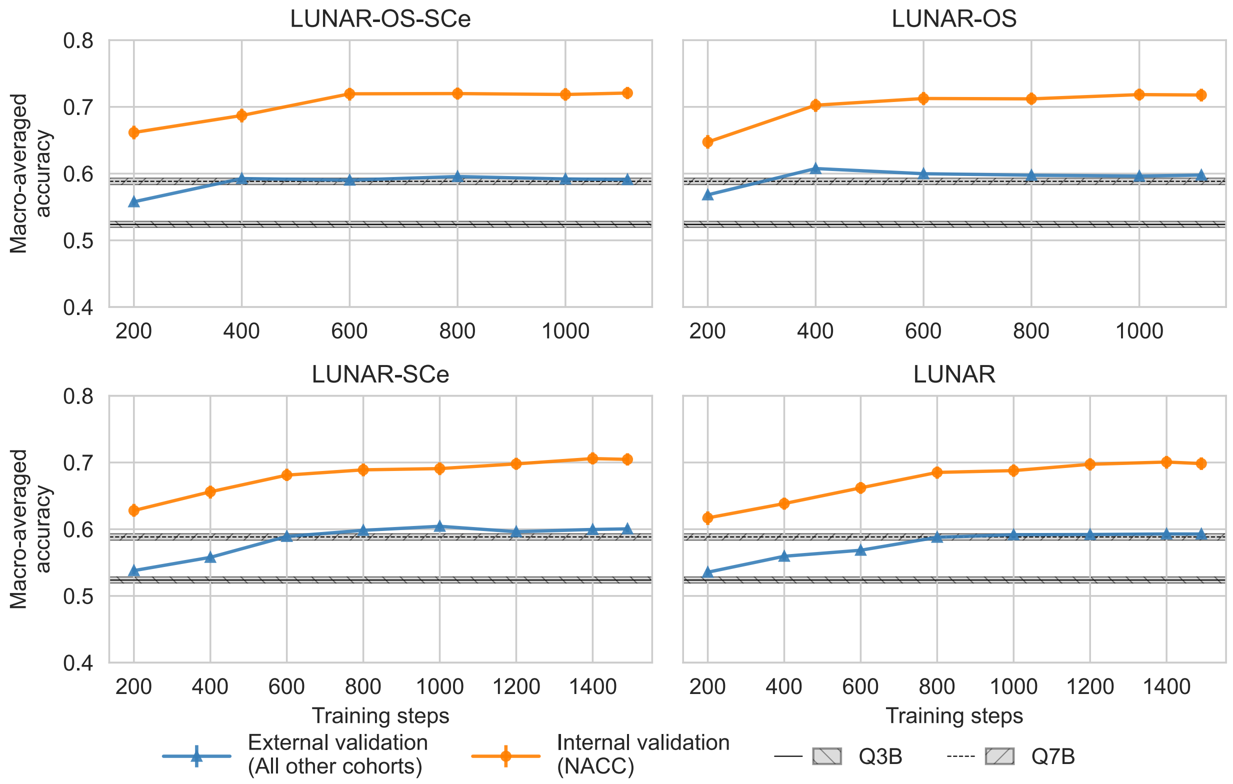


**Figure S10**: **Performance of ablated models over training steps.** Macro-averaged accuracy as a function of training steps for LUNAR and it's ablated variants (LUNAR-OS, LUNAR-SCe, LUNAR-OS-SCe). Performance is shown separately for the internal validation set (NACC; orange) and external validation cohorts (blue), with 95% bootstrap confidence intervals (non–parametric bootstrap, 1,000 iterations resampling individuals within task–cohort combinations before macro-averaging). Dashed gray lines and shaded areas represent baseline performance (Q3B and Q7B) with their 95% bootstrap CIs (baselines do not distinguish internal and external validation, as they were not trained).

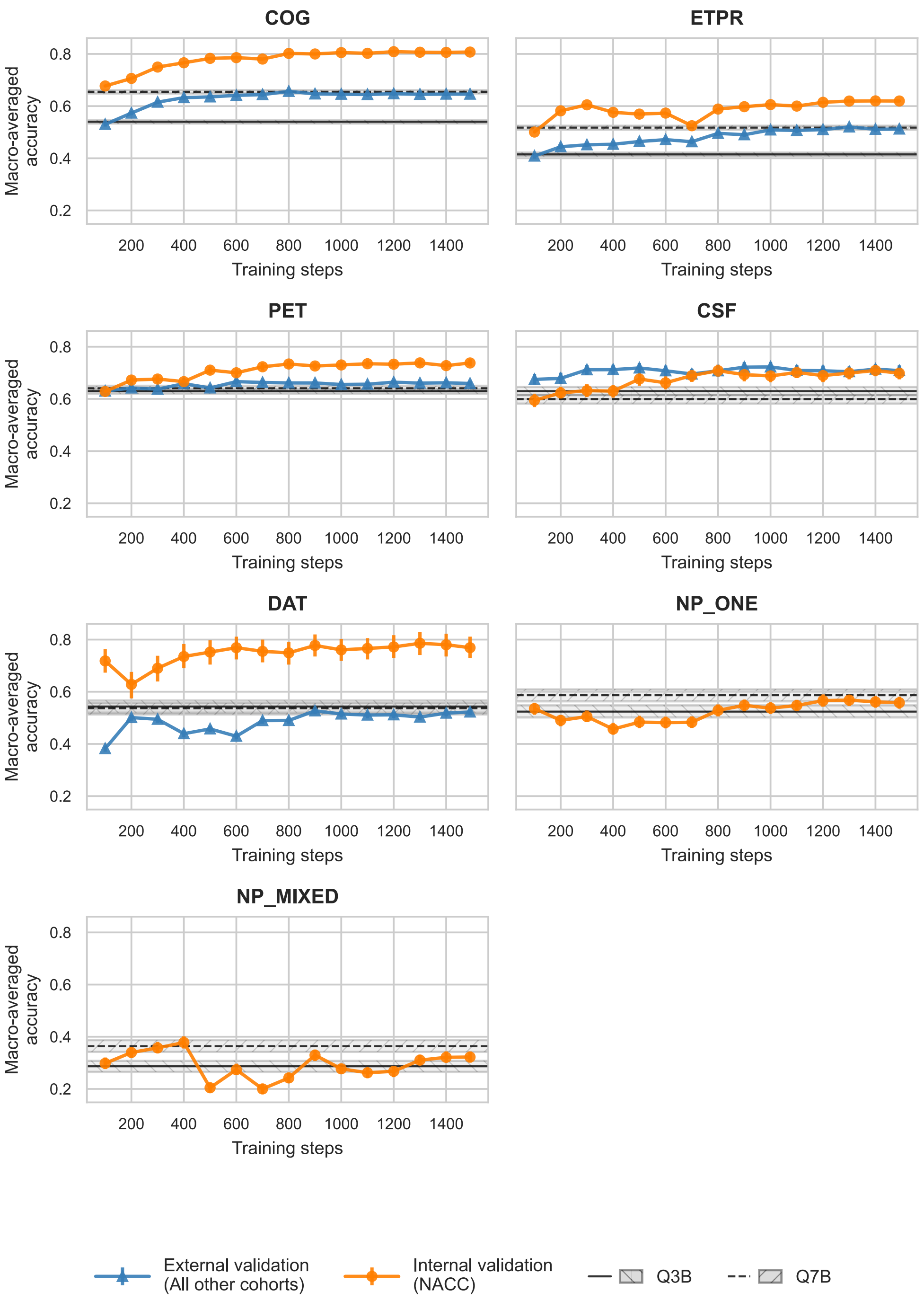


**Figure S11**: **Performance of LUNAR over training steps for each benchmark.** Macro-averaged accuracy as a function of training steps of LUNAR separated by task. Performance is shown separately for the internal validation set (NACC; orange) and external validation cohorts (blue), with 95% bootstrap confidence intervals (non–parametric bootstrap, 1,000 iterations resampling individuals within task–cohort combinations before macro-averaging). Dashed gray lines and shaded areas represent baseline performance (Q3B and Q7B) with their 95% bootstrap CIs (baselines do not distinguish internal and external validation, as they were not trained).

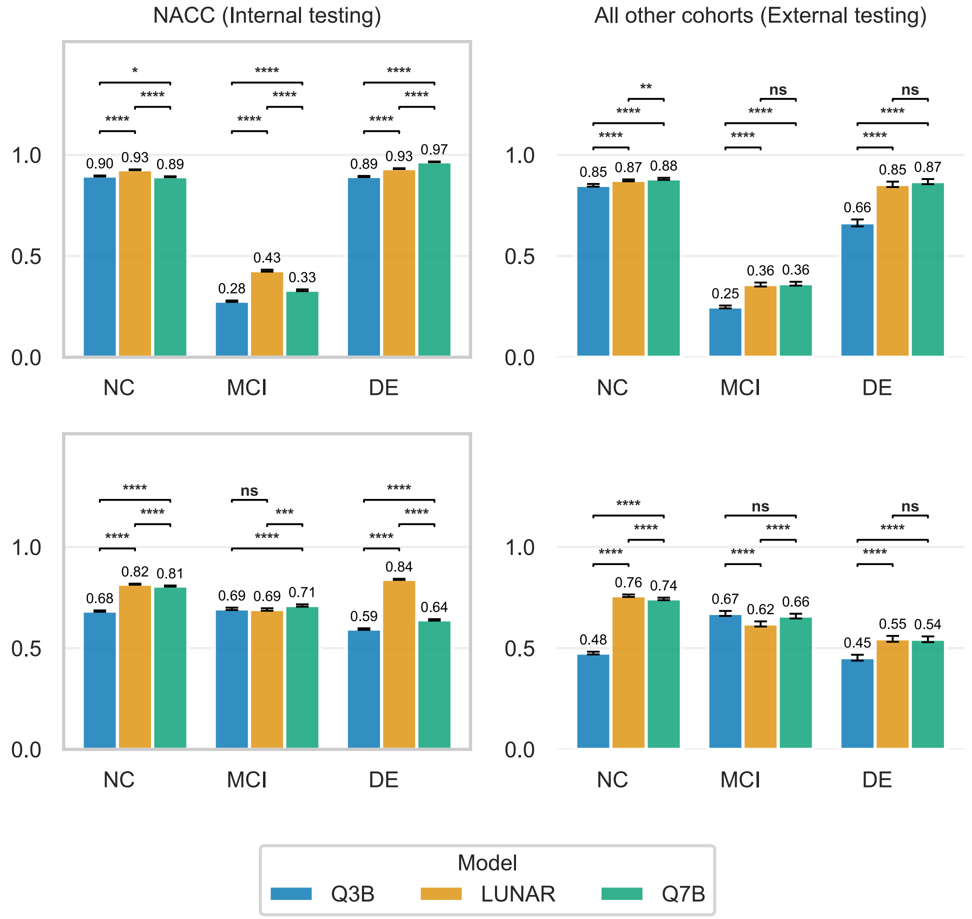


**Figure S12**: **Performance of LUNAR on COG task.** Class-wise Precision and Recall scores for the cognitive classification task COG: normal cognition (NC), mild cognitive impairment (MCI), dementia (DE) on internal testing (NACC; left) and external cohorts (all others; right). For recall we report the ground truth label, and for precision the predicted label on the horizontal axis. Bars represent medians from 1,000 bootstrap samples, with error bars showing 95% bootstrap confidence intervals (2.5th and 97.5th percentiles). The statistical significance of pairwise differences (LUNAR versus Q3B/Q7B) is from non–parametric, two-sided permutation tests ($10,000$ permutations at the participant level). The p-values were adjusted using false discovery rate (FDR) correction via the Benjamini-Hochberg procedure, applied within each cohort, class, and metric for panels (a), (c), and (d), and within each cohort and metric for panel (b). Significance levels are denoted as 'ns' (not significant) for $p \geq 0.05$; * for $p < 0.05$; ** for $p < 0.01$; *** for $p < 0.001$; and **** for $p < 0.0001$.

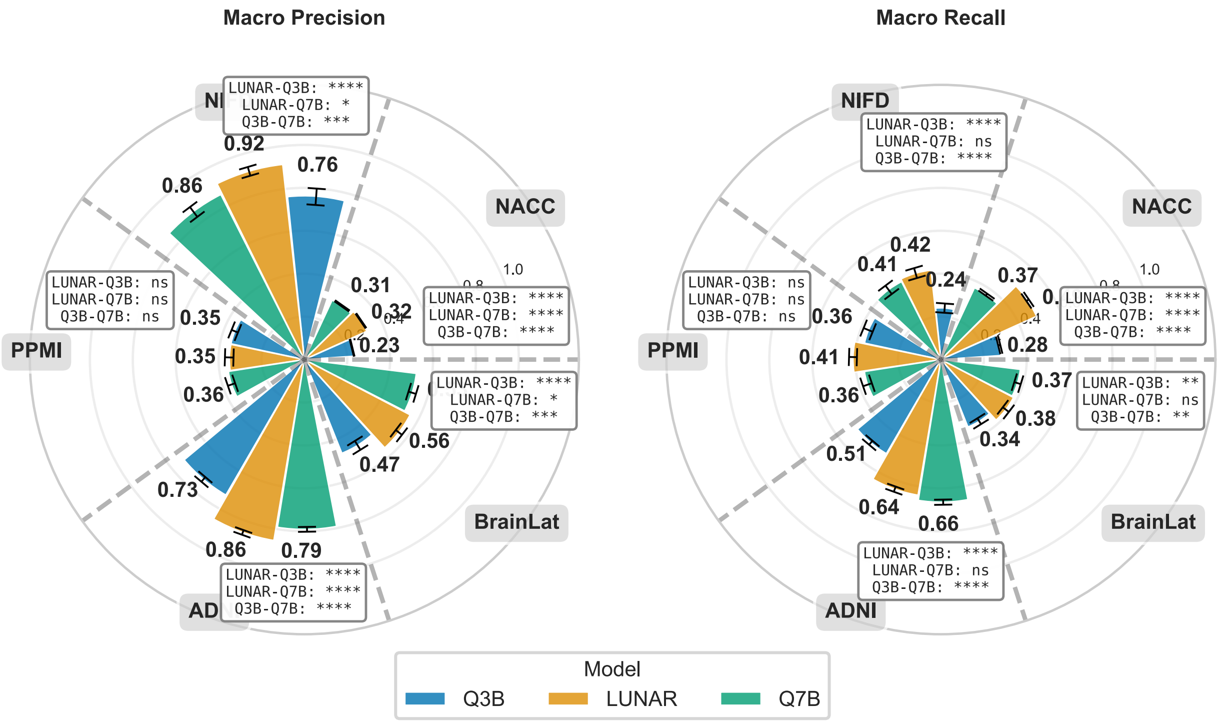


**Figure S13**: **Performance of LUNAR on ETPR task.** Radial (circular) bar plot of macro precision and macro recall scores for the etiology classification task (ETPR) across cohorts (NACC, ADNI, BrainLat, NIFD and PPMI), comparing LUNAR against baseline models (Q3B and Q7B). Each sector corresponds to a cohort, with bars indicating the medians and 95% bootstrap CIs; statistical annotations reflect pairwise comparisons. The p-values were adjusted using false discovery rate (FDR) correction via the Benjamini-Hochberg procedure, applied within each cohort, class, and metric for panels (a), (c), and (d), and within each cohort and metric for panel (b). Significance levels are denoted as 'ns' (not significant) for $p \geq 0.05$; * for $p < 0.05$; ** for $p < 0.01$; *** for $p < 0.001$; and **** for $p < 0.0001$.

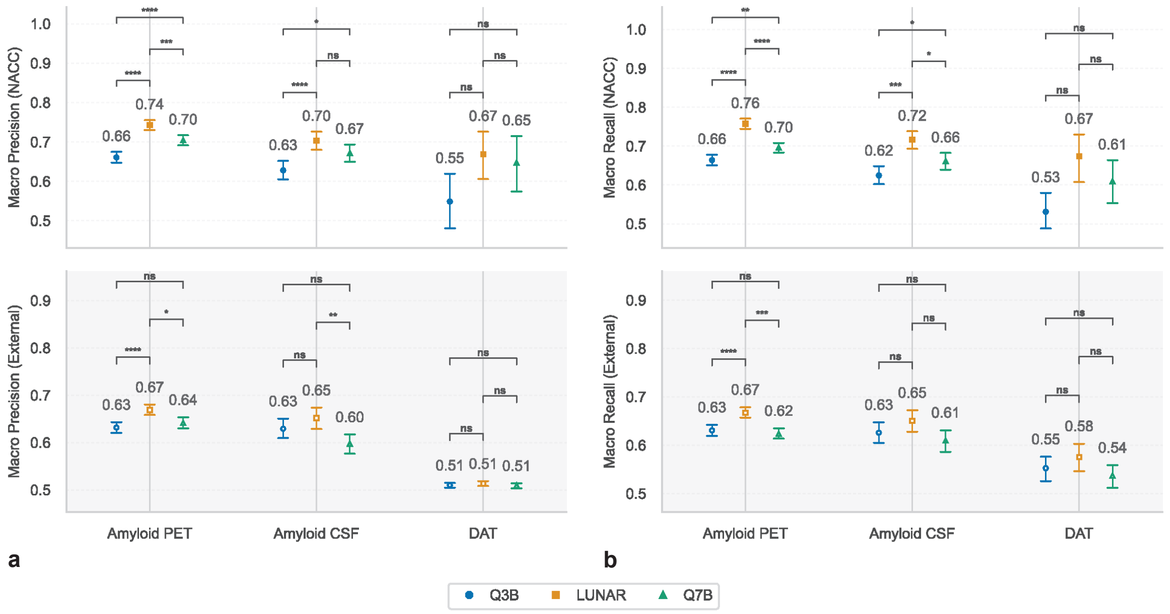


**Figure S14**: **Model performance on biomarker positivity prediction tasks.** Macro-average precision (a) and recall (b) are reported for Q3B, LUNAR, and Q7B on amyloid PET, amyloid CSF, and DaT positivity prediction tasks. Top rows show internal validation performance in NACC (amyloid PET: $n = 808$; amyloid CSF: $n = 288$; DaT: $n = 71$), while bottom rows show external validation in ADNI (amyloid PET: n=$1,392$, amyloid CSF:$n = 406$) and PPMI (n=1276) Median performance with 95% bootstrap confidence intervals is shown. Statistical significance of pairwise differences in medians was assessed via non–parametric permutation tests (10,000 permutations) with FDR correction. Significance levels are denoted as * $p < 0.05$, ** $p < 0.01$, *** $p < 0.001$, **** $p < 0.0001$.

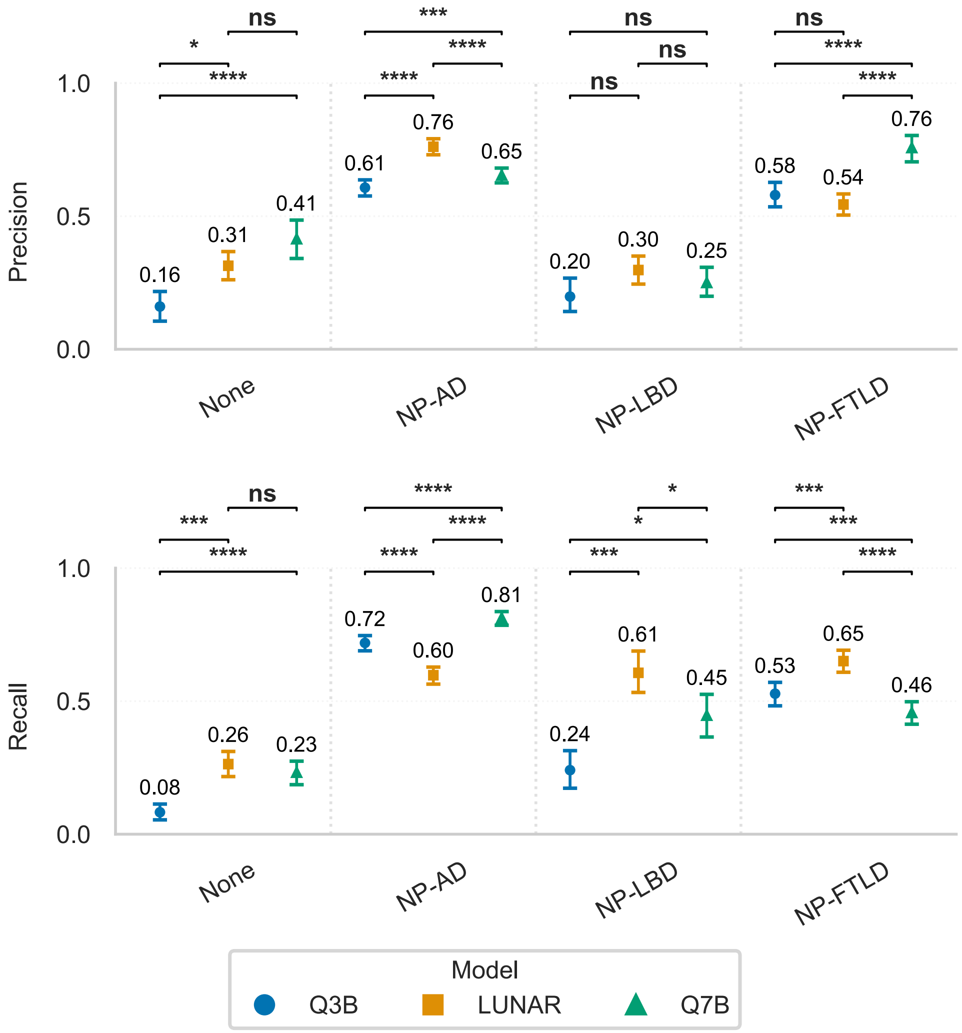


**Figure S15**: **Model performance on neuropathology one-class (NP-ONE) task.** Class-wise precision and recall scores for the neuropathology one-class (NP-ONE) task. Median performance with 95% bootstrap confidence intervals is shown. Statistical significance of pairwise differences in medians was assessed via non–parametric permutation tests (10,000 permutations) with FDR correction. Significance levels are denoted as * $p < 0.05$, ** $p < 0.01$, *** $p < 0.001$, **** $p < 0.0001$.

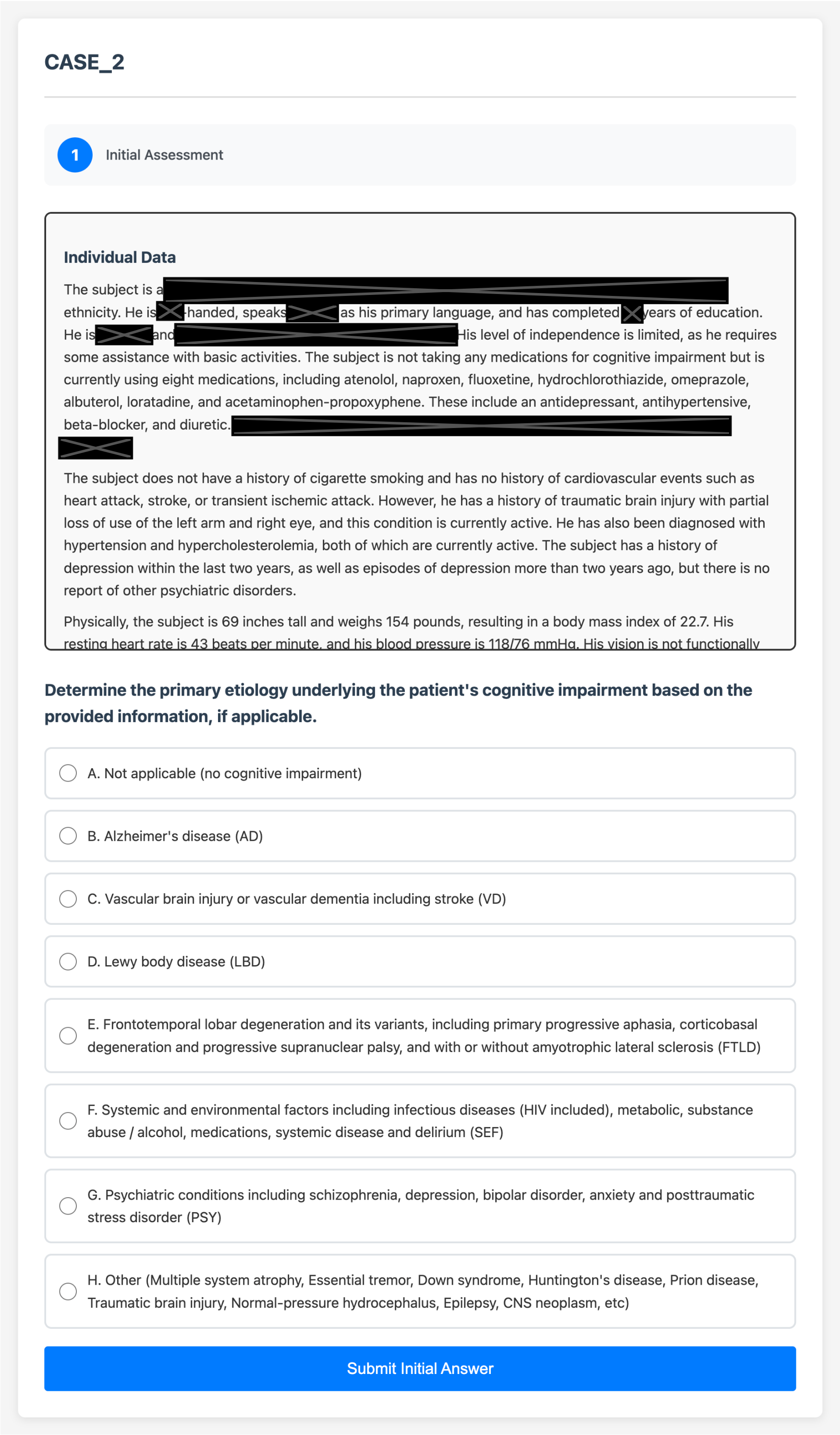


**Figure S16**: **Page one of an example sequential case presentation.** Clinicians are asked to review case information and provide their initial impression.

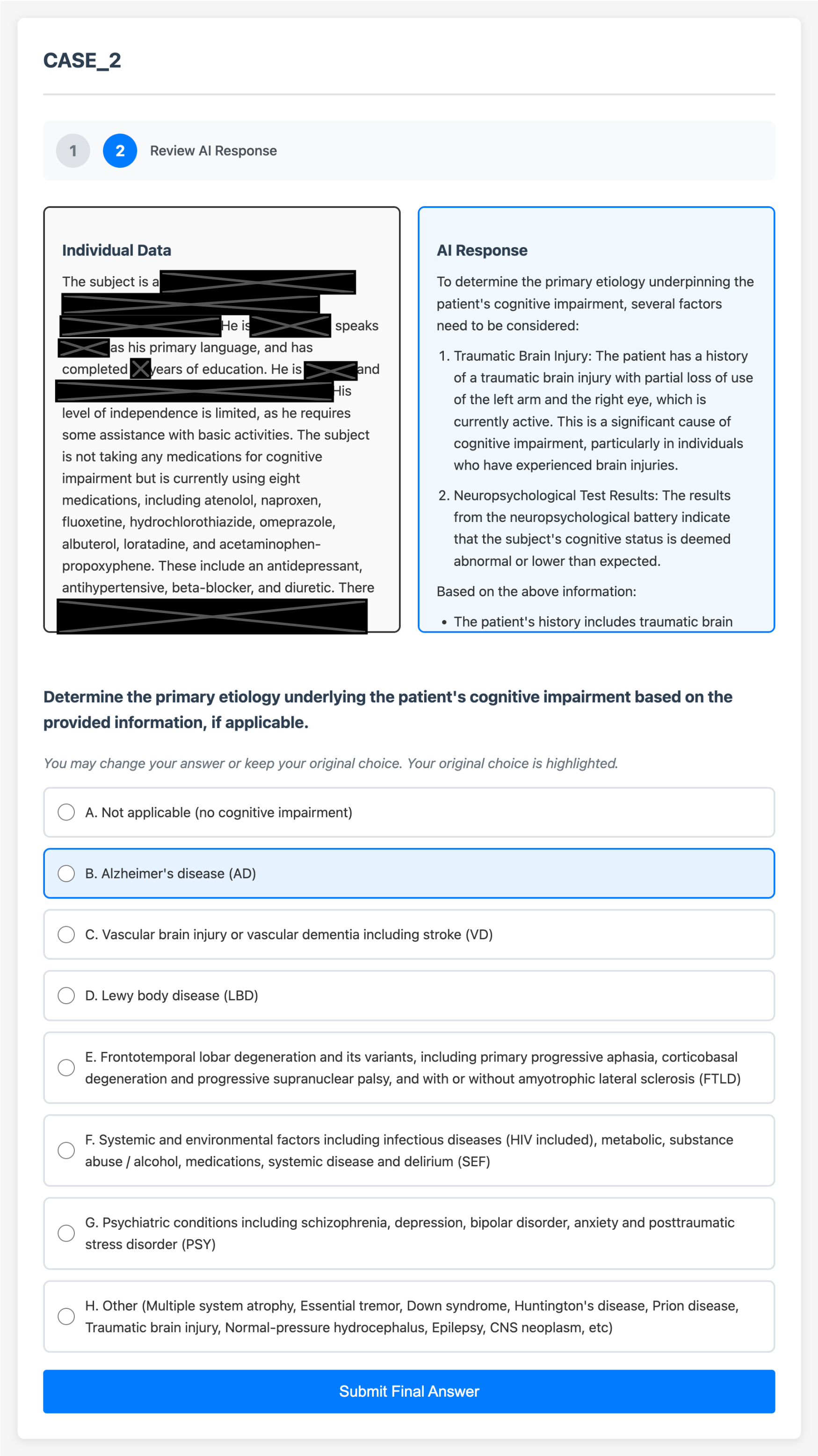


**Figure S17**: **Page two of an example sequential case presentation.** Clinicians are provided with the LLM response, and are provided the opportunity to revise their initial impression. They are still provided with the original case information, question, and answer choices. There is also an indicator showing their initial impression.

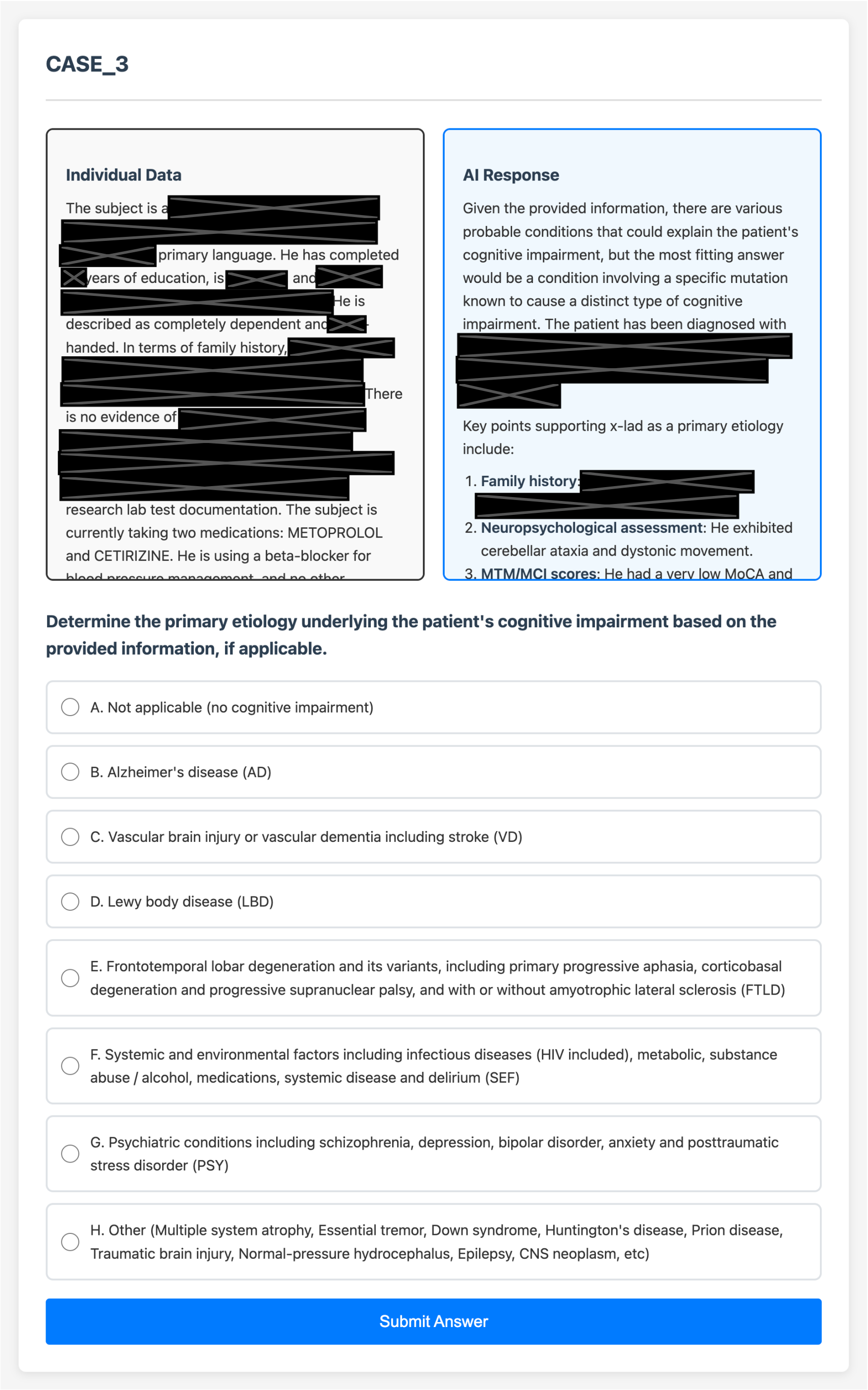


**Figure S18**: **Example of a concurrent case presentation.** Clinicians are provided with the case information alongside the LLM response and are asked to provide their impression.

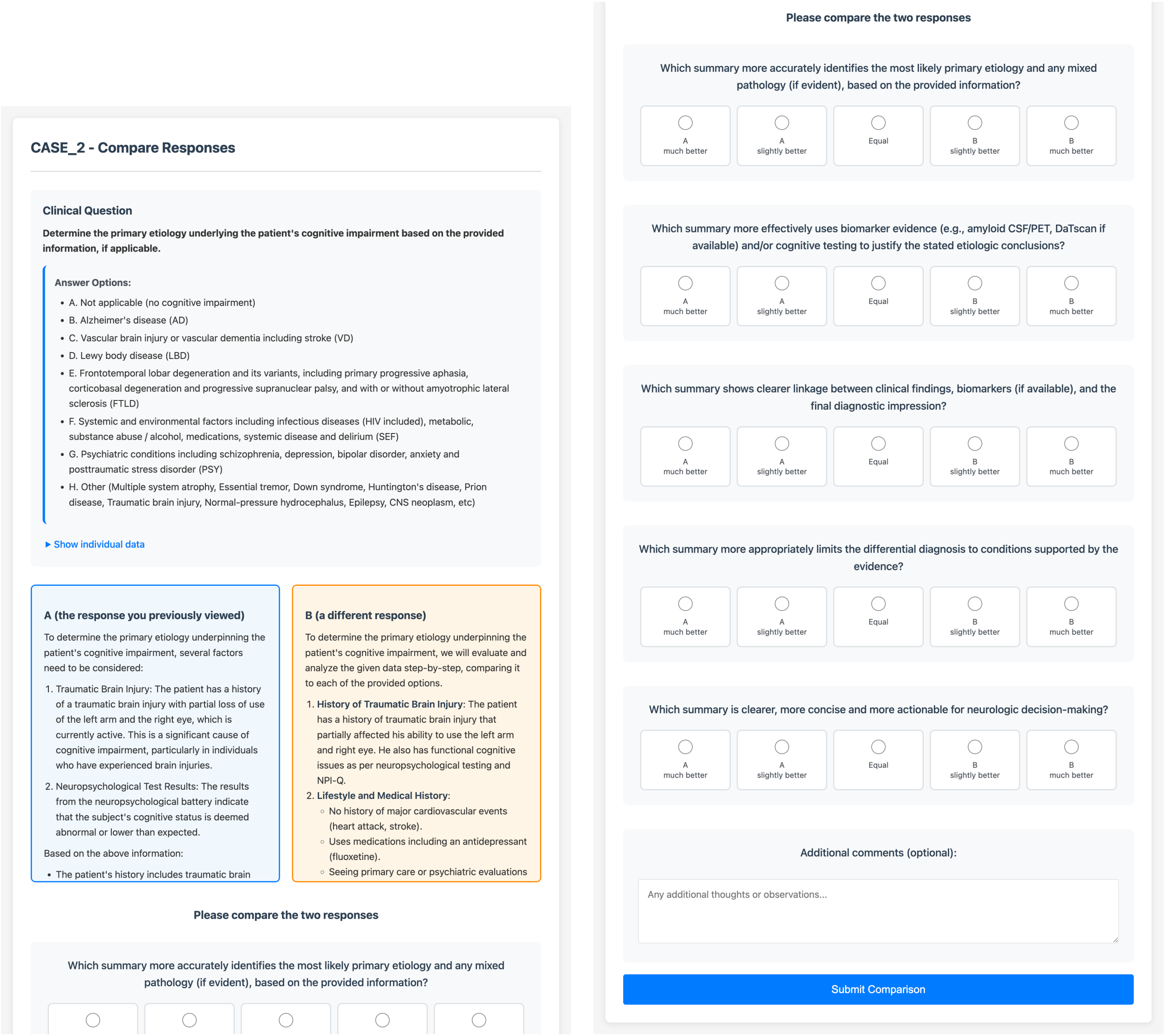


(a) (b)

**Figure S19**: **Example of comparison page.** Clinicians are asked to review the LLM response they previously reviewed as well as a different LLM response (baseline Q3B) and asked to compare the responses on 5 dimensions using a 5-point Likert scale. (A–B) Continuous scroll view of the interface.

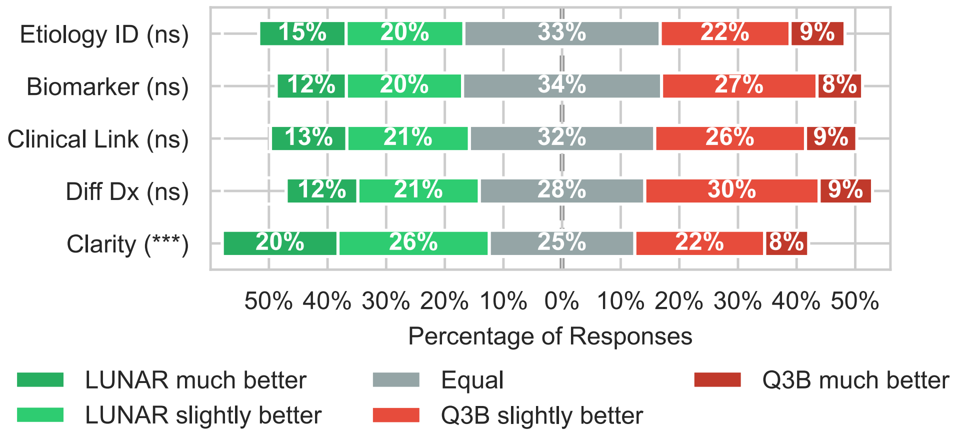


**Figure S20**: Diverging stacked bar chart of neurologists' comparative ratings of diagnostic quality across five dimensions (primary etiology identification, biomarker evidence usage, clinical-biomarker linkage, differential diagnosis appropriateness, and clarity/actionability). Ratings compare LUNAR against the baseline LLM on a 5-point Likert scale. Statistical significance of preference (departure from equal preference) is indicated in parentheses next to each dimension label and was assessed via two-sided Wilcoxon signed-rank tests on per-case median ratings. Asterisks denote: * $p < 0.05$, ** $p < 0.01$, *** $p < 0.001$, **** $p < 0.0001$.

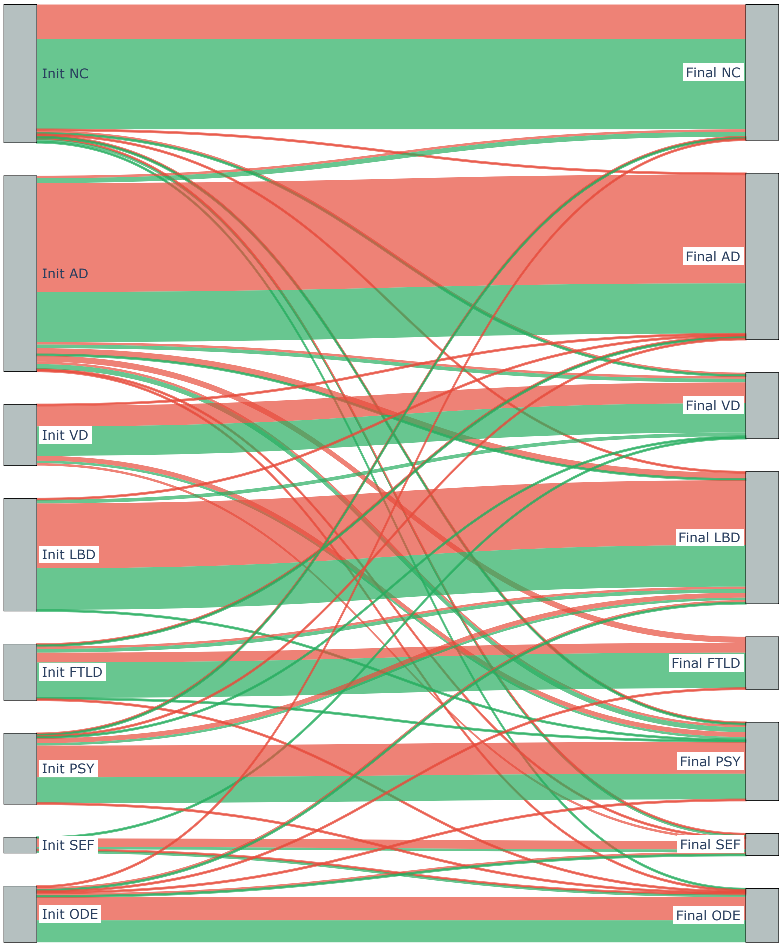


**Figure S21**: **Sankey diagram visualizing answer choice changes in the sequential arm.** $n = 551$ data points are represented, with color indicating the correctness of final responses. The left side indicates the initial answer choice, the right side indicates the final answer choice. Each band represents the aggregate number of responses transitioning between a given initial and final diagnostic category, with band width proportional to the number of responses. Green bands denote transitions yielding a correct final diagnosis; red bands denote transitions yielding an incorrect final diagnosis.

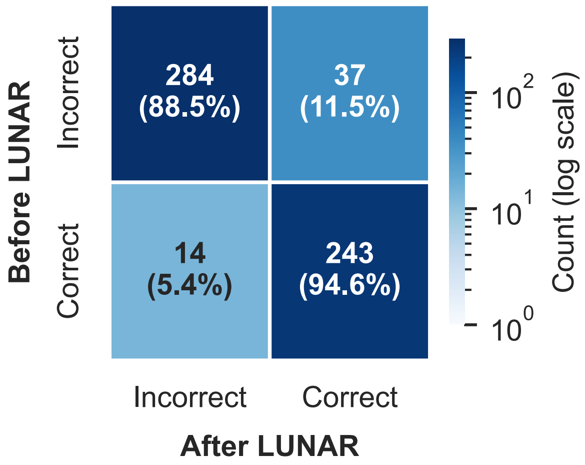


**Figure S22**: Transition matrix of answer correctness in the sequential arm before (Pre) and after (Post) exposure to LUNAR response. Cell values show counts with row percentages in parentheses. The proportion of cases that improved versus worsened in correctness differed significantly (McNemar's test, $\chi^2 = 8.82$, $p = 0.003$; odds ratio = 2.57).

**Table S1**: **Study population.** Demographic and clinical characteristics for five datasets (NACC, ADNI, BrainLat, NIFD and PPMI) totaling $54,535$ participants. Data from NACC $n = 17,847$ was used for training. A held out set from NACC ($n = 31,029$) was used as in-distribution testing set and the remaining cohorts were used as out-of-distribution test sets. The p-value for each dataset indicates the statistical significance of inter-group differences per column, based on one-way ANOVA for continuous variables and two-sided $\chi^2$ tests for categorical variables. Here, N.A. denotes data not available. The symbol ^ indicates that data was not available for some subjects.

| Dataset (group) | Age mean ± std | Female (percentage) | Education in years mean ± std | Race (White; Black; Asian; American Indian; Pacific; Multi-race) | CDR mean ± std |
|---|---|---|---|---|---|
| **NACC** | | | | | |
| NC [n = 18459] | 72.60 ± 11.45 | 12012, 65.07% | 15.88 ± 2.97^ | (13981, 2835, 596, 130, 12, 641)^ | 0.05 ± 0.16^ |
| MCI [n = 9323] | 75.45 ± 10.12 | 4959, 53.19% | 15.20 ± 3.41^ | (6893, 1558, 294, 71, 4, 341)^ | 0.46 ± 0.19^ |
| DE [n = 21094] | 75.82 ± 10.77 | 10969, 52.00% | 14.71 ± 3.66^ | (17608, 1951, 442, 105, 19, 531)^ | 1.60 ± 0.91 |
| *p*-value | <1.0e-200 | 4.554e-166 | <1.0e-200 | 2.631e-127 | <1.0e-200 |
| **ADNI** | | | | | |
| NC [n = 555] | 73.03 ± 7.10 | 321, 57.84% | 16.72 ± 2.42 | (464, 58, 17, 2, 0, 11)^ | 0.01 ± 0.08 |
| MCI [n = 603] | 73.05 ± 7.70 | 278, 46.10% | 16.19 ± 2.63 | (556, 29, 4, 1, 2, 6)^ | 0.53 ± 0.21 |
| DE [n = 234] | 75.46 ± 8.41 | 102, 43.59% | 15.64 ± 2.61 | (215, 8, 8, 0, 0, 3) | 1.11 ± 0.46^ |
| *p*-value | 5.369e-05 | 2.650e-05 | 1.487e-07 | 5.459e-05 | <1.0e-200 |
| **BrainLat** | | | | | |
| NC [n = 242] | 67.88 ± 8.88^ | 159, 65.70% | 14.72 ± 4.31^ | N.A. | N.A. |
| DE [n = 418] | 70.27 ± 9.79^ | 235, 56.22% | 12.25 ± 4.85^ | N.A. | N.A. |
| *p*-value | 3.803e-03 | 2.084e-02 | 1.288e-10 | N.A. | N.A. |
| **NIFD** | | | | | |
| NC [n = 136] | 66.40 ± 8.64 | 78, 57.35% | 17.32 ± 1.92^ | (109, 0, 0, 0, 0, 4)^ | 0.01 ± 0.08^ |
| DE [n = 156] | 65.37 ± 7.03 | 64, 41.03% | 15.84 ± 2.79^ | (144, 1, 0, 0, 0, 8)^ | 1.31 ± 0.81^ |
| *p*-value | 2.634e-01 | 7.650e-03 | 6.880e-07 | 5.524e-01 | 5.211e-38 |
| **PPMI** | | | | | |
| NC [n = 2757] | 64.01 ± 8.75 | 1294, 46.94% | 16.21 ± 2.90^ | (2574, 44, 28, 0, 0, 92)^ | N.A. |
| MCI [n = 495] | 67.30 ± 8.28 | 170, 34.34% | 15.58 ± 3.33 | (464, 8, 3, 0, 0, 17)^ | N.A. |
| DE [n = 63] | 67.76 ± 9.38 | 19, 30.16% | 15.83 ± 4.12 | (58, 1, 1, 0, 0, 3) | N.A. |
| *p*-value | 1.615e-15 | 9.065e-08 | 7.121e-05 | 9.674e-01 | N.A. |

**Table S2**: **Question types and class balancing in the NACC training set.** Overview of the seven diagnostic question types and their training options, with the number of unique cases per class before task-specific oversampling with replacement. Oversampling was applied independently per question type to match the majority class size ("Yes" indicates upsampling), ensuring balanced exposure to rare etiologies and pathologies during fine-tuning. The COG task was divided into three categories to prevent collapse to NC or DE predictions.

| **Question type** | **Unique count** | **Oversampled** |
|---|---|---|
| **Cognitive status (COG)** | | |
| Normal cognition (NC) | 1,300 | No |
| Mild cognitive impairment (MCI) | 1,300 | No |
| Normal cognition (NC) | 1,300 | No |
| Dementia (DE) | 1,300 | No |
| Mild cognitive impairment (MCI) | 1,300 | No |
| Dementia (DE) | 1,300 | No |
| **Primary etiology (ETPR)** | | |
| Alzheimer's disease (AD) | 1,000 | No |
| Frontotemporal lobar degeneration and variants (FTLD) | 1,000 | No |
| Lewy body disease (LBD) | 853 | Yes |
| Vascular brain injury or vascular dementia (VD) | 579 | Yes |
| Psychiatric conditions (PSY) | 176 | Yes |
| Systemic and environmental factors (SEF) | 327 | Yes |
| Other (Multiple system atrophy, Essential tremor, etc.) | 112 | Yes |
| Not applicable (no cognitive impairment) | 1,000 | No |
| **Amyloid CSF (CSF)** | | |
| No | 400 | No |
| Yes | 400 | No |
| **Amyloid PET (PET)** | | |
| No | 1,000 | No |
| Yes | 1,000 | No |
| **DaTscan (DAT)** | | |
| No | 100 | No |
| Yes | 100 | No |
| **Neuropathology (NP-ONE)** | | |
| Alzheimer's disease pathology (NP-AD) | 633 | No |
| Frontotemporal lobar degeneration with tau pathology or TDP-43 pathology (NP-FTLD) | 299 | Yes |
| Lewy body pathology (NP-LBD) | 97 | Yes |
| No listed option is correct | 82 | Yes |
| **Neuropathology (NP-MIXED)** | | |
| NP-AD and NP-LBD | 585 | Yes |
| NP-AD and NP-FTLD | 92 | Yes |
| NP-LBD and NP-FTLD | 58 | Yes |
| NP-AD, NP-LBD, and NP-FTLD | 71 | Yes |
| No listed option is correct | 83 | Yes |

**Table S3**: **Class distribution in the NACC held-out test set.** Number of unique cases per class across the diagnostic question types in the internal NACC test partition (n = 31,029 participants). No oversampling was applied to test sets; distributions reflect the natural prevalence in the cohort, highlighting imbalance particularly in etiological and neuropathological categories.

| **Question type** | **Unique count** |
|---|---|
| **Cognitive status (COG)** | |
| Normal cognition (NC) | 13,819 |
| Mild cognitive impairment (MCI) | 4,669 |
| Dementia (DE) | 12,541 |
| **Primary etiology (ETPR)** | |
| Alzheimer's disease (AD) | 13,099 |
| Frontotemporal lobar degeneration and variants (FTLD) | 1,258 |
| Lewy body disease (LBD) | 595 |
| Vascular brain injury or vascular dementia (VD) | 403 |
| Psychiatric conditions (PSY) | 205 |
| Systemic and environmental factors (SEF) | 240 |
| Other (Multiple system atrophy, Essential tremor, etc.) | 169 |
| Not applicable (no cognitive impairment) | 13,819 |
| **Amyloid CSF (CSF)** | |
| No | 109 |
| Yes | 179 |
| **Amyloid PET (PET)** | |
| No | 308 |
| Yes | 500 |
| **DaTscan (DAT)** | |
| No | 55 |
| Yes | 16 |
| **Neuropathology (NP-ONE)** | |
| Alzheimer's disease pathology (NP-AD) | 184 |
| Frontotemporal lobar degeneration with tau pathology or TDP-43 pathology (NP-FTLD) | 100 |
| Lewy body pathology (NP-LBD) | 29 |
| No listed option is correct | 63 |
| **Neuropathology (NP-MIXED)** | |
| NP-AD and NP-LBD | 206 |
| NP-AD and NP-FTLD | 52 |
| NP-LBD and NP-FTLD | 10 |
| NP-AD, NP-LBD, and NP-FTLD | 24 |
| No listed option is correct | 63 |

**Table S4**: **Class distribution in the ADNI external test set.** Number of unique cases per class across applicable diagnostic question types in the ADNI cohort (n = 1,392 participants). Distributions reflect the Alzheimer's disease neuroimaging focus, with no oversampling applied.

| Question type | Unique count |
|---|---|
| **Cognitive status (COG)** | |
| Normal cognition (NC) | 555 |
| Mild cognitive impairment (MCI) | 603 |
| Dementia (DE) | 234 |
| **Primary etiology (ETPR)** | |
| Alzheimer's disease (AD) | 504 |
| Not applicable (no cognitive impairment) | 555 |
| **Amyloid PET (PET)** | |
| No | 715 |
| Yes | 677 |
| **Amyloid CSF (CSF)** | |
| No | 130 |
| Yes | 276 |

**Table S5**: **Class distribution in the BrainLat external test set.** Number of unique cases per class across applicable diagnostic question types in the BrainLat cohort (n = 660 participants). Distributions reflect the Latin American dementia research focus, with no oversampling applied.

| Question type | Unique count |
|---|---|
| **Cognitive status (COG)** | |
| Normal cognition (NC) | 242 |
| Dementia (DE) | 418 |
| **Primary etiology (ETPR)** | |
| Alzheimer's disease (AD) | 269 |
| Frontotemporal lobar degeneration and variants (FTLD) | 149 |
| Not applicable (no cognitive impairment) | 242 |

**Table S6**: **Class distribution in the NIFD external test set.** Number of unique cases per class across applicable diagnostic question types in the NIFD cohort (n = 292 participants). Distributions reflect cohort-specific focus on frontotemporal lobar degeneration syndromes, with no oversampling applied.

| Question type | Unique count |
|---|---|
| **Cognitive status (COG)** | |
| Normal cognition (NC) | 136 |
| Dementia (DE) | 156 |
| **Primary etiology (ETPR)** | |
| Frontotemporal lobar degeneration and variants (FTLD) | 156 |
| Not applicable (no cognitive impairment) | 136 |

**Table S7**: **Class distribution in the PPMI external test set.** Number of unique cases per class across applicable diagnostic question types in the PPMI cohort (n = 3,315 participants). Distributions reflect the Parkinson's disease-focused design of the cohort, with no oversampling applied.

| **Question type** | **Unique count** |
|---|---|
| **Cognitive status (COG)** | |
| Normal cognition (NC) | 2757 |
| Mild cognitive impairment (MCI) | 495 |
| Dementia (DE) | 63 |
| **Primary etiology (ETPR)** | |
| Idiopathic Parkinson's disease Idiopathik PD) | 357 |
| Prodromal Parkinson's disease (Prodromal PD) | 117 |
| Other neurological disorder(s) (Other ND) | 36 |
| No Parkinson's disease nor other neurological disorder (No PD/ND) | 48 |
| **DaTscan (DAT)** | |
| No | 60 |
| Yes | 1216 |

**Table S8**: **Paraphrased question variants for diagnostic tasks.** Five linguistically varied versions were created for each question type to increase surface variation in training and evaluation. This was intended to reduce rote responses and promote explicit chain-of-thought reasoning while keeping semantics and reference answers identical across versions.

| Task type | Question variants |
|---|---|
| **COG** | - Using the information provided, determine the patient's cognitive status. |
| | - Identify the patient's cognitive status based on the given information. |
| | - Based on the provided clinical details, classify the patient's cognitive status. |
| | - From the information available, determine the correct cognitive status for the patient. |
| | - Analyze the patient's information and identify their cognitive status. |
| **ETPR** | - Identify the primary etiologic diagnosis of the patient using the information provided, if applicable. |
| | - Identify the primary etiology of the patient's cognitive impairment using the information provided, if applicable. |
| | - Based on the clinical details, determine the main cause of the patient's cognitive impairment, if applicable. |
| | - Using the information provided, identify the primary cause of the patient's cognitive decline, if applicable. |
| | - Determine the primary etiology underlying the patient's cognitive impairment based on the provided information, if applicable. |
| **PET** | - Is the patient likely to have elevated amyloid pathology in the brain given the provided information? |
| | - Does the patient likely have abnormal amyloid accumulation in the brain based on the clinical data? |
| | - Based on the available information, is the patient likely amyloid positive in the brain? |
| | - Does the evidence suggest the presence of elevated amyloid burden in the brain in this patient? |
| | - Is there a high likelihood of underlying amyloid pathology in the brain based on the patient's profile? |
| **CSF** | - Is the patient likely to have elevated amyloid pathology in the brain given the provided information? |
| | - Does the patient likely have abnormal amyloid accumulation in the brain based on the clinical data? |
| | - Based on the available information, is the patient likely amyloid positive in the brain? |
| | - Does the evidence suggest the presence of elevated amyloid burden in the brain in this patient? |
| | - Is there a high likelihood of underlying amyloid pathology in the brain based on the patient's profile? |
| **DAT** | - Is there evidence of Lewy body disease in the brain based on the provided information? |
| | - Does the provided information suggest the presence of Lewy body disease in the brain? |
| | - Is the patient likely to have Lewy body pathology in the brain based on the available data? |
| | - Based on the information given, is there evidence consistent with Lewy body disease in the brain? |
| | - Does the information support the presence of underlying Lewy body pathology in the brain? |
| **NP-ONE** | - Which of the following pathology is causing the patient's cognitive impairment based on the clinical information provided? |
| | - Using the provided clinical information, determine the pathological cause of the patient's cognitive decline from the options listed below. |
| | - Based on the clinical data, identify the neuropathology responsible for the patient's cognitive impairment by selecting from the given choices. |
| | - From the options below, select the disease pathology causing cognitive impairment as indicated by the clinical information. |
| | - Review the provided clinical information and choose the pathology underlying the patient's cognitive symptoms from the available options. |
| **NP-MIXED** | - Which of the following pathologies are causing the patient's cognitive impairment based on the clinical information provided? |
| | - Using the provided clinical information, determine the pathological causes of the patient's cognitive decline from the options listed below. |
| | - Based on the clinical data, identify the neuropathologies responsible for the patient's cognitive impairment by selecting from the given choices. |
| | - From the options below, select the disease pathologies causing cognitive impairment as indicated by the clinical information. |
| | - Review the provided clinical information and choose the pathologies underlying the patient's cognitive symptoms from the available options. |

**Table S9**: **Per-class performance on the COG task.** Precision, recall and F1-scores (bootstrap median and 95% confidence intervals) for normal cognition (NC), mild cognitive impairment (MCI) and dementia (DE) across models (Qwen2.5-3B-Instruct or Q3B, LUNAR-OS-SCe, LUNAR-OS, LUNAR-SCe, LUNAR (Ours) and Qwen2.5-7B-Instruct or Q7B). Results are shown for the internal NACC testing set and the combined external testing cohorts (ADNI, BrainLat, NIFD and PPMI). Best model per task is **bolded** and the second best one is <u>underlined</u>.

| Dataset | Class | Model | Precision | Recall | F1-score |
|---|---|---|---|---|---|
| Internal validation | NC | Q3B | 0.896 [0.893, 0.898] | 0.684 [0.680, 0.687] | 0.775 [0.773, 0.778] |
| | | LUNAR-OS-SCe | **0.933 [0.931, 0.935]** | 0.779 [0.776, 0.782] | 0.849 [0.847, 0.851] |
| | | LUNAR-OS | 0.927 [0.925, 0.929] | 0.804 [0.801, 0.806] | 0.861 [0.859, 0.863] |
| | | LUNAR-SCe | <u>0.931 [0.929, 0.933]</u> | 0.804 [0.801, 0.807] | <u>0.863 [0.861, 0.865]</u> |
| | | LUNAR | 0.926 [0.924, 0.929] | **0.816 [0.814, 0.819]** | **0.868 [0.866, 0.870]** |
| | | Q7B | 0.892 [0.890, 0.894] | <u>0.807 [0.804, 0.810]</u> | 0.848 [0.846, 0.850] |
| | MCI | Q3B | 0.277 [0.273, 0.281] | 0.695 [0.689, 0.700] | 0.396 [0.392, 0.401] |
| | | LUNAR-OS-SCe | 0.390 [0.385, 0.394] | **0.732 [0.727, 0.738]** | 0.509 [0.504, 0.513] |
| | | LUNAR-OS | 0.398 [0.393, 0.402] | <u>0.732 [0.726, 0.738]</u> | 0.515 [0.511, 0.520] |
| | | LUNAR-SCe | **0.431 [0.426, 0.435]** | 0.681 [0.675, 0.687] | <u>0.528 [0.523, 0.532]</u> |
| | | LUNAR | <u>0.428 [0.423, 0.433]</u> | 0.692 [0.686, 0.697] | **0.529 [0.524, 0.533]** |
| | | Q7B | 0.331 [0.327, 0.335] | 0.711 [0.705, 0.717] | 0.452 [0.447, 0.456] |
| | DE | Q3B | 0.893 [0.890, 0.896] | 0.595 [0.591, 0.599] | 0.714 [0.711, 0.717] |
| | | LUNAR-OS-SCe | 0.938 [0.936, 0.940] | 0.801 [0.798, 0.804] | 0.864 [0.862, 0.866] |
| | | LUNAR-OS | <u>0.946 [0.944, 0.948]</u> | 0.789 [0.786, 0.792] | 0.861 [0.858, 0.863] |
| | | LUNAR-SCe | 0.922 [0.920, 0.924] | **0.862 [0.859, 0.864]** | **0.891 [0.889, 0.893]** |
| | | LUNAR | 0.932 [0.930, 0.934] | <u>0.840 [0.837, 0.843]</u> | <u>0.884 [0.882, 0.886]</u> |
| | | Q7B | **0.966 [0.964, 0.967]** | 0.640 [0.636, 0.644] | 0.770 [0.767, 0.772] |
| External validation | NC | Q3B | 0.850 [0.843, 0.856] | 0.476 [0.469, 0.483] | 0.610 [0.603, 0.617] |
| | | LUNAR-OS-SCe | **0.893 [0.887, 0.898]** | 0.620 [0.613, 0.627] | 0.732 [0.727, 0.737] |
| | | LUNAR-OS | 0.877 [0.872, 0.882] | 0.738 [0.732, 0.744] | 0.802 [0.797, 0.806] |
| | | LUNAR-SCe | <u>0.889 [0.884, 0.894]</u> | 0.714 [0.708, 0.721] | 0.792 [0.787, 0.797] |
| | | LUNAR | 0.874 [0.869, 0.879] | **0.759 [0.753, 0.766]** | **0.813 [0.808, 0.817]** |
| | | Q7B | 0.882 [0.876, 0.886] | <u>0.744 [0.737, 0.750]</u> | <u>0.807 [0.802, 0.811]</u> |
| | MCI | Q3B | 0.249 [0.242, 0.256] | <u>0.672 [0.659, 0.684]</u> | 0.363 [0.354, 0.372] |
| | | LUNAR-OS-SCe | 0.313 [0.305, 0.321] | **0.716 [0.704, 0.728]** | 0.436 [0.427, 0.445] |
| | | LUNAR-OS | 0.347 [0.338, 0.356] | 0.644 [0.632, 0.656] | 0.451 [0.441, 0.460] |
| | | LUNAR-SCe | 0.352 [0.343, 0.361] | 0.655 [0.642, 0.666] | <u>0.458 [0.448, 0.468]</u> |
| | | LUNAR | <u>0.359 [0.349, 0.368]</u> | 0.620 [0.607, 0.633] | 0.454 [0.445, 0.464] |
| | | Q7B | **0.363 [0.353, 0.373]** | 0.658 [0.646, 0.671] | **0.468 [0.458, 0.478]** |
| | DE | Q3B | 0.664 [0.646, 0.680] | 0.453 [0.438, 0.468] | 0.539 [0.525, 0.553] |
| | | LUNAR-OS-SCe | 0.815 [0.801, 0.829] | 0.545 [0.530, 0.559] | 0.653 [0.639, 0.666] |
| | | LUNAR-OS | <u>0.859 [0.845, 0.872]</u> | 0.507 [0.493, 0.523] | 0.638 [0.625, 0.651] |
| | | LUNAR-SCe | 0.809 [0.796, 0.822] | **0.608 [0.593, 0.623]** | **0.694 [0.682, 0.706]** |
| | | LUNAR | 0.854 [0.841, 0.867] | <u>0.547 [0.533, 0.562]</u> | 0.667 [0.655, 0.679] |
| | | Q7B | **0.868 [0.855, 0.882]** | 0.544 [0.530, 0.560] | <u>0.669 [0.657, 0.682]</u> |

**Table S10**: **Macro-averaged performance on the ETPR task.** Macro precision, recall and F1-scores (bootstrap median and 95% confidence intervals) for the primary etiology task across models (Qwen2.5-3B-Instruct or Q3B, LUNAR-OS-SCe, LUNAR-OS, LUNAR-SCe, LUNAR (Ours) and Qwen2.5-7B-Instruct or Q7B). Results are shown for the internal NACC testing set and the external testing cohorts (ADNI, BrainLat, NIFD and PPMI). **Bold** values indicate the highest score and underlined values indicate the second highest score for each class–dataset combination.

| Dataset | Model | Macro Precision | Macro Recall | Macro F1 |
|---|---|---|---|---|
| NACC | Q3B | 0.229 [0.227, 0.231] | 0.280 [0.275, 0.285] | 0.208 [0.206, 0.210] |
| | LUNAR-OS-SCe | **0.330 [0.325, 0.336]** | 0.405 [0.400, 0.410] | **0.324 [0.320, 0.328]** |
| | LUNAR-OS | 0.323 [0.318, 0.328] | 0.409 [0.404, 0.414] | 0.323 [0.320, 0.327] |
| | LUNAR-SCe | 0.324 [0.322, 0.326] | **0.494 [0.486, 0.501]** | 0.316 [0.313, 0.318] |
| | LUNAR | 0.320 [0.317, 0.322] | 0.490 [0.483, 0.496] | 0.318 [0.315, 0.320] |
| | Q7B | 0.306 [0.303, 0.309] | 0.370 [0.364, 0.375] | 0.284 [0.280, 0.287] |
| ADNI | Q3B | 0.731 [0.717, 0.745] | 0.508 [0.496, 0.520] | 0.539 [0.525, 0.553] |
| | LUNAR-OS-SCe | 0.811 [0.800, 0.822] | 0.739 [0.727, 0.750] | 0.773 [0.762, 0.783] |
| | LUNAR-OS | 0.815 [0.804, 0.826] | **0.756 [0.744, 0.768]** | **0.783 [0.772, 0.794]** |
| | LUNAR-SCe | 0.848 [0.836, 0.859] | 0.659 [0.645, 0.672] | 0.739 [0.727, 0.751] |
| | LUNAR | **0.855 [0.844, 0.866]** | 0.642 [0.629, 0.655] | 0.731 [0.720, 0.743] |
| | Q7B | 0.791 [0.780, 0.803] | 0.665 [0.652, 0.677] | 0.705 [0.692, 0.717] |
| BrainLat | Q3B | 0.471 [0.447, 0.494] | 0.338 [0.323, 0.354] | 0.337 [0.322, 0.353] |
| | LUNAR-OS-SCe | 0.536 [0.520, 0.551] | 0.427 [0.411, 0.443] | 0.439 [0.423, 0.455] |
| | LUNAR-OS | 0.544 [0.527, 0.560] | **0.449 [0.431, 0.466]** | **0.463 [0.445, 0.480]** |
| | LUNAR-SCe | 0.548 [0.530, 0.566] | 0.347 [0.330, 0.362] | 0.386 [0.368, 0.403] |
| | LUNAR | **0.556 [0.539, 0.573]** | 0.382 [0.365, 0.399] | 0.404 [0.386, 0.422] |
| | Q7B | 0.526 [0.506, 0.544] | 0.373 [0.357, 0.389] | 0.375 [0.358, 0.391] |
| NIFD | Q3B | 0.764 [0.722, 0.797] | 0.241 [0.221, 0.264] | 0.366 [0.340, 0.394] |
| | LUNAR-OS-SCe | 0.872 [0.840, 0.904] | 0.388 [0.365, 0.411] | 0.504 [0.478, 0.527] |
| | LUNAR-OS | 0.876 [0.844, 0.906] | 0.355 [0.335, 0.378] | 0.476 [0.453, 0.501] |
| | LUNAR-SCe | 0.912 [0.888, 0.934] | 0.404 [0.380, 0.430] | 0.550 [0.524, 0.576] |
| | LUNAR | **0.915 [0.892, 0.936]** | **0.419 [0.393, 0.445]** | **0.570 [0.542, 0.595]** |
| | Q7B | 0.862 [0.831, 0.889] | 0.406 [0.380, 0.431] | 0.538 [0.511, 0.563] |
| PPMI | Q3B | 0.350 [0.334, 0.367] | 0.365 [0.342, 0.389] | 0.340 [0.322, 0.359] |
| | LUNAR-OS-SCe | 0.362 [0.337, 0.392] | 0.409 [0.393, 0.428] | 0.351 [0.336, 0.368] |
| | LUNAR-OS | **0.377 [0.352, 0.406]** | **0.422 [0.404, 0.439]** | 0.348 [0.332, 0.366] |
| | LUNAR-SCe | 0.374 [0.353, 0.396] | 0.421 [0.402, 0.440] | **0.357 [0.339, 0.375]** |
| | LUNAR | 0.351 [0.331, 0.374] | 0.411 [0.391, 0.430] | 0.334 [0.319, 0.351] |
| | Q7B | 0.359 [0.343, 0.379] | 0.362 [0.342, 0.385] | 0.353 [0.335, 0.374] |

**Table S11**: **Per-class performance on the ETPR task for the NACC cohort.** Precision, recall and F1-scores (bootstrap median and 95% confidence intervals) for different classes across models. Best model per task is **bolded** and the second best one is underlined.

| Dataset | Class | Model | Precision | Recall | F1-score |
|---|---|---|---|---|---|
| NACC | NC | Q3B | 0.855 [0.851, 0.859] | 0.363 [0.359, 0.366] | 0.509 [0.505, 0.513] |
| | | LUNAR-OS-SCe | 0.888 [0.885, 0.890] | **0.864 [0.861, 0.866]** | **0.876 [0.874, 0.877]** |
| | | LUNAR-OS | 0.899 [0.897, 0.901] | 0.851 [0.848, 0.853] | 0.874 [0.872, 0.876] |
| | | LUNAR-SCe | 0.922 [0.920, 0.924] | 0.780 [0.777, 0.783] | 0.845 [0.843, 0.847] |
| | | LUNAR | 0.921 [0.919, 0.923] | 0.781 [0.778, 0.784] | 0.845 [0.843, 0.847] |
| | | Q7B | **0.929 [0.926, 0.931]** | 0.598 [0.594, 0.602] | 0.727 [0.725, 0.730] |
| | AD | Q3B | 0.586 [0.583, 0.590] | 0.631 [0.627, 0.635] | 0.608 [0.605, 0.611] |
| | | LUNAR-OS-SCe | 0.842 [0.839, 0.845] | **0.693 [0.689, 0.697]** | **0.760 [0.757, 0.763]** |
| | | LUNAR-OS | 0.836 [0.833, 0.839] | 0.693 [0.689, 0.696] | 0.758 [0.755, 0.760] |
| | | LUNAR-SCe | **0.899 [0.896, 0.902]** | 0.488 [0.484, 0.492] | 0.633 [0.629, 0.636] |
| | | LUNAR | 0.895 [0.892, 0.898] | 0.482 [0.478, 0.486] | 0.627 [0.623, 0.630] |
| | | Q7B | 0.715 [0.712, 0.719] | 0.677 [0.673, 0.680] | 0.696 [0.693, 0.698] |
| | FTLD | Q3B | 0.185 [0.177, 0.192] | 0.282 [0.272, 0.294] | 0.223 [0.215, 0.232] |
| | | LUNAR-OS-SCe | 0.411 [0.397, 0.426] | 0.270 [0.260, 0.282] | 0.326 [0.315, 0.338] |
| | | LUNAR-OS | 0.372 [0.358, 0.385] | 0.285 [0.274, 0.296] | 0.323 [0.311, 0.334] |
| | | LUNAR-SCe | 0.337 [0.326, 0.349] | 0.332 [0.320, 0.343] | **0.335 [0.324, 0.345]** |
| | | LUNAR | 0.281 [0.271, 0.291] | **0.403 [0.392, 0.415]** | 0.332 [0.321, 0.341] |
| | | Q7B | **0.426 [0.410, 0.442]** | 0.250 [0.239, 0.261] | 0.315 [0.303, 0.328] |
| | LBD | Q3B | 0.104 [0.097, 0.111] | 0.239 [0.223, 0.254] | 0.145 [0.135, 0.154] |
| | | LUNAR-OS-SCe | 0.189 [0.181, 0.196] | 0.713 [0.697, 0.730] | 0.298 [0.288, 0.308] |
| | | LUNAR-OS | 0.175 [0.169, 0.182] | 0.736 [0.721, 0.751] | 0.283 [0.274, 0.293] |
| | | LUNAR-SCe | 0.206 [0.198, 0.214] | **0.744 [0.729, 0.759]** | 0.323 [0.313, 0.333] |
| | | LUNAR | **0.236 [0.228, 0.244]** | 0.728 [0.713, 0.744] | **0.356 [0.346, 0.367]** |
| | | Q7B | 0.208 [0.196, 0.219] | 0.315 [0.299, 0.330] | 0.250 [0.238, 0.263] |
| | VD | Q3B | 0.056 [0.053, 0.059] | 0.495 [0.473, 0.516] | 0.101 [0.095, 0.106] |
| | | LUNAR-OS-SCe | 0.116 [0.109, 0.122] | 0.567 [0.545, 0.588] | 0.192 [0.183, 0.202] |
| | | LUNAR-OS | **0.120 [0.114, 0.128]** | 0.545 [0.523, 0.567] | **0.197 [0.188, 0.208]** |
| | | LUNAR-SCe | 0.109 [0.103, 0.115] | 0.636 [0.615, 0.657] | 0.186 [0.177, 0.195] |
| | | LUNAR | 0.110 [0.104, 0.115] | 0.630 [0.609, 0.651] | 0.187 [0.178, 0.196] |
| | | Q7B | 0.063 [0.060, 0.066] | **0.697 [0.676, 0.717]** | 0.116 [0.110, 0.121] |
| | SEF | Q3B | 0.013 [0.010, 0.016] | 0.065 [0.052, 0.079] | 0.022 [0.017, 0.027] |
| | | LUNAR-OS-SCe | 0.031 [0.020, 0.043] | 0.025 [0.016, 0.034] | 0.028 [0.018, 0.038] |
| | | LUNAR-OS | 0.027 [0.014, 0.039] | 0.014 [0.008, 0.021] | 0.018 [0.010, 0.028] |
| | | LUNAR-SCe | **0.034 [0.030, 0.039]** | **0.159 [0.140, 0.181]** | **0.056 [0.049, 0.064]** |
| | | LUNAR | 0.030 [0.026, 0.035] | 0.145 [0.126, 0.166] | 0.050 [0.043, 0.057] |
| | | Q7B | 0.018 [0.014, 0.022] | 0.060 [0.046, 0.074] | 0.028 [0.021, 0.034] |
| | PSY | Q3B | 0.019 [0.015, 0.023] | 0.085 [0.068, 0.102] | 0.031 [0.025, 0.038] |
| | | LUNAR-OS-SCe | 0.056 [0.044, 0.068] | 0.074 [0.059, 0.089] | 0.064 [0.051, 0.077] |
| | | LUNAR-OS | **0.059 [0.049, 0.071]** | 0.109 [0.090, 0.130] | **0.076 [0.064, 0.092]** |
| | | LUNAR-SCe | 0.034 [0.032, 0.037] | **0.531 [0.502, 0.563]** | 0.064 [0.059, 0.070] |
| | | LUNAR | 0.034 [0.031, 0.037] | 0.523 [0.493, 0.552] | 0.064 [0.059, 0.069] |
| | | Q7B | 0.043 [0.038, 0.049] | 0.236 [0.212, 0.263] | 0.073 [0.065, 0.082] |
| | Other | Q3B | 0.015 [0.012, 0.019] | 0.080 [0.062, 0.099] | 0.026 [0.020, 0.032] |
| | | LUNAR-OS-SCe | **0.110 [0.073, 0.150]** | 0.033 [0.022, 0.045] | 0.051 [0.033, 0.068] |
| | | LUNAR-OS | 0.093 [0.065, 0.125] | 0.040 [0.028, 0.054] | 0.056 [0.040, 0.075] |
| | | LUNAR-SCe | 0.049 [0.043, 0.055] | **0.278 [0.248, 0.309]** | **0.083 [0.073, 0.093]** |
| | | LUNAR | 0.049 [0.042, 0.055] | 0.226 [0.198, 0.252] | 0.081 [0.070, 0.091] |
| | | Q7B | 0.043 [0.035, 0.051] | 0.123 [0.103, 0.147] | 0.064 [0.053, 0.075] |

**Table S12**: **Per-class performance on the ETPR task for the ADNI cohort.** Precision, recall and F1-scores (bootstrap median and 95% confidence intervals) for different classes across models. Best model per task is **bolded** and the second best one is <u>underlined</u>.

| Dataset | Class | Model | Precision | Recall | F1-score |
|---|---|---|---|---|---|
| ADNI | NC | Q3B | 0.864 [0.840, 0.885] | 0.273 [0.257, 0.290] | 0.415 [0.395, 0.435] |
| | | LUNAR-OS-SCe | 0.847 [0.832, 0.862] | 0.730 [0.714, 0.745] | 0.784 [0.772, 0.796] |
| | | LUNAR-OS | 0.855 [0.841, 0.869] | <u>0.736 [0.720, 0.752]</u> | <u>0.791 [0.778, 0.804]</u> |
| | | LUNAR-SCe | 0.860 [0.847, 0.874] | **0.740 [0.723, 0.756]** | **0.795 [0.783, 0.807]** |
| | | LUNAR | <u>0.865 [0.851, 0.878]</u> | 0.722 [0.706, 0.740] | 0.787 [0.774, 0.799] |
| | | Q7B | **0.887 [0.870, 0.902]** | 0.538 [0.520, 0.556] | 0.669 [0.653, 0.685] |
| | AD | Q3B | 0.599 [0.581, 0.616] | 0.742 [0.724, 0.760] | 0.663 [0.648, 0.677] |
| | | LUNAR-OS-SCe | 0.774 [0.758, 0.790] | 0.748 [0.730, 0.764] | <u>0.761 [0.748, 0.774]</u> |
| | | LUNAR-OS | 0.776 [0.760, 0.793] | <u>0.775 [0.760, 0.791]</u> | **0.776 [0.763, 0.789]** |
| | | LUNAR-SCe | <u>0.837 [0.818, 0.854]</u> | 0.577 [0.557, 0.596] | 0.683 [0.665, 0.699] |
| | | LUNAR | **0.846 [0.827, 0.863]** | 0.562 [0.542, 0.582] | 0.675 [0.658, 0.692] |
| | | Q7B | 0.697 [0.680, 0.714] | **0.792 [0.775, 0.807]** | 0.741 [0.729, 0.755] |

**Table S13**: **Per-class performance on the ETPR task for the BrainLat cohort.** Precision, recall and F1-scores (bootstrap median and 95% confidence intervals) for different classes across models. Best model per task is **bolded** and the second best one is <u>underlined</u>.

| Dataset | Class | Model | Precision | Recall | F1-score |
|---|---|---|---|---|---|
| BrainLat | NC | Q3B | 0.641 [0.576, 0.698] | 0.115 [0.097, 0.133] | 0.195 [0.167, 0.223] |
| | | LUNAR-OS-SCe | **0.847 [0.815, 0.879]** | <u>0.331 [0.307, 0.357]</u> | <u>0.476 [0.449, 0.505]</u> |
| | | LUNAR-OS | 0.835 [0.804, 0.868] | **0.371 [0.345, 0.399]** | **0.514 [0.486, 0.543]** |
| | | LUNAR-SCe | <u>0.846 [0.807, 0.882]</u> | 0.267 [0.243, 0.292] | 0.405 [0.376, 0.435] |
| | | LUNAR | 0.844 [0.809, 0.875] | 0.291 [0.264, 0.317] | 0.432 [0.402, 0.463] |
| | | Q7B | 0.838 [0.790, 0.883] | 0.189 [0.167, 0.210] | 0.308 [0.276, 0.338] |
| | AD | Q3B | 0.532 [0.503, 0.558] | 0.509 [0.484, 0.537] | 0.520 [0.498, 0.543] |
| | | LUNAR-OS-SCe | 0.516 [0.492, 0.542] | <u>0.587 [0.561, 0.613]</u> | <u>0.549 [0.528, 0.570]</u> |
| | | LUNAR-OS | 0.539 [0.514, 0.564] | **0.619 [0.595, 0.646]** | **0.576 [0.554, 0.597]** |
| | | LUNAR-SCe | <u>0.556 [0.525, 0.586]</u> | 0.378 [0.353, 0.403] | 0.450 [0.424, 0.475] |
| | | LUNAR | **0.577 [0.544, 0.610]** | 0.368 [0.343, 0.395] | 0.450 [0.424, 0.476] |
| | | Q7B | 0.503 [0.476, 0.530] | 0.548 [0.521, 0.576] | 0.525 [0.501, 0.548] |
| | FTLD | Q3B | 0.240 [0.218, 0.265] | 0.390 [0.356, 0.423] | 0.297 [0.273, 0.323] |
| | | LUNAR-OS-SCe | 0.244 [0.219, 0.270] | 0.361 [0.328, 0.397] | 0.292 [0.264, 0.318] |
| | | LUNAR-OS | **0.256 [0.230, 0.286]** | 0.357 [0.321, 0.391] | 0.298 [0.271, 0.327] |
| | | LUNAR-SCe | 0.244 [0.219, 0.268] | <u>0.396 [0.362, 0.429]</u> | <u>0.301 [0.274, 0.328]</u> |
| | | LUNAR | <u>0.248 [0.226, 0.270]</u> | **0.487 [0.450, 0.520]** | **0.329 [0.303, 0.352]** |
| | | Q7B | 0.236 [0.214, 0.260] | 0.383 [0.350, 0.416] | 0.292 [0.267, 0.318] |

**Table S14**: **Per-class performance on the ETPR task for the NIFD cohort.** Precision, recall and F1-scores (bootstrap median and 95% confidence intervals) for different classes across models. Best model per task is **bolded** and the second best one is underlined.

| Dataset | Class | Model | Precision | Recall | F1-score |
|---|---|---|---|---|---|
| NIFD | NC | Q3B | 0.864 [0.818, 0.908] | 0.278 [0.246, 0.311] | 0.420 [0.380, 0.459] |
| | | LUNAR-OS-SCe | 0.903 [0.876, 0.929] | **0.607 [0.571, 0.645]** | **0.725 [0.696, 0.755]** |
| | | LUNAR-OS | 0.899 [0.870, 0.925] | 0.552 [0.515, 0.589] | 0.684 [0.650, 0.715] |
| | | LUNAR-SCe | 0.932 [0.907, 0.955] | 0.526 [0.487, 0.563] | 0.672 [0.637, 0.703] |
| | | LUNAR | **0.952 [0.929, 0.971]** | 0.526 [0.486, 0.566] | 0.677 [0.643, 0.712] |
| | | Q7B | 0.908 [0.879, 0.934] | 0.563 [0.524, 0.601] | 0.695 [0.664, 0.726] |
| | FTLD | Q3B | 0.662 [0.600, 0.721] | 0.205 [0.178, 0.235] | 0.313 [0.278, 0.352] |
| | | LUNAR-OS-SCe | 0.842 [0.780, 0.895] | 0.169 [0.144, 0.192] | 0.282 [0.245, 0.315] |
| | | LUNAR-OS | 0.854 [0.800, 0.908] | 0.159 [0.134, 0.184] | 0.267 [0.231, 0.303] |
| | | LUNAR-SCe | **0.890 [0.849, 0.929]** | 0.282 [0.253, 0.313] | 0.429 [0.391, 0.466] |
| | | LUNAR | 0.878 [0.838, 0.915] | **0.314 [0.283, 0.347]** | **0.462 [0.425, 0.500]** |
| | | Q7B | 0.818 [0.760, 0.863] | 0.248 [0.218, 0.280] | 0.381 [0.341, 0.419] |

**Table S15**: **Per-class performance on the ETPR task for the PPMI cohort.** Precision, recall and F1-scores (bootstrap median and 95% confidence intervals) for different classes across models. Best model per task is **bolded** and the second best one is <u>underlined</u>.

| Dataset | Class | Model | Precision | Recall | F1-score |
|---|---|---|---|---|---|
| PPMI | Idiopathic PD | Q3B | 0.845 [0.825, 0.864] | 0.697 [0.675, 0.719] | 0.764 [0.747, 0.780] |
| | | LUNAR-OS-SCe | 0.901 [0.887, 0.915] | 0.799 [0.780, 0.816] | 0.847 [0.834, 0.859] |
| | | LUNAR-OS | 0.877 [0.862, 0.892] | **0.845 [0.829, 0.863]** | 0.861 [0.849, 0.873] |
| | | LUNAR-SCe | **0.926 [0.913, 0.938]** | <u>0.839 [0.821, 0.858]</u> | **0.880 [0.869, 0.892]** |
| | | LUNAR | <u>0.914 [0.901, 0.927]</u> | 0.823 [0.804, 0.841] | <u>0.866 [0.853, 0.879]</u> |
| | | Q7B | 0.881 [0.865, 0.896] | 0.689 [0.665, 0.710] | 0.773 [0.757, 0.789] |
| | Prodromal PD | Q3B | <u>0.292 [0.250, 0.338]</u> | <u>0.206 [0.175, 0.242]</u> | <u>0.242 [0.206, 0.279]</u> |
| | | LUNAR-OS-SCe | 0.256 [0.218, 0.300] | 0.191 [0.158, 0.222] | 0.219 [0.185, 0.254] |
| | | LUNAR-OS | **0.324 [0.262, 0.386]** | 0.110 [0.086, 0.138] | 0.164 [0.131, 0.202] |
| | | LUNAR-SCe | 0.252 [0.196, 0.306] | 0.102 [0.077, 0.128] | 0.145 [0.110, 0.179] |
| | | LUNAR | 0.213 [0.151, 0.287] | 0.057 [0.039, 0.076] | 0.090 [0.062, 0.119] |
| | | Q7B | 0.235 [0.208, 0.263] | **0.349 [0.311, 0.387]** | **0.280 [0.252, 0.311]** |
| | No PD/ND | Q3B | 0.162 [0.127, 0.194] | 0.307 [0.248, 0.367] | 0.212 [0.167, 0.251] |
| | | LUNAR-OS-SCe | 0.205 [0.178, 0.236] | 0.633 [0.573, 0.695] | <u>0.310 [0.273, 0.347]</u> |
| | | LUNAR-OS | <u>0.207 [0.177, 0.236]</u> | **0.697 [0.638, 0.754]** | **0.319 [0.279, 0.355]** |
| | | LUNAR-SCe | 0.194 [0.168, 0.222] | 0.660 [0.599, 0.717] | 0.299 [0.264, 0.337] |
| | | LUNAR | 0.192 [0.166, 0.219] | <u>0.681 [0.622, 0.738]</u> | 0.300 [0.263, 0.334] |
| | | Q7B | **0.208 [0.168, 0.253]** | 0.296 [0.243, 0.352] | 0.244 [0.200, 0.292] |
| | Other ND | Q3B | 0.101 [0.073, 0.129] | **0.250 [0.188, 0.321]** | **0.143 [0.105, 0.180]** |
| | | LUNAR-OS-SCe | 0.083 [0.000, 0.194] | 0.016 [0.000, 0.039] | 0.028 [0.000, 0.065] |
| | | LUNAR-OS | 0.100 [0.033, 0.185] | 0.033 [0.011, 0.061] | 0.049 [0.017, 0.091] |
| | | LUNAR-SCe | **0.123 [0.071, 0.189]** | 0.082 [0.047, 0.124] | 0.098 [0.057, 0.146] |
| | | LUNAR | 0.083 [0.044, 0.126] | 0.083 [0.042, 0.124] | 0.083 [0.044, 0.124] |
| | | Q7B | <u>0.112 [0.068, 0.162]</u> | <u>0.115 [0.073, 0.167]</u> | <u>0.113 [0.072, 0.162]</u> |

**Table S16**: Amyloid PET positivity prediction. Median [95% CI] from 1,000 bootstrap resamples. Best model is bolded, second best is underlined.

| Cohort | Model | Macro F1 | Macro Precision | Macro Recall |
|---|---|---|---|---|
| NACC | Q3B | 0.631 [0.617, 0.647] | 0.661 [0.647, 0.675] | 0.664 [0.651, 0.678] |
| | LUNAR-OS-SCE | 0.732 [0.718, 0.746] | 0.736 [0.722, 0.749] | 0.750 [0.736, 0.763] |
| | LUNAR-OS | 0.737 [0.724, 0.751] | 0.742 [0.729, 0.754] | 0.756 [0.743, 0.769] |
| | LUNAR-SCE | **0.745 [0.732, 0.759]** | **0.746 [0.734, 0.759]** | **0.760 [0.747, 0.774]** |
| | LUNAR | 0.738 [0.725, 0.751] | 0.743 [0.730, 0.756] | 0.757 [0.744, 0.770] |
| | Q7B | 0.648 [0.633, 0.662] | 0.704 [0.691, 0.717] | 0.696 [0.683, 0.708] |
| ADNI | Q3B | 0.629 [0.617, 0.640] | 0.632 [0.620, 0.643] | 0.631 [0.619, 0.642] |
| | LUNAR-OS-SCE | 0.660 [0.649, 0.671] | 0.662 [0.651, 0.673] | 0.662 [0.650, 0.673] |
| | LUNAR-OS | 0.662 [0.652, 0.675] | 0.662 [0.652, 0.675] | 0.662 [0.653, 0.675] |
| | LUNAR-SCE | 0.664 [0.654, 0.676] | 0.665 [0.655, 0.677] | 0.664 [0.655, 0.676] |
| | LUNAR | **0.665 [0.655, 0.677]** | **0.669 [0.658, 0.680]** | **0.667 [0.657, 0.679]** |
| | Q7B | 0.614 [0.603, 0.627] | 0.641 [0.630, 0.654] | 0.623 [0.614, 0.635] |

**Table S17**: Amyloid CSF positivity prediction. Median [95% CI] from 1,000 bootstrap resamples. Best model is bolded, second best is underlined.

| Cohort | Model | Macro F1 | Macro Precision | Macro Recall |
|---|---|---|---|---|
| NACC | Q3B | 0.581 [0.554, 0.605] | 0.628 [0.605, 0.652] | 0.625 [0.602, 0.648] |
| | LUNAR-OS-SCE | 0.689 [0.664, 0.713] | 0.692 [0.668, 0.715] | 0.704 [0.679, 0.729] |
| | LUNAR-OS | 0.700 [0.674, 0.721] | 0.703 [0.677, 0.723] | 0.715 [0.689, 0.738] |
| | LUNAR-SCE | **0.729 [0.705, 0.751]** | **0.727 [0.704, 0.749]** | **0.739 [0.716, 0.762]** |
| | LUNAR | <u>0.701 [0.677, 0.724]</u> | <u>0.704 [0.680, 0.726]</u> | <u>0.716 [0.693, 0.738]</u> |
| | Q7B | 0.610 [0.585, 0.635] | 0.672 [0.649, 0.693] | 0.661 [0.639, 0.682] |
| ADNI | Q3B | 0.627 [0.607, 0.649] | 0.629 [0.610, 0.651] | 0.626 [0.605, 0.647] |
| | LUNAR-OS-SCE | **0.669 [0.647, 0.690]** | **0.667 [0.645, 0.688]** | **0.672 [0.650, 0.692]** |
| | LUNAR-OS | <u>0.664 [0.642, 0.684]</u> | <u>0.660 [0.639, 0.680]</u> | <u>0.671 [0.649, 0.693]</u> |
| | LUNAR-SCE | 0.659 [0.637, 0.680] | 0.659 [0.636, 0.680] | 0.660 [0.638, 0.682] |
| | LUNAR | 0.651 [0.629, 0.673] | 0.652 [0.629, 0.674] | 0.650 [0.628, 0.672] |
| | Q7B | 0.564 [0.541, 0.585] | 0.597 [0.576, 0.617] | 0.609 [0.586, 0.631] |

**Table S18**: DAT positivity prediction. Median [95% CI] from 1,000 bootstrap resamples. Best model is bolded, second best is underlined.

| Cohort | Model | Macro F1 | Macro Precision | Macro Recall |
|---|---|---|---|---|
| NACC | Q3B | 0.528 [0.475, 0.586] | 0.549 [0.481, 0.619] | 0.531 [0.488, 0.580] |
| | LUNAR-OS-SCE | 0.667 [0.611, 0.720] | 0.662 [0.607, 0.716] | **0.675 [0.617, 0.728]** |
| | LUNAR-OS | 0.651 [0.599, 0.714] | 0.650 [0.597, 0.716] | 0.654 [0.601, 0.717] |
| | LUNAR-SCE | 0.665 [0.605, 0.721] | 0.660 [0.601, 0.716] | 0.670 [0.610, 0.731] |
| | LUNAR | **0.670 [0.606, 0.723]** | **0.669 [0.606, 0.726]** | 0.674 [0.607, 0.730] |
| | Q7B | 0.620 [0.555, 0.679] | 0.647 [0.574, 0.715] | 0.609 [0.553, 0.664] |
| PPMI | Q3B | 0.315 [0.306, 0.324] | 0.510 [0.505, 0.515] | 0.552 [0.526, 0.576] |
| | LUNAR-OS-SCE | 0.398 [0.388, 0.407] | 0.512 [0.507, 0.517] | 0.564 [0.537, 0.594] |
| | LUNAR-OS | 0.373 [0.364, 0.383] | 0.508 [0.503, 0.513] | 0.545 [0.516, 0.575] |
| | LUNAR-SCE | **0.410 [0.401, 0.421]** | **0.515 [0.510, 0.521]** | **0.583 [0.555, 0.612]** |
| | LUNAR | 0.388 [0.379, 0.399] | 0.513 [0.508, 0.519] | 0.576 [0.546, 0.603] |
| | Q7B | 0.250 [0.240, 0.259] | 0.509 [0.503, 0.514] | 0.537 [0.512, 0.559] |

**Table S19**: **Per-class performance on the NP-ONE task.** Precision, recall and F1-scores (bootstrap median and 95% confidence intervals) for the NP-ONE task across models (Qwen2.5-3B-Instruct or Q3B, LUNAR-OS-SCe, LUNAR-OS, LUNAR-SCe, LUNAR (Ours) and Qwen2.5-7B-Instruct or Q7B). Results are shown for the internal NACC testing set. **Bold** values indicate the highest score and underlined values indicate the second highest score for each class–dataset combination.

| Class | Model | Precision | Recall | F1-score |
|---|---|---|---|---|
| None | Q3B | 0.160 [0.106, 0.216] | 0.082 [0.054, 0.113] | 0.109 [0.072, 0.147] |
| | LUNAR-OS-SCe | 0.329 [0.212, 0.452] | 0.066 [0.041, 0.096] | 0.110 [0.069, 0.155] |
| | LUNAR-OS | 0.394 [0.286, 0.509] | 0.089 [0.061, 0.120] | 0.145 [0.101, 0.192] |
| | LUNAR-SCe | 0.317 [0.266, 0.367] | **0.322 [0.276, 0.379]** | **0.319 [0.274, 0.370]** |
| | LUNAR | 0.314 [0.260, 0.371] | 0.264 [0.216, 0.310] | 0.287 [0.238, 0.334] |
| | Q7B | **0.410 [0.335, 0.490]** | 0.228 [0.183, 0.277] | 0.294 [0.239, 0.346] |
| AD | Q3B | 0.607 [0.574, 0.634] | 0.719 [0.688, 0.747] | 0.658 [0.631, 0.681] |
| | LUNAR-OS-SCe | 0.631 [0.604, 0.657] | **0.889 [0.868, 0.908]** | **0.738 [0.717, 0.757]** |
| | LUNAR-OS | 0.627 [0.600, 0.656] | 0.853 [0.832, 0.876] | 0.723 [0.702, 0.744] |
| | LUNAR-SCe | 0.734 [0.701, 0.763] | 0.629 [0.598, 0.658] | 0.677 [0.651, 0.702] |
| | LUNAR | **0.761 [0.728, 0.794]** | 0.598 [0.566, 0.629] | 0.669 [0.643, 0.695] |
| | Q7B | 0.653 [0.625, 0.681] | 0.812 [0.785, 0.838] | 0.724 [0.699, 0.745] |
| LBD | Q3B | 0.200 [0.146, 0.259] | 0.243 [0.177, 0.309] | 0.219 [0.163, 0.279] |
| | LUNAR-OS-SCe | 0.281 [0.226, 0.339] | 0.454 [0.377, 0.531] | 0.347 [0.289, 0.403] |
| | LUNAR-OS | 0.268 [0.217, 0.321] | 0.490 [0.407, 0.574] | 0.347 [0.287, 0.407] |
| | LUNAR-SCe | 0.265 [0.215, 0.315] | 0.574 [0.496, 0.654] | 0.362 [0.305, 0.419] |
| | LUNAR | **0.298 [0.245, 0.351]** | **0.606 [0.528, 0.685]** | **0.399 [0.336, 0.456]** |
| | Q7B | 0.251 [0.199, 0.304] | 0.452 [0.374, 0.533] | 0.322 [0.264, 0.378] |
| FTLD | Q3B | 0.582 [0.538, 0.626] | 0.528 [0.487, 0.574] | 0.554 [0.515, 0.593] |
| | LUNAR-OS-SCe | **0.789 [0.742, 0.836]** | 0.448 [0.404, 0.493] | 0.572 [0.530, 0.615] |
| | LUNAR-OS | 0.777 [0.728, 0.822] | 0.454 [0.408, 0.496] | 0.574 [0.530, 0.614] |
| | LUNAR-SCe | 0.594 [0.551, 0.643] | 0.541 [0.501, 0.584] | 0.567 [0.529, 0.605] |
| | LUNAR | 0.544 [0.508, 0.585] | **0.651 [0.610, 0.692]** | **0.592 [0.560, 0.625]** |
| | Q7B | 0.755 [0.708, 0.803] | 0.455 [0.410, 0.496] | 0.568 [0.526, 0.605] |

| Domain | Question |
|---|---|
| Etiology ID | Which summary more accurately identifies the most likely primary etiology and any mixed pathology (if evident), based on the provided information? |
| Biomarker | Which summary more effectively uses biomarker evidence (e.g., amyloid CSF/PET, DaTscan if available) and/or cognitive testing to justify the stated etiologic conclusions? |
| Clinical Link | Which summary shows clearer linkage between clinical findings, biomarkers (if available), and the final diagnostic impression? |
| Diff Dx | Which summary more appropriately limits the differential diagnosis to conditions supported by the evidence? |
| Clarity | Which summary is clearer, more concise and more actionable for neurologic decision-making? |

**Table S20**: **Evaluation domains and questions for comparison of clinician-only versus clinician+AI diagnostic summaries.** These five structured domains and corresponding questions were used by expert neurologists to systematically compare the quality of differential dementia diagnosis summaries generated with and without AI assistance. The domains assess etiological precision, biomarker interpretation, clinical coherence, diagnostic specificity and overall clarity/actionability to support neurologic decision-making.
